# Controlled colonization of the human gut with a genetically engineered microbial medicine

**DOI:** 10.1101/2024.10.03.24314621

**Authors:** Weston R. Whitaker, Zachary N. Russ, Elizabeth Stanley Shepherd, Lauren M. Popov, Alexander Louie, Kathy Lam, David M. Zong, Clare C. C. Gill, Jeanette Gehrig, Harneet S. Rishi, Jessica A. Tan, Areta Buness, Janeth Godoy, Domenique Banta, Sonia Jaidka, Katheryne Wilson, Jake Flood, Polina Bukshpun, Richard Yocum, David N. Cook, Tariq Warsi, Lachy McLean, Justin L. Sonnenburg, William C. Deloache

## Abstract

Precision microbiome programming for therapeutic applications has been limited by challenges in achieving reproducible colonization of the colon. Previously, we used a porphyran prebiotic to create a synthetic niche to engraft engineered bacteria into diverse microbiota in mice. Here we extend that work with biocontainment that links essential gene expression to porphyran presence yielding a platform for controlled colonization and decolonization of humans with engineered *Bacteroides*. We engineered this chassis with a five-gene oxalate degradation pathway, creating a therapeutic candidate that reduces hyperoxaluria, a cause of kidney stones, in pre-clinical models. Our Phase 1/2a clinical trial demonstrates tunable and reversible engraftment in humans, shows promising oxalate reductions, highlights addressable challenges in this novel modality for therapeutics, and queries key questions in microbiome science.

## Main Text

Evolved to harness diverse functions from its resident microbiota, the lower gastrointestinal tract offers a biologically meaningful but compartmentalized site for deploying engineered cell therapies. Pioneering efforts to realize this potential have taken the direct approach of delivering the maximally tolerated dose of auxotrophic bacteria engineered to express therapeutic activities while transiently inhabiting the gut (*1-8*). Despite showing efficacy in animals and a good safety profile in humans, these therapeutics have struggled to show clinical efficacy.

To enable the large physiological impact needed for many therapeutic applications, we introduce the approach of stably colonizing the gut at a high level with bacteria engineered to robustly perform a therapeutic function. We have focused on engineering gut-resident species from the *Bacteroides*, one of the most abundant genera in the industrialized gut that is well studied and genetically manipulatable (*9-14*). Previously our group engineered a *Bacteroides* strain with high-density, tunable engraftment capability by adding genes that encode utilization of the rare seaweed-derived polysaccharide porphyran (*15*). In mice, this synbiotic is capable of overcoming colonization resistance, even from established isogenic strains that lack the porphyran utilizing functionality (*15, 16*). While a robust engraftment technology is necessary, it alone is not sufficient to enable an engineered microbial cell therapy due to the need to reversibly control colonization levels through a biocontainment mechanism. Here we present a novel platform for controllably delivering therapeutic activities to the human gut that we use to develop a candidate hyperoxaluria therapeutic and show the results of Phase 1 and 2a clinical trials.

## Results

### Colonization by bacteria engineered to degrade oxalate reduces urine oxalate in rat hyperoxaluria models

Enteric hyperoxaluria (EH) is a disease caused by pathological over-absorption of dietary oxalate, often leading to recurrent calcium oxalate kidney stones (*17*). Previous attempts to treat EH by colonizing patients with natively occurring oxalate-consuming commensals found engraftment to be challenging (*18*). No oxalate consuming *Bacteroides* have been described previously. To engineer oxalate consumption into *Bacteroides vulgatus (Bv)*, we introduced an oxalate:formate transporter, as well as a pathway for conversion of oxalate to formate (Fig. 1A). We screened enzymes for an optimized expression of different 5 or 7 gene metabolic pathways to create strains able to rapidly degrade oxalate into formate at a rate of up to 15 mM per hour when at the 10^10^ CFU/g densities expected in the gut (Fig. S1A-C). We observed a tradeoff between the oxalate degradation rate and growth rate in the presence of oxalate *in vitro*, which was not observed in growth conditions lacking oxalate (Fig. S1C-D).

**Fig. 1.**
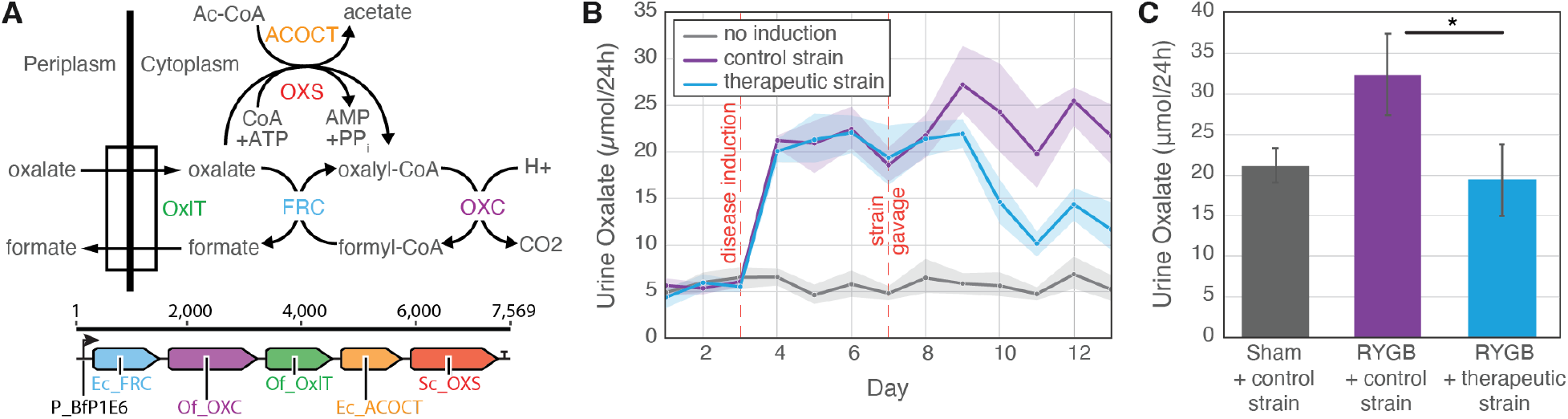
*Bacteroides* with engineered oxalate degradation reduce urine oxalate in rat hyperoxaluria models. (**A**) Oxalate degradation was engineered into a *Bacteroides* strain by introducing genes encoding oxalate transport (OxlT) and four enzymatic activities (OXS, ACOCT, FRC, and OXC). (**B**) High urine oxalate was induced (first dashed red line) in rats by incorporating spinach into their diet on day 3 (blue and purple). Strains with (blue) or without (purple) an oxalate degrading pathway were gavaged on day 7 (second dashed red line). 24-hour urine was collected for each rat and the average total urine oxalate (μmol/24h) and 95% confidence interval (shaded area) is plotted. (**C**) Compared to a sham surgery (grey), urine oxalate was induced via Roux-en-Y gastric bypass (RYGB) in rats subsequently colonized with a control (purple) or oxalate degrading (blue) strain; \**P* < 0.05.

In a dietary model of induced EH in rats, high density (7.2 × 10^8^ to 2.6 × 10^10^ CFU/ml) microbiota engraftment with porphyan-utilizing *Bv* strain containing the oxalate degradation pathway resulted in a 40% decrease in urine oxalate relative to a non-oxalate degrading control strain (Fig. 1B). When porphyran concentration was gradually increased, the corresponding colonization level and urine oxalate reductions were both increased (Fig. S2). Gastric bypass surgery is clinically associated with EH onset in people (*17*), and we further demonstrated our therapeutic strain reduces urine oxalate in a rat Roux-en-Y gastric bypass (RYGB) surgical model. The RYGB gastric bypass surgery resulted in a ∼50% increase in the urine oxalate in rats colonized by the control strain, an increase that was completely eliminated in animals harboring the oxalate degrading strain (Fig. 1C).

### Combining porphyran utilization and essential gene regulation enables reversible colonization

Reversible colonization is a key component of microbial cell therapies (*2, 5, 19, 20*). Dietary porphyran can create an exclusive niche for a porphyran-utilizing strains to enable predictable colonization of diverse microbiotas (*15, 16*). However, persistence of porphyran-utilizing strains after porphyran removal is difficult to predict due to the complexity of the ecosystem and broad range of nutrients *Bacteroides* can utilize. An effective and safe therapeutic platform would allow robust control of treatment duration and ideally enable total clearance of engineered cells from the gut microbiota.

To improve strain clearance without complicating treatment dosing, we engineered our strain to be more dependent on porphyran by replacing the native regulation of an essential gene with a porphyran inducible promoter (Fig. 2A). Using a short, 115-nucleotide promoter sequence from the porphyran polysaccharide utilization locus (PUL) (*21*), that can result in a >500-fold induction in response to porphyran (Fig. S3A). By replacing and tuning the native regulation of arginyl-tRNA synthetase (argS) or other essential genes, growth becomes dependent on the presence of porphyran, even in rich media where other carbon sources are available (Fig. S3). This conditional biocontainment approach has advantages of being difficult to complement by cross-feeding, being resistant to simple loss-of-function mutations, imposing a minimal fitness burden, and utilizing a rare, non-absorbable and safe dietary control molecule.

**Fig. 2.**
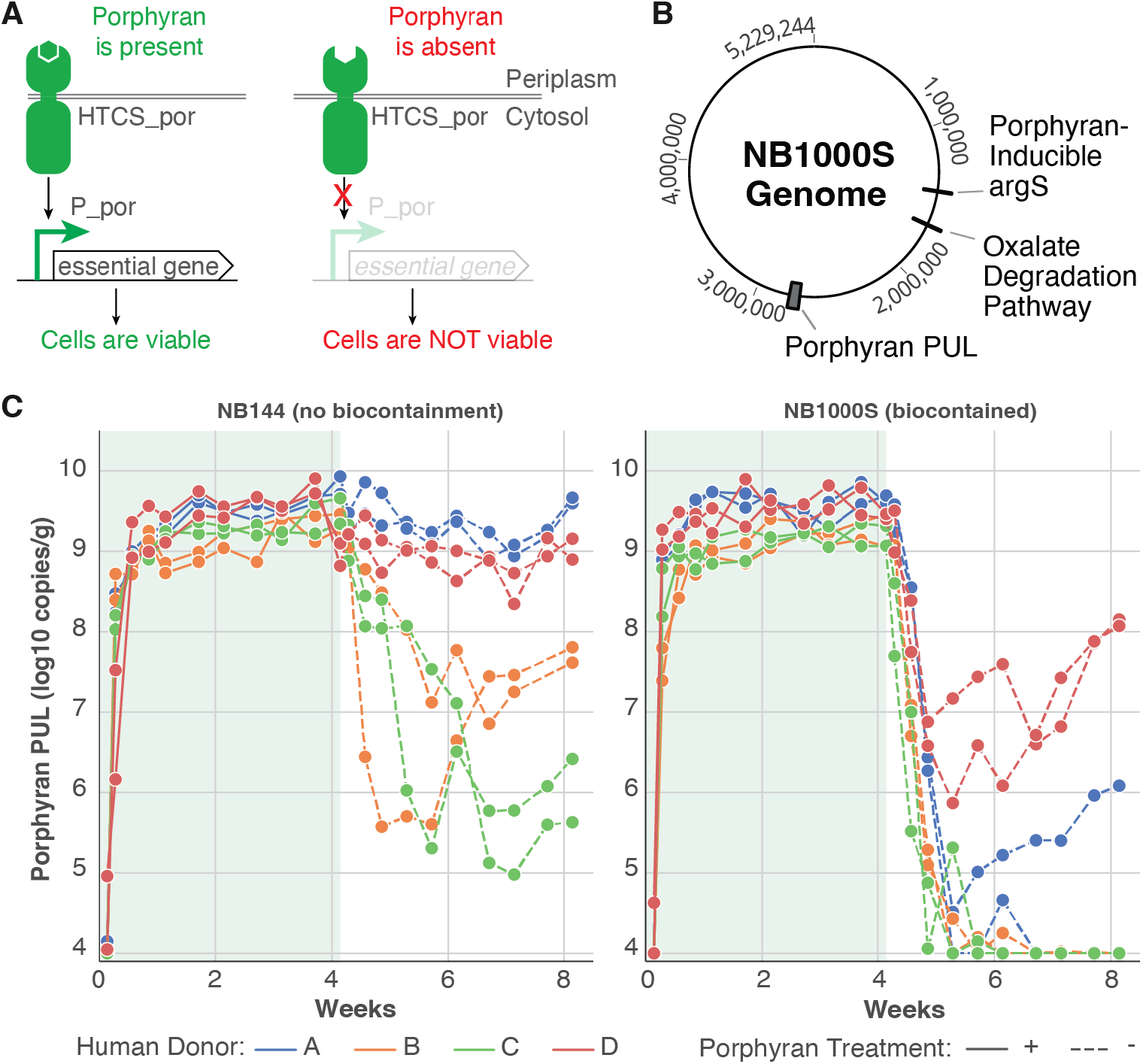
Combining porphyran utilization and essential gene regulation enables reversible colonization. (**A**) Biocontainment strategy uses a porphyran sensing HTCS to drive an essential gene. (**B**) Diagram of the NB1000S therapeutic strain containing biocontainment, an oxalate degradation pathway and a porphyran PUL. (**C**) Gnotobiotic mice harboring one of four human donor microbiotas were gavaged on day 1 with either a strain with a porphyran PUL, NB144 (left), or a therapeutic strain, NB1000S (right), containing a porphyran PUL and porphyran-dependent biocontainment of argS. Porphyran was supplied in the diet for the first 28 days. The copies of the porphyran PUL, corresponding to colonization level, are plotted for individual mice when porphyran is present (solid lines) or absent (dashed lines). The copies of the porphyran PUL dropped to the limit of detection, 10^4^ copies/g, in 8 of the 12 NB1000S-gavaged mice after porphyran was removed.

We added this biocontainment module to our oxalate degrading, porphyran-utilizing *Bv* strain to make NB1000S (Fig. 2B). NB1000S or a control strain carrying the porphyran PUL but lacking biocontainment or the oxalate pathway (NB144), were introduced into ex-germ-free mice harboring different healthy human donor gut microbiotas and fed a porphyran containing diet. After four weeks, removal of porphyran from the diet resulted in variable resting colonization levels of the non-biocontained strain between 10^5^ and 10^10^ copies/g (Fig. 2C, left panel).

Porphyran removal from the NB1000S colonized mice resulted in complete clearance of the strain from 5 of 8 mice (Fig. 2C, right panel). The remaining 3 mice that NB1000S continued to colonize demonstrated marked reduction in strain abundance upon removal of porphyran, before rebounding. Upon isolation of shed NB1000S cells from one of the mice that rebounded, we determined the strain contained a single point mutation (S811L) in the cytoplasmic region of the porphyran hybrid two component system (HTCS), consistent with a gain-of-function mutation that results in constitutive expression of the essential gene in the absence of porphyran.

To test the long-term genetic stability of NB1000S in the presence of porphyran we colonized conventional mice, periodically isolating and phenotyping isolates for oxalate degradation and biocontainment. We found colonization remained high and, although by day 421 52% of isolates lost oxalate degradation, no communities showed detectable biocontainment escapes (Fig. S4).

As human microbiomes reconstituted in gnotobiotic mice are likely to contain less diversity than the original human community, we sought to test if this imperfect level of biocontainment would be sufficient for clearance in a potentially more competitive human gut. Despite a potential for biocontainment escape, human testing of a strain of *Bacteroides* that has been engineered to degrade oxalate carries minimal risk since oxalate is not required for human physiological function, and natural oxalate-degrading bacteria and the genes used here exist in the gut microbiome of healthy individuals.

### Engineered Bacteroides controllably colonizes healthy volunteers

To test the safety, tolerability, and colonization performance of NB1000S in humans, we designed and ran study NOV-001-CL01, an adaptive Phase 1/2a clinical trial conducted in the US and Canada (Fig. S5-8). Further details of this study may be found in the Supplemental materials and at clinicaltrials.gov/study/NCT04909723. In Phase 1, a total of 39 healthy volunteer subjects were randomized to 7 study arms. Participants received the antacid *Alka Seltzer Gold* and NB1000S (one or two doses of 10^9^ CFU as frozen glycerol stocks) or placebo, administered orally. Subjects received 0g to 20g porphyran daily for 14 days upon strain or placebo dosing. NB1000S abundance in subject fecal samples was measured via a quantitative polymerase chain reaction (qPCR) of porphyran PUL and oxalate degradation sequences immediately before, during and for at least 8 weeks after cessation of porphyran dosing (Fig 3A).

**Fig. 3.**
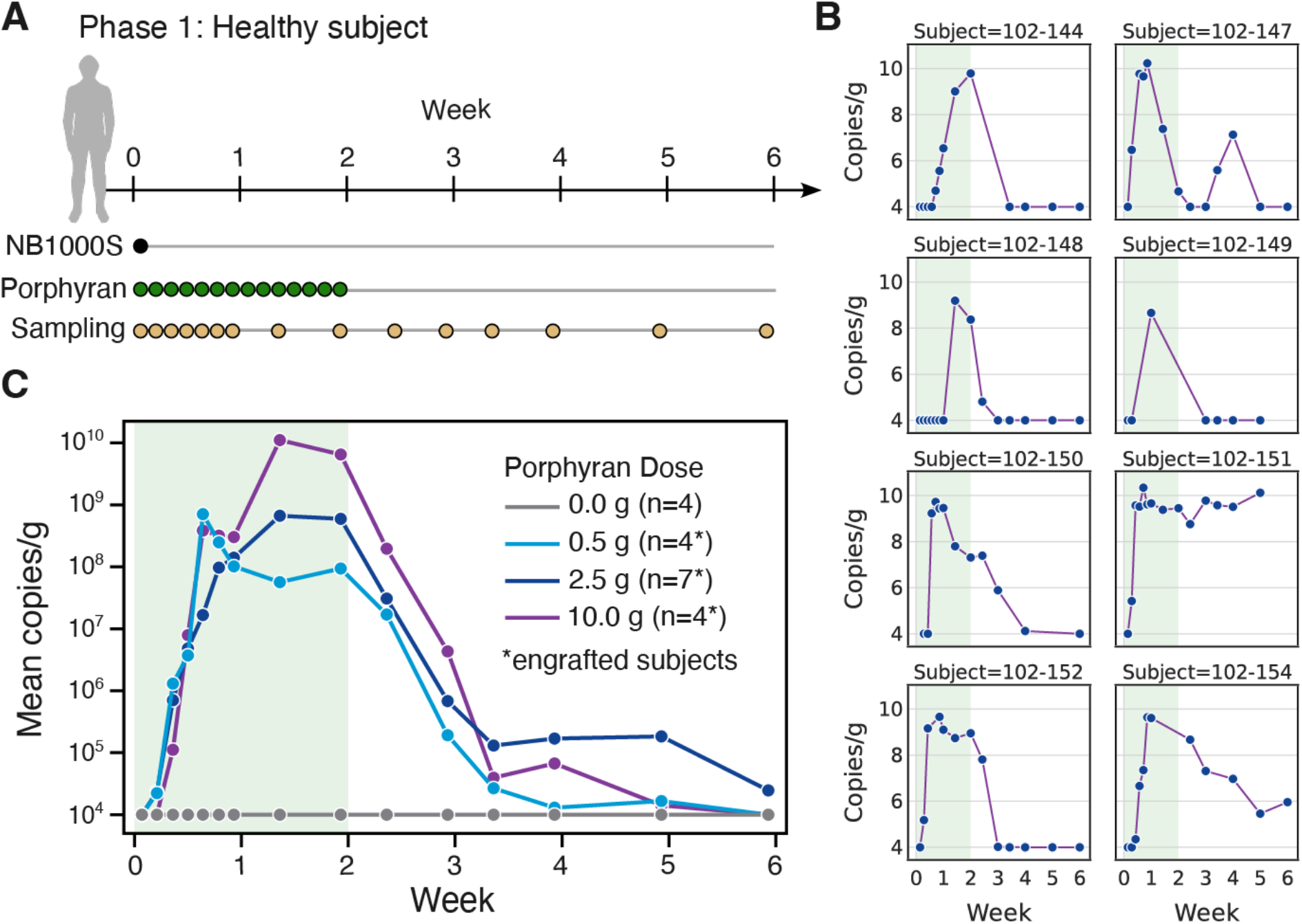
Porphyran-dose-dependent colonization of healthy volunteers by therapeutic strain, NB1000S. (**A**) Volunteers were administered a single dose of NB1000S on day 1 and administered porphyran daily for 2 weeks, with periodic sampling to assess colonization. (**B**) Copies of NB1000S per gram of feces are shown for all 8 individuals in the 2.5g porphyran cohort. Period of porphyran dosing is highlighted in green. (**C**) For engrafted subjects, defined as those that were colonized above 10^7^ copies/g on day 14, mean NB1000S copies per gram of feces is shown for the four cohorts given different daily doses of porphyran.

In early groups dosed with NB1000S, colonization was variable, with half of subjects failing to engraft (engraftment defined by NB1000S detected in stool at ≥10^7^ copies/g on the last sample during porphyran dosing) (Fig. S9A). Upon addition of a 40mg dose of Omeprazole extended release (a proton pump inhibitor) prior to dosing NB1000S, colonization improved substantially with 7 of 8 subjects becoming engrafted (Fig. 3B). When comparing only engrafted subjects across all groups, strain colonization density showed dose responsiveness to porphyran, with an approximately 10-fold increase in NB1000S for each 5-fold increase in porphyran (Fig. 3C).

Upon cessation of porphyran administration, NB1000S fecal abundance dropped below the limit of detection (LOD) in 15 of 19 engrafted subjects in total from the 4 groups dosed with NB1000S (Fig. S9). Six subjects that showed NB1000S measurements below LOD participated in a porphyran rechallenge after 56 to 98 days of no porphyran (‘wash-out’), consuming either 10g of porphyran or 5.2g of seaweed snacks (which contain porphyran) daily for 7 days. In all six subjects, levels of NB1000S in stool remained undetectable upon porphyran rechallenge, suggesting clearance of the strain was achieved.

NB1000S was still detectable in the stool of four subjects 56 days after cessation of porphyran administration. These subjects were treated with a 7-day course of oral antibiotics, which failed to clear NB1000S in most patients (Fig. S10) despite the strain’s sensitivity to the antibiotics (Table S2).

In all four persistently colonized subjects, we successfully established the mechanism of NB1000S biocontainment escape via genomic sequencing of fecal isolates. In all cases, NB1000S harbored a distinct genomic mutation causing the conditional attenuation system to lose responsiveness to porphyran. In two cases, the HTCS contained point mutations that are likely activating, consistent with observed escape mechanisms observed in our humanized mouse study. In the other two subjects, genome rearrangements were present, placing other promoters immediately upstream of the essential gene.

Adverse events (AEs) were mostly mild and transient, with a predominance of events of a gastrointestinal nature such as diarrhea and flatulence (Table S5-S6). There were no severe or serious AEs attributable to NOV-001. There was no apparent relationship between product dose, engraftment or persistent fecal shedding by NB1000S and the incidence or severity of AEs. Consistent with the synthetic metabolic niche engraftment approach in mice (*15, 16*), metagenomic profiling of subject feces before, during and after treatment revealed no significant change in alpha diversity of the native microbiota.

### Redundancy prevents biocontainment escape mutants

Upon observation of biocontainment escape mutants in both humanized mice and healthy volunteer subjects in our clinical trial, we sought to further characterize escape mechanism and decrease escape rate. We inoculated a biocontained strain into a chemostat initially containing porphyran, and after two days of ‘wash-out’ with media lacking porphyran we observed a large drop in abundance followed by takeover by biocontainment escapes (Fig. 4A). Whole Genome sequencing (WGS) of 385 escape mutants revealed 95% were driven by point mutations resulting in constitutive activation of the HTCS (Fig. S11A). The remaining 5% of mutants revealed genomic rearrangements or transposon insertion immediately upstream of argS, likely inserting an alternate promoter to drive argS. These escape mechanisms were consistent with those observed in the humanized mice and human subjects.

**Fig. 4.**
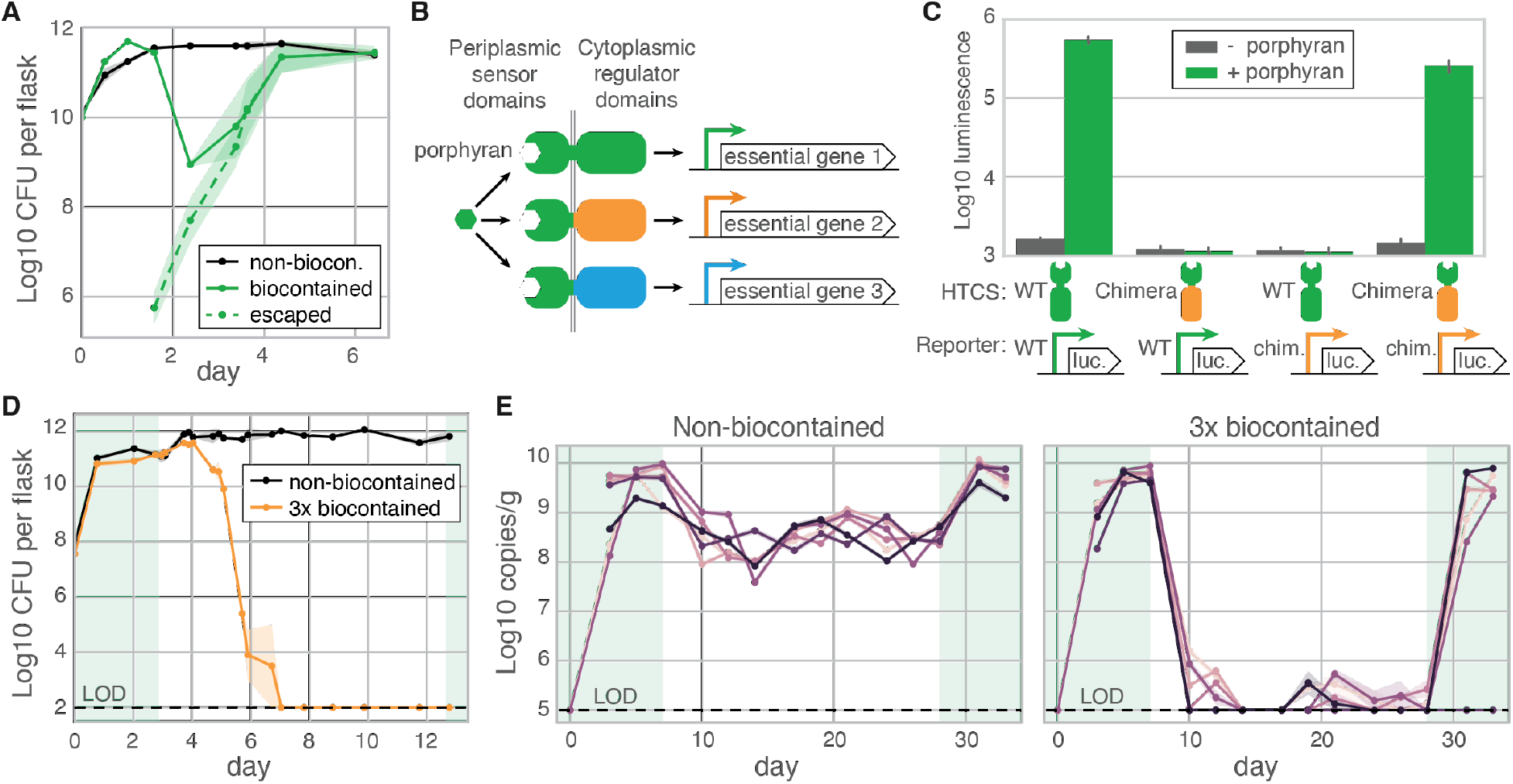
Additional layers of biocontainment via chimeric porphyran sensors improve attenuation. (**A**) Chemostats initially containing porphyran were inoculated with a strain with either no biocontainment (black) or one layer of biocontainment (green) and then continuously diluted by media lacking porphyran. The mean per chemostat colony forming units (CFU) on porphyran containing (solid) or porphyran lacking (dashed, associated with biocontainment escape) plates are shown. (**B**) Heterologous regulator domains and corresponding promoters were used to create chimera HTCSs capable of independently drive porphyran-dependent expression of three essential genes. (**C**) The wild type (WT) and a chimeric HTCS show more than a 100-fold induction in response to porphyran (green) only in their corresponding promoter, with no observed crosstalk. (**D**) Chemostats were switched from dilution with porphyran containing media (green background) to media without porphyran on day 3, and average CFU on porphyran containing plates is plotted for non-biocontained (black) or triple-biocontained (orange) strains. (**E**) Strain abundance was monitored for mice colonized with porphyran PUL containing strains without biocontainment (left) or with triple-biocontainment. Porphyran was supplied in the diet in the periods highlighted in green.

As all observed escaped isolates appear to be the result of rare gain-of-activity mutations to the HTCS or rearrangements specific to the essential gene promoter, we attempted to improve our biocontainment strategy such that multiple independent rare mutations would be required for escape. We hypothesized that engineering the porphyran sensor domain to drive three different essential genes with three unrelated orthogonal promoters (Fig. 4B) would provide a system requiring three independent rare mutations to achieve escape, resulting in vanishingly small probability of mutational escape. We generated chimeric HTCSs (*22*) using porphyran sensors (*21, 23*) and regulatory domain/promoter pairs from the PULDB database (*24*) that were rarely found in other *Bacteroides*, resulting in functional but weak (2-15 fold) porphyran activated chimeras (Fig. S11C). Additional tuning of the HTCS expression and by mutagenesis of the chimeric HTCS at the fusion point resulted in orthogonal chimeras with >100-fold porphyran induction (Fig. 4C and Fig S11). The chimeras were used to create a strain with three layers of biocontainment (3xBC), which cleared from a chemostat and showed no rebound in growth upon reintroduction of porphyran for an additional week (Fig. 4D).

We tested the 3xBC strain for clearance in conventionally raised mice, and when porphyran is removed, a non-biocontained strain decreased in density ∼1-2 logs, whereas the 3xBC strains decreased 4 logs to the limit of detection (Fig. 4E). Despite 3 weeks in the absence of porphyran and most mice showing undetectable fecal levels of the 3xBC strain, 5 of 6 animals displayed strain rebound upon reintroduction of porphyran (Fig. 4E). 3xBC strains isolated from the persistently colonized animals remarkably showed no mutations by WGS and phenotypically behaved as biocontained *in vitro* (Fig. S12A). In mice humanized with permissive microbiotas when porphyran was removed the 3xBC strain appeared more attenuated than the 1xBC strain, but similarly failed to clear (Fig. S12B). Recreating and testing 2xBC in three other *Bacteroides* species resulted in the same persistence without mutation (Fig. S12C). We ran more than 100 experiments to determine the non-mutational means of persisting in the absence of porphyran, including RNAseq in germ-free mice and Tn-Seq screens, but we could not determine the mechanism of *in vivo* persistence nor identify any genetic escapes from the 3xBC strains.

### EH patient colonization revealed unique insights into commensal horizontal gene transfer (HGT) in humans

In parallel to our biocontainment improvement efforts and after demonstrating successful and safe engraftment in healthy volunteers, NB1000S moved into patients with EH secondary to RYGB or to Biliopancreatic Diversion with Duodenal Switch (BPD-DS) bariatric surgical procedures in Phase 2 of study NOV-001-CL01. Nine EH patients received NB1000S (2 doses of 10^9^ CFU orally) followed by 10g of porphyran daily for 28 days; 3 patients received placebo (Fig. 5A, S7). NB1000S engraftment was less consistent and overall reached a lower density in patients as compared to healthy volunteers (Fig. S13), possibly related to the strain’s fitness defect observed only in the presence of high oxalate *in vitro* (Fig. S1E) or in patient 103-203 due to prior colonization with a porphyran utilizing strain (Fig. S14A).

**Fig. 5.**
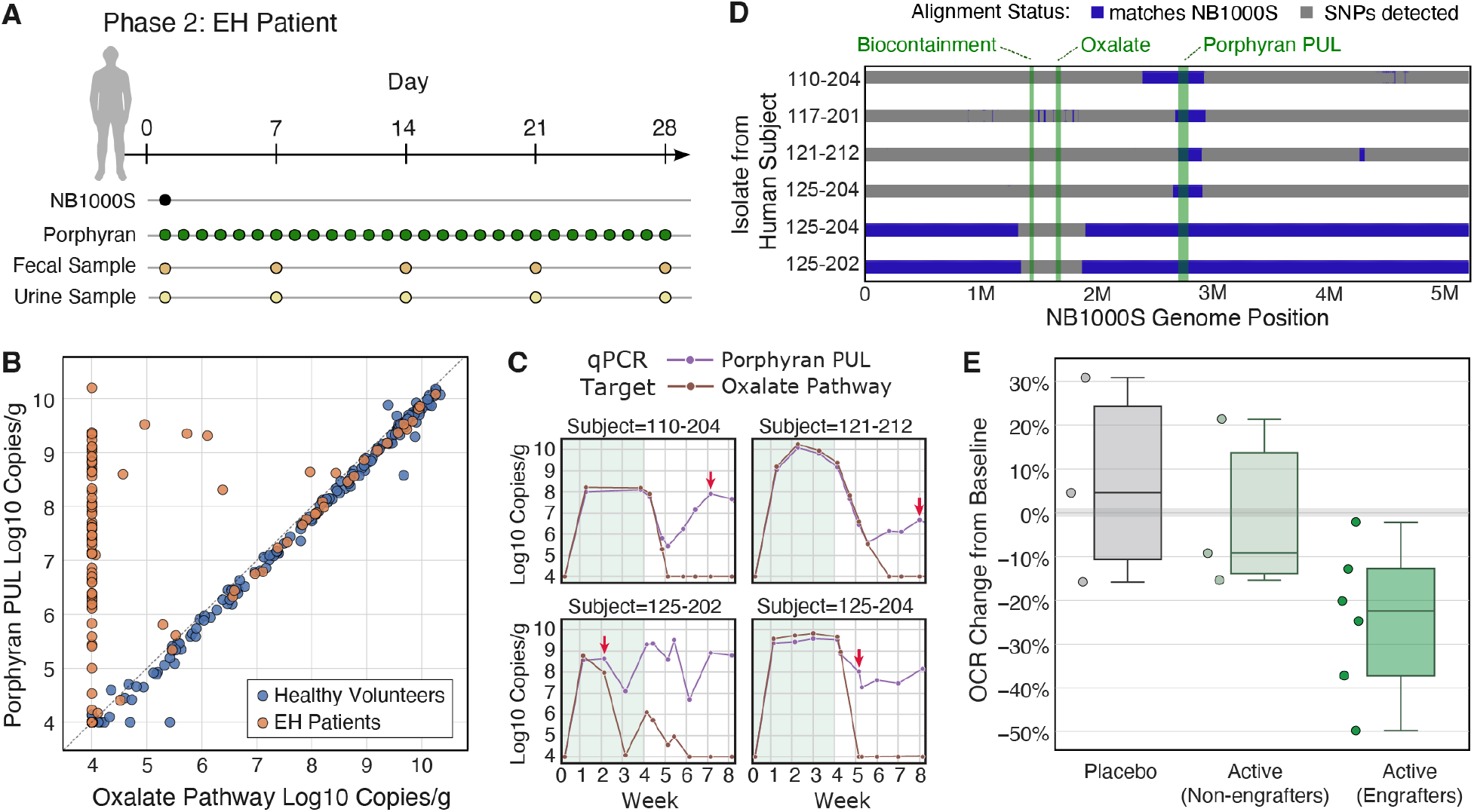
In hyperoxaluria patients, but not healthy volunteers, large horizontal gene transfer events were observed. (**A**) In Phase 2, EH patients that had prior RYBG or BPD-DS treatment and exhibited high levels of urine oxalate were administered NB1000S on day 1 and porphyran daily. Once per week fecal and 24-hour urine samples were taken. (**B**) Copies of the porphyran PUL and the oxalate degradation pathway are plotted for the healthy volunteers, Phase 1 (blue), and the hyperoxaluria patients, Phase 2 (orange). Dashed line indicates equal abundance. (**C**) Copies of the oxalate pathway (brown) and porphyran PUL (purple) are plotted for representative patients. Daily porphyran dosing period is highlighted in light green. Red arrows indicate point of strain isolation. (**D**) The genomes of strains isolated from Phase 2 patients are compared to the original NB1000S strain (matches in blue). Introduced loci locations are highlighted green. Regions with single nucleotide polymorphisms likely from a non-NB1000S source are shown in grey. (**E**) Percent change in baseline urine oxalate to creatinine ratio (OCR) is shown for placebo (grey), non-engrafted patients falling below 10^7^ oxalate pathway copies/g on last porphyran day (light green), and the remaining engrafted patients (dark green; P = 0.08 vs Placebo).

As with healthy volunteers, NB1000S abundance was tracked in patients via qPCR of both unique porphyran PUL and oxalate pathway sequences. However, unlike in healthy volunteers where the oxalate pathway and porphyran PUL sequences were very closely correlated, many patient samples showed substantially higher PUL copy number than the oxalate pathway (Fig. 5B). This discrepancy appears prior to strain dosing (subject 103-203), during porphyran treatment (subject 125-202) or shortly after porphyran is removed (subjects 121-212, 110-204, 117-201, 125-204) (Fig. 5C; Fig. S14A). Two subjects (103-201 and 121-216) show no discrepancy. To determine the reason for this discrepancy, 6 strains harboring the porphyran PUL were isolated from 5 different patients (timepoints indicated in Fig. S14) and underwent WGS. Comparative sequence analysis to NB1000S (Fig. 5D; SNP regions show in blue) revealed two NB1000S strains had incoming 667 or 724 kilobase HGT that fully replaced the oxalate pathway and biocontainment. The other 3 strains were the native *Bacteroides* that took up the porphyran PUL from NB1000S through 216 to 561 kb HGT events. Using the samples before NB1000S was dosed and unique qPCR primers for the HGT recipients, we also isolated the native *Bacteroides* strains before they received the porphyran PUL. Comparative analysis (Fig. S14) identified DNA not present in NB1000S or the native parent strain, which included an ICE (integrative and conjugative element) possibly facilitating to an outgoing HGT (*25*). Experimentally introducing an ICE element into NB1000S significantly increases the rate of outgoing HGT *in vitro* (Fig. S15A), which can be substantially reduced when elements of the PUL are genomically split into 2 pieces separated by 200 kb (Fig. S15B).

All five predefined analyses of urine oxalate for efficacy showed a directional but statistically insignificant improvement in the treated patients compared to placebo. *Post hoc* analysis of the urine oxalate to creatinine ratio (OCR) showed a 27% reduction comparing all treated patients to placebo (P=0.13; Fig. 5E) or comparing engrafted patients to placebo and non-engrafted patients (P=0.03; Fig. S15F). Among both phases of NOV-001-CL01, there were no product-related serious AEs and no subjects left the study or required dose adjustment due to safety or tolerability issues (Table S5-8).

## Discussion

In a first-in-human study of this novel class of controllably colonizing therapeutic bacteria, we tested our oxalate-degrading clinical candidate strain and paired porphyran polysaccharide in healthy subjects and patients with EH. We demonstrated it is possible to colonize humans with an engineered gut commensal for a sustained period of time at high levels. Remarkably, a single dose of the strain is sufficient to enable colonization if provided proper gastric protection, and even at high doses of porphyran the synbiotic treatment appeared safe and well tolerated.

Because the human gut is a highly competitive environment, it was unclear if biocontainment would be required for strain clearance. Our data demonstrate that a strain with a single biocontainment layer can escape in humans via similar mutations seen in preclinical models, leading to persistent colonization. Adding two additional layers of biocontainment successfully prevented emergence of escape mutations, but for cryptic—perhaps epigenetic or ecological—reasons, though attenuated, the strain may still persist in highly permissive microbiotas.

Unlike in healthy volunteers, our strain colonized at more variable levels in EH patients and underwent large HGT events that separated the porphyran PUL from the other engineered elements in 5 of the of 7 patients, which is expected to impact durability of the treatment (*26*). Patients were selected to have high urine oxalate levels, and oxalate imposes a fitness burden for our therapeutic strain, which may explain those results, though other factors associated with gastric bypass may also be relevant.

Though biocontainment and genetic stability issues limited the potential effectiveness of NB1000S, we saw promising, though inconclusive, directional reductions in urine oxalate levels. A future strain with reduced oxalate-associated fitness burden to improve patient colonization level and genetic stability could be a promising treatment for EH. Additionally, although clearance is currently imperfect, finding no breakage of 1x biocontainment in colonized mice after 421 days on porphyran suggests that biocontainment could be a promising approach to clearing out and replacing strain that have broken therapeutic activities due to modest fitness defects.

Taken together, these data serve as an important proof-of-concept for the future development of engineered bacterial cellular therapies. As causal molecular mechanisms underlying associations between the gut microbiota and human disease become clearer, this platform offers a powerful tool to deliver defined therapeutic activities robustly to the human colon. It may also be possible to effectively combine multiple engineered activities into a single therapeutic strain to better treat complex diseases without substantially increasing the complexity of manufacturing, trial design and other downstream activities.

## Data Availability

All data produced in the present study are available upon reasonable request to the authors.

## Author contributions

WRW, ZNR, ESS, LMP, AL, KL, DMZ, JG, HSR, JAT, KW, JF, JLS, PB, RY, DNC, TW, LM, WCD conceived of the project or study plans. WRW, ZNR, ESS, LMP, AL, KL, DMZ, CCCG, JG, HSR, JAT, AB, JG, DB, SJ, KW, JF, PB, LM, WCD were all involved with data acquisition and or interpretation. WRW, ZNR, ESS, LMP, KL, CCCG, SJ, KW, JF, PB, TW, LM, WCD were involved in supervision and administration. WRW wrote the manuscript with input from all authors.

