## Supplemental Materials for "Controlled colonization of the human gut with a genetically engineered microbial medicine"

**The PDF file includes:**

Materials and Methods
Supplementary Text
Figs. S1 to S16
Tables S1 to S9
Supplemental References

**Materials and Methods**

Bacteria culturing

Strains were grown at 37˚C within a Coy anaerobic growth chamber containing a gas mix of 5% $H_2$, 10% $CO_2$ and 85% $N_2$. Brain Heart Infusion supplemented with hemin and menadione (BHIS) in liquid or on plates was used as the primary media in strain constructions and "rich media" in vitro assays. To prepare BHIS, 37 g/L of Bacto Brain Heart Infusion (BD Diagnostics) was added to distilled water, autoclaved, cooled to below 55˚C and supplemented at a 1:1000 dilution with hemin (200mg NaOH, 250mg hemin, 50 mL dH2O, mixed to dissolve, filter sterilized and stored at 4˚C) and menadione (50mg menadione and 50 mL 200 proof ethanol filter sterilized and stored at 4˚C). Defined media was based on EZ MOPS Defined media (Teknova), was similarly supplemented with hemin and menadione and filter sterilized. Immediately before use, liquid media was reduced by supplementing with a 1:100 dilution of cysteine (2.5g L-cysteine and 50 mL dH2O, filter sterilized and stored at -20˚C). Solid media BHIS plates are prepared similarly to liquid media but also included of 15 g agar or agarose added prior to autoclaving. The porphyran PUL is induced by agar but not agarose. To grow biocontained strains, 0.5% porphyran extract was added to the liquid or solid media.

Construction of Bacterial Strains

A sample of *B. vulgatus* type strain DSM 1447 / ATCC 8482 was received from DSMZ (Braunschweig, Germany) and individual colonies were isolated and sequenced as described below. A couple of mutations were noted versus the reference sequence.

Strains (listed in Table S1) were constructed via conjugation of the parent Bacteroides strain with S-17 *E. coli* bearing a plasmid. Plasmids were constructed via Golden Gate reactions carried out according to standard procedures (*1*). Completed Golden Gate reactions of 5 µL were transformed with addition of 25 µL of chemically competent E. coli S17-1 (*2*) cells (mid-log cells resuspended 1:20 in TSS/KCM: LB medium with 8.3% PEG-3350, 4.2% DMSO, 58 mM MgCl2, 167 mM CaCl2 and 457 mM KCl), followed by a 90 second heat shock at 42 °C, recovery at 37 °C for 30 minutes, followed by plating on LB agar plates with the appropriate antibiotic and aerobic growth at 37 °C. Conjugation was done by mixing 50 uL of saturated S-17 *E. coli* culture and 50 uL of saturated *B. vulgatus* parent culture, concentrating the mixture to 10 uL, and plating on BHIS plates. The plates were incubated at 37˚C overnight, scraped, and the scraped mixture resuspended in sterile PBS before plating on selective BHIS media supplemented with 200 µg/mL gentamycin, and either 25 µg/mL erythromycin, 2 µg/mL tetracycline, or 34 µg/mL chloramphenicol to select for the indicated plasmid.

The porphyran PUL plasmid, pWD035 (*3*), was integrated into the genome using expression of NBU2 in the target Bacteroides cell, which leads to linearization of the plasmid adjacent the attN2 site, and integration of the linearized plasmid at one of several serine tRNA sites. Subsequent genetic modifications relied on allelic exchange using two homology regions of approximately 1500 base pairs instead. Removal of the vector backbones was achieved with expression of a mutant PheS phenylalanine-tRNA ligase that accepts 4-chloro-phenylalanine (4CP) as a toxic antimetabolite for counterselection against cells that retain the vector backbone, resulting in a clean allele substitution (*4*). Counterselection took place after BHIS with antibiotic selection on plates followed by a restreak and outgrowth onto BHIS plates with antibiotic. These isolated colonies were allowed to loop out the vector backbone by expansion in liquid BHIS without antibiotic, and saturated cultures were restruck onto minimal media plates prepared as described by Salyers et al. 1977 (*5*) supplemented with 2 mM 4CP. Correct deletions were identified via picking colonies that displayed the correct phenotype and confirmed via PCR and Sanger sequencing.

Strain WGS

Strains were isolated by streaking onto porphyran-containing BHIS and incubated for 24 hours at 37˚C. A single colony was picked into rich BHIS media containing 0.5% porphyran supplemented with 200 ug/mL gentamicin, and a culture was grown up overnight anaerobically at 37˚C.

The culture was immediately pelleted, subjected to DNA extraction using the Qiagen QIAamp Mini kit and subsequently prepared for next generation sequencing using the Nextera DNA Flex bead-linked tagmentation kit. The library was run on the Illumina iSeq 100 one-color benchtop sequencing instrument using iSeq i1 Reagent v1 cartridges.

A long-read sequencing library was prepared using the Oxford Nanopore Rapid Sequencing Kit (SQK-RAD004) following the manufacturer's instructions. The library was run on a Nanopore MinION Flow Cell (R9.4.1) connected to a MinION Mk1B sequencing device and a dedicated computer running the MinKNOW control software which was also used to perform local basecalling using default settings.

*De novo* assembly was performed using the Unicycler hybrid assembler (v0.4.9b) with default settings. All short and long read datasets were provided directly to the algorithm with no additional pre-processing steps. Due to the complex, repetitive nature of the *Bacteroides vulgatus* genome, it was necessary to perform manual polishing steps on the Unicycler *de novo* assembly contig prior to calling differences with the reference genome. To ensure that no SNPs were erroneously modified during manual NB1000S polishing, both the short and long read WGS data were re-mapped to the polished genome, yielding consistent read depth and no detectable variants.

Preclinical *in vitro* strain stability assessment

To assess genomic stability NB1000S was outgrown for ~100 generations. NB1000S was struck onto porphyran-containing BHIS plates incubated for 24 hours at 37˚C. Three colonies representing biological replicates were picked into rich BHIS media containing 0.5% porphyran, grown up overnight anaerobically at 37C and the resulting saturated culture was considered the starting point. 1 μL of saturated culture was diluted into 1 mL of pre-reduced rich BHIS media containing 0.5% porphyran grown anaerobically for 12 hours and similarly diluted every hour for a total of 10 dilutions over 120 hours. 40 colonies of each replicate were picked into rich BHIS media containing 0.5% porphyran and grown overnight to give 120 isolate cultures. Each culture was then assessed or growth on porphyran, biocontainment and oxalate degradation as described below.

Preclinical *in vitro* strain high throughput characterization

Porphyran utilization and conditional attenuation was assessed by comparative parallel sub-culturing of a saturated clonal culture into defined media containing either 0.2% glucose or 0.5% porphyran as the sole carbon. Porphyran utilization was confirmed by growth in the media containing 0.5% porphyran, while conditional attenuation was confirmed by the absence of growth in the media containing 0.2% glucose. $OD_{595nm} > 0.5$ were considered positive for growth.

An isolate was affirmed as oxalate-consuming using an oxalate degradation assay where bacteria are sub-cultured into rich media containing 0.5% porphyran supplemented with 6 mM sodium oxalate, and grown anaerobically at 37°C for 22 hours. Cultures were spun down and the remaining cell-free supernatants and media only blanks were analyzed using a commercially available colorimetric oxalate quantification kit, EnzyChrom Oxalate Assay Kit (Bioassay Systems, Hayward, CA). Absorption readings $OD_{595\ nm} < 0.5$ (at least ~80% of oxalate removed) were indicative of oxalate degradation.

<u>Preclinical *in vitro* strain oxalate degradation quantification</u>

Strains were struck onto porphyran-containing BHIS plates from glycerol stocks and incubated for 24 hours at 37˚C. Three colonies of each strain representing biological replicates were picked into rich BHIS media containing 0.5% porphyran, and cultures were grown up overnight anaerobically at 37˚C. Saturated cultures were then diluted 1:250 into fresh BHIS-Por media, grown for 22 hours to late log phase, concentrated 20X, and diluted 2X to approximately $10^{10}$ cells/mL into pre-reduced BHIS-Por media containing 4 mM or 8 mM oxalate under anaerobic conditions. After 10, 30, 45, and 80 minutes cultures were removed from the Coy, immediately spun down and supernatant was assayed for oxalate concentration. using the EnzyChrom Oxalate Assay Kit (Bioassay Systems, Hayward, CA), a colorimetric kit for quantitative oxalate measurement. All oxalate readings in the linear range for the indicated test conditions were used to calculate the oxalate consumption rate for each biological replicate.

<u>Preclinical *in vitro* strain reporter assays</u>

To measure luciferase reporter activity, colonies were picked into BHIS media and cultures were grown up overnight anaerobically at 37˚C. Saturated cultures were then diluted 1:500 into regular BHIS or BHIS containing porphyran at 0.5% and grown anaerobically for an additional 12 hours. Cultures were then removed from the anaerobic chamber and assayed. All cultures were saturated, with OD 595nm reading of between 0.90 and 1.05. Luminescence of each culture was then measured using the Nano-Glo Luciferase Assay System (Promega, Madison, WI) read on a Tecan Spark plate reader.

<u>Preclinical *in vitro* biocontainment growth curves</u>

To generate growth curves, colonies were picked into BHIS media containing porphyran at 0.5% and cultures were grown up overnight anaerobically at 37C. Cultures were then diluted 1:250 into rich BHIS media containing 0.5% porphyran and grown anaerobically an additional 22 hours to reach saturation. Saturated cultures were then spun down, resuspended in PBS and diluted 1:1000 into 80 µL regular BHIS or BHIS containing porphyran at 0.5% in a 384-well clear bottom plate. The cultures were then used to obtain a 24-hour growth curve where OD595 was measured every 5 minutes using a Tecan Sunrise plate reader set to 37˚C and housed in a Coy anaerobic growth chamber containing a gas mix of 5% $H_2$, 10% $CO_2$ and 85% $N_2$.

<u>Preclinical *in vitro* biocontainment plate assay</u>

To assess robustly determine biocontainment phenotype dilution plating on solid media was used. Fecal samples were dilution plated on BHIS agar plates supplemented with 0.5% porphyran, gentamycin and strain specific antibiotics (erythromycin and tetracycline) and incubated anaerobically at 37°C. Isolated colonies were picked into BHIS liquid media supplemented with 0.5% porphyran, incubated anaerobically at 37°C. 200 µL saturated cultures were spun down and washed three time in PBS, resuspended in 200 µL PBS, serially 10-fold 7 times, and then 3 µL of each diluted sample was plated onto BHIS agarose (not agar) plates with and without 0.5% porphyran supplementation. The most concentrated spots on the plates lacking porphyran typically show some weak outgrowth of lawns were disregarded, while spots in or near the countable range are quantified. Biocontained strains typically show robust individual colonies on the porphyran supplemented plates at even at five or more 10-fold dilutions further than the weak lawn growth on the plate that lacks porphyran.

Preclinical *in vitro* biocontainment chemostats

A miniaturized chemostat was assembled using 125 mL stoppered flasks with three 16Ga hypodermic needles each (one 5” and two 1.5”). The 5” needle was used as a gas/liquid outlet to set maximum media height (and thus volume) while the two 1.5” needles served as a nitrogen gas inlet and media inlet, respectively. A 1” stir bar was placed in each and set to the minimum speed (~100 rpm). The stoppered flasks were warmed to 37C via hotplate and kept pressurized and anaerobic by a continuous flow of nitrogen gas.

Glycerol stocks of Bacteroides strains were inoculated into 17 mL of BHIS with 0.5% porphyran and the appropriate antibiotics (25 mg/mL erythromycin) with or without anhydrotetracycline (aTc) and incubated anaerobically at 37C overnight. These cultures were then transferred into flasks and the volume raised to 100 mL with fresh BHIS media with 0.5% porphyran + 25 mg/mL erythromycin and aTc as needed. With a volume set point of 100 mL, up to 10L of new BHIS media (with 2 g/L added sodium bicarbonate to prevent depletion due to N2 flow, and with/without 0.5% porphyran) was pumped in at a rate of 0.1178 mL/min per flask using a Watson-Marlow 205S peristaltic pump (set to 5 rpm) and black/black (0.76 mm bore) Marprene tubing. The positive pressure from the nitrogen pushed spent media out into waste or sampling, and the effective dilution rate was ~0.1 volumes/hour, or approximately 1 volume change every 12 hours. Samples were plated at various dilutions onto BHIS plates formulated with either 1.5% porphyran-containing agar or 1.5% agarose as the gelling agent, with or without aTc added. Colony-forming units (CFUs) were measured to determine the number of surviving or escaped organisms. To validate complete strain clearance, porphyran was added back.

Preclinical *in vitro* HGT assay

To assay *in vitro* HGT rates 4 mL of saturated culture of donor and recipient were combined, spun down and resuspended in 500 µL PBS with cysteine, and 100 µL was spread over ~ 10 $cm^2$ on a prewarmed HGT inducing plate (defined media agarose plates with 0.05% porphyran and glucose) and grown anaerobically at 37°C overnight. The resulting lawn is scraped off with a sterile loop, resuspended in 1 mL PBS with cysteine, washed three times and resuspended in 500 µL PBS with cysteine. 50 µL of resuspended cells are diluted into 10 selective liquid media, grown anaerobically at 37°C for 2 days and streaked onto selective plates to obtain HGT isolates.

Preclinical *in vivo* rat handling for hyperoxaluria

Studies were conducted at a CRO vivarium (Explora BioLabs, located in South San Francisco, CA) in strict accordance with a protocol for Care and Use of Laboratory Animals approved by the Explora BioLabs IACUC. Sprague-Dawley rats weighing between 150-200 g (Taconic) were housed in pairs using the Innovive Disposable IVC Rodent Caging System, until being singly housed in Tecniplast metabolic cages.

To limit oxalate degradation during collection, 200 µL 6 M HCl was added to empty tubes, oxalate was collected for 24 hours and 200 µL 6 M NaOH was added to neutralize collected urine before storing at -20˚C. Strains were administered by a 1 mL dose of ~$10^9$ CFU saturated BHIS with porphyran bacterial culture that was added to the daily food ration. Fresh feces for bacterial enumeration were collected directly from the rats. In accordance with the Novome SOP, at times of fecal sampling, animal body condition was evaluated. Well-conditioned animals are defined by non-prominent vertebrae and dorsal pelvis that is palpable with slight pressure. General sickness behaviors to monitor include lethargy, ruffled fur appearance, alterations in nasal/skin/ocular appearance and secretions, and motor activity. Any deviations from normal body condition were noted. All animals were weighed on study days 1, 13, and 18.

Preclinical *in vivo* rat dietary HO induction

To induce SH in Sprague-Dawley rats weighing between 150-200 g (Taconic), a custom high-oxalate low-calcium (HOLC) diet was used composed of (per rat per day) 18 g Envigo Low Calcium (0.4%) Diet, 72 mg xanthan gum, 340 mg porphyran, 23 mL water and enough rinsed minced (0.1" to 0.25") spinach to reach 52 µmol oxalate. On study the two study days prior to disease induction, animals were restricted to ½ normal daily caloric rations with *ad libitum* water to increase appetite and facilitate dietary induction of hyperoxaluria. Secondary hyperoxaluria was induced from baseline in experimental animals by the addition of frozen spinach to the low-calcium diet (TD.160869, Teklad diet, Envigo). To achieve the intake of 52 µmol of oxalate—scaled to the average daily human consumption by kilocalorie—a variable amount of frozen spinach, approximately 2.7 g per rat daily, was used.

After at least 3 days of HOLC diet, 24-hr UOx measurements were assayed. To minimize starting differences in UOx individual animals were randomized to their experimental groups (*n* = 8 per strain) using an algorithm that randomly sampled possible group assortments and scores the groups to minimize differences in average baseline urine oxalate between groups over all measured days.

Preclinical *in vivo* rat dietary HO ascending porphyran study design for Figure S2

Two strains were utilized in this study: the control *Bv* strain NB124 containing only the porphyran utilization modification, and the porphyran -utilizing and oxalate-consuming research strain sWW627. A third no treatment control group was included (no strain and no porphyran).

24 male Sprague-Dawley rats were transferred into metabolic cages on study day 0. On day 2 secondary hyperoxaluria was induced from baseline in experimental animals by the addition of frozen spinach to the low-calcium diet. On day 7 of the study, animals were randomized into 8 per group dosed with the corresponding strain. Porphyran was included in the food starting on day 11 and was gradually increased over the remainder of the study according to the following schedule: 0%, days 7-10; 0.05% days 11-13; 0.17% days 14-16; 0.5% days 17-19; 1.5% days 20-22. On study days 8, 9, 10, 14, 17, 20, and 23, fecal samples were collected to enumerate bacterial density in feces. On day 23, animals were euthanized.

Preclinical *in vivo* rat dietary HO high porphyran study design for Figure 1B

Two strains were utilized in this study: the control *Bv* strain NB124 containing only the porphyran utilization modification, and the porphyran-utilizing and oxalate-consuming research strain sWW554. A third no hyperoxaluria induction control group was included.

18 male Sprague-Dawley rats were transferred into metabolic cages on study day 0. On day 2 secondary hyperoxaluria was induced from baseline in 12 experimental animals by the addition of frozen spinach to the low-calcium diet. On day 7 of the study porphyran was included in the

food for the remained of the study, the induced animals were randomized into 6 per group dosed with the strain corresponding to their group.

Preclinical *in vivo* rat RYGB HO induction

20 male Sprague-Dawley rats at 11 weeks of age and 20 male Sprague-Dawley rats at 12 weeks of age (Taconic) were ordered to The Mouse Biology Program at the University of California, Davis. Prior to surgical manipulation, animals were subject to a dietary fattening phase of 4 weeks on Teklad TD.06414 chow. The animals arriving at 12 weeks of age were placed on TD.06414 chow immediately, and the animals arriving at 11 weeks of age were placed on TD.06414 chow one week later (at 12 weeks of age). Animals underwent surgical manipulation at 16 weeks of age, after 4 weeks on the high fat diet. Animals were targeted for a Body Condition Score of 4 in order to proceed with surgery.

Under the supervision of expert UC Davis veterinary staff, 35 animals underwent RYGB surgery, and 5 were subjected to a sham procedure. In the sham procedure, both the stomach and intestines are exteriorized and a superficial line of suture is applied along the same line on the stomach where the suture barrier is applied in the RYGB intervention; further, a small incision in the intestine is applied and then sutured closed at the same anatomical site where the intestine is bisected for RYGB animals.

Following surgery, animals recuperated at MBP for ~3 weeks, while body condition, weight, and glucose monitoring was evaluated. Due to adverse complications from the RYGB surgical procedure, 10 animals met the criteria for humane euthanasia prior to shipment. The UC Davis veterinary staff attributed the low post-surgical survival rate to an unexpected complication from the recovery diet and paper bedding, which were corrected during the recuperation period. The 25 surviving animals were weighed at the time of packing and shipped to Novome's Vivarium CRO, Explora BioLabs in South San Francisco by special courier.

Animals were fed a low-fat diet for study days 0-3 during an initial period of acclimation. On day 4, 0.50 g of powdered spinach was added to the daily diet preparation as a source of oxalate, which equates to 52umol oxalate, the rat-equivalent of average human consumption of oxalate. Between study days 4-13, the concentration of calcium, dietary fat, and spinach levels were gradually titrated in a uniform manner across all animals to induce hyperoxaluria in the RYGB-treated animals before finalizing the SH-disease inducing dietary conditions. The final SH dietary conditions on a daily, per cage basis was 30 g high-fat chow, 1.5 mmol $CaCl_2$, 0.833 g spinach, 0.06 g xanthan gum.

Preclinical *in vivo* rat RYGB HO study design for Figure 1C

Two strains were utilized in this study: the control *Bv* strain NB124 containing only the porphyran-utilization modification, and the porphyran-utilizing and oxalate-consuming research strain sWW627.

On study day 14, 24-hr UOx measurements were used to randomized RYGB rats to their experimental groups (*n* = 8 per strain) using an algorithm that randomly sampled possible group assortments and scores the groups to minimize differences in average baseline urine oxalate between groups over all measured days. Also on study day 14, 1.5% porphyran (w/w) was added to the diet of all animals in both sham and RYGB groups. Urine oxalate was sampled daily and on study days 15, 16, 17, and 26 fecal samples were also collected. All 24-hour urine oxalate measurements for each rat collected from study days 20 to 24 were averaged for each animal.

Preclinical urine oxalate assay

Urine was collected from animals daily and frozen at -20°C until quantification. Activated charcoal is first used to purify urine samples. Purified urine samples are then subjected to EnzyChrom's™ Oxalate Assay Kit in a multiplexed format. The kit uses an HRP enzyme, resulting in a color changing reaction where the intensity of the reaction product is directly proportional to the concentration of oxalate in the experimental samples and can be compared to oxalate standards to quantify the concentration. Six dilutions of each urine sample with and without a known oxalate spike-in (to calculate inhibition) is performed, and all dilutions in the linear range and with minimal inhibition are extrapolated based on dilution and a standard curve and averaged to obtain a sample's urine oxalate concentration.

Preclinical fecal CFU strain quantification

All research strains incorporated antibiotic resistance markers to enable enumeration of colony forming units in fecal samples. Using a 1 µL inoculation loop, 1 µL of feces was resuspended in 200 µL of PBS and serially diluted 7 times. 3 µL of each diluted sample was plated onto BHIS agar plates with the appropriate selective antibiotics and incubated anaerobically at 37°C. 24 to 48 hours after plating, number of countable colonies as well as uncountable lawns (>30 colonies per spot) and empty spots were recorded and used to calculate the most probably CFU for each dilution series.

Preclinical fecal qPCR strain quantification

Fecal samples are weighed and mixed with a known amount of a spike-in control strain that contains a genomically integrated GFP sequence, which is used as a normalization control for quantifying absolute cell concentrations. Samples are then subjected to physical lysis by bead-beating before total DNA is extracted using the Zymobiomics DNA Miniprep Kit. Bacterial strain density was enumerated from total DNA by qPCR using primers (ACGGCTGTCCATTCAAAGAGC and TTGTTCCGTCAGTTTCGCGTC) to amplify a region of the porphyran PUL. The GFP gene (introduced in the spike-in extraction control) was also amplified by qPCR with primers (ACGAGCGTCTGATTAACCCG and ACCGTTGACATCACCATCCAG) and used to normalize Ct values across experimental samples for a given extraction batch.

Preclinical *in vivo* mouse strain dosing

Strain banks were prepared by inoculating BHIS with a picked colony or dilution limited culture and grown anaerobically at 37˚C to an $OD_{595nm}$ of 0.6-1.0. Cultures were diluted 1:100 into pre-reduced BHIS and grown anaerobically at 37˚C to an $OD_{595nm}$ of 0.8 to 1.0, then the culture was spun down and resuspended in PBS with 10% glycerol and stored at -80˚C. Immediately before dosing to mice, the strain bank is thawed, mixed 1:1 with 10% w/v sodium bicarbonate solution (pH = 8.5) and each mouse was gavaged with 200 µL of the strain bank bicarbonate mix.

Preclinical *in vivo* mouse handling

Studies were conducted at a CRO vivarium, Charles River Accelerator Development Labs (CRADL), located in South San Francisco, CA in strict accordance with a protocol for Care and Use of Laboratory Animals approved by the CRADL IACUC. In all studies C57BL/6 mice from Taconic or Charles River Laboratories (CRL) were housed in Innovive Disposable IVC Rodent

Caging System and administered a standard rodent chow (TD.2920x, Teklad diet) with *ad libitum* water during an initial one-week acclimation and observation period.

<u>Preclinical *in vivo* humanized 1x biocontainment study design for Figure 2</u>

For the humanized mouse experiment from Figure 2, germ-free mice C57BL/6 from Taconic were handled aseptically and housed with HEPA air filtration. Novome partnered with a human specimen procurement CRO, iSpecimen, to gather stool samples from healthy volunteers under Institutional Review Board (IRB) approval. Stool samples were flash frozen and resuspended in anaerobic phosphate-buffered saline (PBS) for oral gavage into germ-free animals housed under gnotobiotic conditions at a partner research animal CRO. Five donor stool samples were chosen to independently generate distinct humanized mouse colonies— each representing a single donor community. One of the five samples was found to contain a porphyran-utilizing strain and was excluded. Following a 3-week stabilization period after stool gavage, two humanized mice from each donor group were dosed with Novome strains representing two treatment conditions: a porphyran-consuming control strain (NB144) or the therapeutic strain NB1000S.

Porphyran was supplied in the diet 1 week before Novome strain treatment and maintained for 4 weeks after the introduction of NB144 or NB1000S. During this period, strain and transgene abundance in feces was quantified by qPCR. Shed strains were clonally isolated on Day 29, one day before removal of porphyran from the diet, and subjected to *in vitro* assays to assess maintenance of the key NB1000S phenotypes. Four weeks after addition of NB144 or NB1000S, porphyran was removed from the diet, and strain abundance in shed feces was monitored by qPCR of transgenes for an additional 4 weeks. NB1000S was isolated from feces from two animals demonstrating persistent colonization at the study termination and subjected to whole genome sequencing.

<u>Preclinical *in vivo* humanized 3x biocontainment study design for Figure S12B</u>

Human stool samples were obtained from the NOV-001-CL01 clinical trial corresponding to subjects 102-151, 102-154 and 102-158. On study day 0, germ-free C57BL/6 mice from Taconic randomized into groups of 3 mice, gavaged with stool associated with the group and housed together followed by a 4-week stabilization period. On study day 28 mice were switched to chow containing 0.7% porphyran and feces was sampled every ~3 days for 19 days. On study day 47 mice were switched to a diet lacking porphyran, were individually housed and continued to have feces sampled every ~3 days for 14 days. Strain abundance was quantified with qPCR for the porphyran PUL.

<u>Preclinical *in vivo* conventional 3x biocontainment study design for Figure 4E</u>

On study day 0, specific pathogen free (SPF) 8-week old C57BL/6 mice from CRL were randomized into two groups of 6 mice, switched to chow containing 0.7% porphyran and gavage with a 1x or 3x biocontained strain. On study day 7, mice were switched to a diet lacking porphyran and were singly housed. Cage bedding was changed on study days 7, 12 and 21. Throughout the study mice, feces was sampled every 2 or 3 days and strain abundance was quantified by qPCR of the porphyran PUL.

<u>Preclinical *in vivo* biocontained alternative species study design for Figure 12C</u>

On study day 0, specific pathogen free (SPF) 8-week old C57BL/6 mice from CRL were randomized into eight groups of 6 mice (for the 3x biocontained strain) or 4 mice (for all other strains), switched to chow containing 0.7% porphyran and gavage with a 1x or 3x biocontained strain. On study day 7, mice were switched to a diet lacking porphyran and were singly housed. Cage bedding was changed on study days 7, 12. Throughout the study feces was sampled and strain abundance was quantified by qPCR of the porphyran PUL.

Preclinical *in vivo* long-term colonization study design for Figure S4

On study day 0, 15 specific pathogen free (SPF) C57BL/6 mice from CRL were all gavaged with NB1000S and switched to chow containing 0.7% porphyran. On day 21, 6 mice were euthanized and cecal contents were collected and isolates were phenotyped as described above in "Preclinical *in vitro* strain high throughput characterization". For the remainder of the study mice were euthanized when they met the humane endpoint criteria and were otherwise kept on porphyran containing chow and sampled regularly. Samples taken on day 421 were also used to isolate and phenotype strains.

NB1000S clinical material preparation

Upon completion of all genetic engineering steps, a research cell bank (RCB) of NB1000S (also referred to as sZR0323) was generated at Novome via re-streak and isolation of a single bacterial colony on solid media plates and outgrowth in liquid media prior to cryopreservation. The RCB was whole-genome sequenced and phenotypically tested for biocontainment and oxalate consumption as indicated above. Additional PCR, aerobic culturing, and petrifilm tests were done for adventitious organisms (contaminants). The RCB was then sent to Arranta Bio (now Recipharm) (Watertown, MA) for expansion into a master cell bank (MCB). Cells were expanded as described previously with a few modifications. The culture media used, CMB001, was identical to BHIS with three changes: Vegitone-derived BHI (VegBHI MV10 from HiMedia) was substituted for BHI, a non-porcine hemin from Binbo Biological was used, and sodium hydroxide or sulfuric acid was used to neutralize pH to 7.0. Additionally, cell expansion was a two-stage passaging of the cells – first, a 1:100 dilution into 50 mL of media, then, once OD600 reaches 1.0-1.4, another 1:100 dilution into 5 x 1 L cultures. These were incubated again until OD600 reached 1.0-1.4, and cells were harvested via refrigerated 4C centrifugation at 16k RCF. A final resuspension of 4L of culture into a 570 mL solution of 80% (v/v) fresh media and 20% (v/v) glycerol cryopreservative yielded hundreds of tubes of 7-fold concentrated cells. These were frozen at -80C.

Manufacture of the NB1000S drug product from MCB also took place at Arranta Bio. One vial of the master cell bank (MCB) was taken from the -80°C freezer to thaw at room temperature for 15 minutes. Inside the COY anaerobic chamber set at 85%N2/10%CO2/5%H2, six 1L bottles were inoculated with 250 µL of MCB into 1 L CMB001 medium. These bottles were sealed and incubated at 37°C, 125 RPM in a shaking incubator outside of the COY. After approximately 12 hours, the culture was sampled until the OD600 ≥ 1.0. At that point, the culture was harvested by selecting the five 1 L bottles with the lowest final OD600 ≥ 1.0. Cultures were harvested aerobically by centrifugation at 4700rcf for 10 minutes at 4°C. The supernatant was discarded, and the cell pellet is resuspended in a 1:1 volume of phosphate buffered saline (PBS) to wash the cells. The washed cells were centrifuged again at 4700rcf for 10 minutes at 4°C. The supernatant was discarded, and the cell pellet is resuspended in a 1:2 volume of PBS/10% glycerol. The resuspended cell slurry was aerobically dispensed using a Flexicon PF6 semiautomated liquid filler, at 1.1 mL volume into each of 2000 or more cryovials (2.0 mL). The dispensed vials were frozen at or below -70°C and sent to Catalent for labeling and distribution.

NB2000P clinical material preparation

NB2000P, or food-grade dried porphyran extract, was produced from *Porphyra spp.* seaweed in a multi-stage, multi-facility process. Briefly, dried raw nori seaweed from China was hot water extracted in the presence of hydrogen peroxide and ascorbic acid. Solids, proteins, and simple sugars were removed, leaving behind polysaccharide. This polysaccharide was then freeze-dried, milled, sterilized, and packaged.

The hot water extraction took place at a single facility, Revela Foods / North Star Processing (Litchfield, MN). Per batch, 1890 L of preheated 80C water was mixed with 1.5 kg of ascorbic acid (Spectrum Chemical) and 200 kg of dry seaweed (Nantong Haida Aquatic Food Co, Jiangsu Province, China). The mixture was blended, and another 910 L of hot water was added. The mixture was held at 80 C and continuously blended while $H2O2$ was added (ultimately 10L of 12% in 2L increments) at 15-minute intervals to achieve mild free radical hydrolysis of the seaweed polysaccharides to reduce the viscosity and prevent gelling. After about 2.5 hours from seaweed addition, the $H2O2$ was undetectable. The hydrolyzed extracted seaweed slurry was passed through a decanting centrifuge running at 2100 RCF @ 3250 RPM (Alfa-Laval Sharples P-3400 sanitary centrifuge decanter) to separate out most of the bulk sedimenting solids. The feed rate was 30 L / min, so processing took about 90 minutes. Sedimented material was discarded as solid waste. Another 1060L of hot water was added back to top up the mixture and keep solute concentration low. The diluted decanted solution was further clarified through a second centrifuge running at 10000 RCF (Westfalia SA 14 sanitary clarifying centrifuge). At a feed rate of 10-15 L/min, this step took about 4.5 hours. Solids were again discarded.

Finally, the fully clarified liquid was cooled to 55C, combined with a second batch of the same size (for a total of 400 kg nori processed) and cycled through an ultrafiltration (UF) system featuring two 30kDa MWCO 8038-sized filter units (Synder Filtration MK-3B-8038), and freshwater was added as permeate was discarded. Retentate was re-diluted and passed through the clarifying centrifuge again, and pH was adjusted to 7.2 by addition of NaOH. With ongoing UF, 10000L of water was added stepwise in 1000L units. A final finishing UF step concentrated the retentate down to 10% w/v solute concentration, and heat treatment at 93C for 30 minutes prepared the liquid for transport to the drying facility. A yield of 7.8%, or 320 L of 10% porphyran, from 400 kg nori was obtained.

The 10% porphyran solution was transported to Oregon Freeze Dry (Albany, OR) for lyophilization and milling. Product was pumped into trays at 7L / tray and frozen at -29C. Trays were then lyophilized over 24 hours, and dry product was first broken by hand and then milled through a #12 screen. Finally, the dried porphyran was vacuum packed into polyethylene bags.

Dry porphyran was then shipped to National Food Lab (Naples, FL) to be packed into 0.5, 2.5, and 10 g sachets. These sachets were then shipped to Steri-Tek (Fremont, CA) for electron-beam sterilization. The finished product was sent to Catalent for labeling and distribution and Eurofins SF-Analytical (New Berlin, WI) for bioburden, heavy metal, and other safety testing.

NOV-001 Clinical Trial Design Summary

Details of the study design can found in the Clinical Protocol  and in the NOV-001-CL01 Clinical Study Report posted at https://www.clinicaltrials.gov/study/NCT04909723, and are described briefly below. The study was conducted according to ICH Good Clinical Practice guidance under US Investigational New Drug (IND) approval 026990 and under Health Canada approval e264188, and with ethical approval from the Institutional Review Board of Advarra.

Study NOV-001-CL01 was a prospective, multicenter, randomized, controlled, safety, tolerability, PD, and early efficacy study of NOV-001 in adult healthy volunteers (HV) and patients with EH secondary to RYGB or BPD-DS bariatric surgical procedures. The study design, actual enrollment, treatment, and follow-up durations utilized in Stage 1 are shown schematically in figure S5, figure S6 and figure S8 below, and the schedule of assessments in Stage 1 is shown in table S3.

In Stage 1, 39 healthy volunteers were enrolled in 7 groups to assess initial safety, tolerability, and PD at varying doses of NOV-001 or of its components, NB1000S and NB2000P. Subjects were randomized into Groups A and B and were assigned to Groups 1, 2, 3, 2X, and C. Groups A, B, and C comprised control groups, where placebo, NB1000S alone, or NB2000P alone, respectively, were administered. The treatment randomization/assignment was single-blinded.

Stage 1 subjects underwent screening (up to 35 days) and if eligible were randomized or assigned, then treated with NOV-001 or placebo for 14 days. The duration of intended follow up was variable depending on group assignment and, in subjects treated with NB1000S, on any persistent fecal shedding of NB1000S.

An internal Safety and Engraftment Assessment Committee (SEAC) was established to review emerging safety and engraftment data and to support decision making for NOV-001 dose changes and escalations. In addition, for both stages of the study, individual and study-wide Stopping Rules were in place to help safeguard the safety of the subjects.

A porphyran rechallenge portion of the study was conducted on a subset of Stage 1 subjects in which groups of HVs were re-randomized to be treated with either NB2000P or dietary porphyran after completion of their initial treatment period (Fig. S6). NB1000S engraftment was assessed following treatment with NB2000P and dietary porphyran.

In Stage 2 of the study approximately 20 evaluable patients with EH were to be randomized in a single-blinded manner in a 3:1 ratio to either NOV-001 (~15) or placebo (~5). Patients underwent screening (up to 35 days), and if eligible were randomized and treated with NOV-001 or placebo for 28 days. The study design and actual enrolment are shown schematically in figure S7, below.

Patients were to be followed for a minimum of 56 days and maximum of 168 days after treatment cessation. Stage 2 patients with persistent shedding at 84 days after NOV-001 treatment cessation (Study Day 112), or early termination, were to be treated with an antibiotic, at approximately 112 days after NOV-001 treatment cessation (Study Day 140) (Fig. S8).

Stage 2 enrollment was terminated following the randomization of 12 patients and a review of emerging engraftment and efficacy data. The schedule of assessments in Stage 2 is shown in table S4.

NOV-001 Clinical Trial Strain Quantification

The bacteria DNA was extracted and purified using the E.Z.N.A Universal pathogen kit (Omega Bio-tek). The purified DNA was analyzed by quantitative PCR (qPCR) using Applied Biosystems custom designed TaqMan™ Multiplex Probes Assay (ThermoFisher Scientific).

In order to quantify NB1000S, fecal samples were weighed out and mixed with a known concentration aliquot of ZymoBIOMICS Spike-in Control I (High Microbial Load). ZymoBIOMICS Spike-in Control I consists of equal cell numbers ($2 \times 10^7$ per 20µl) of two bacteria strains alien to the human microbiome, *Imtechella halotolerans* and *Allobacillus halotolerans*. When spiked into an unknown sample, this product served as an *in situ* positive control to ensure each sample can be quantified accurately. As our target bacteria strain is gram-negative, *Imtechella halotolerans* is chosen for the referral calculation and the 16S gene copy number per cell/genome of *Imtechella halotolerans* is 3.

Samples with unknown concentration of NB1000S was amplified and cycle number of NB1000S construct was compared against spiking-in *Imtechella halotolerans* bacteria cycle number to determine concentration. Before strain quantification calculation, the raw Cp value was subtracted from 40 (the max possible Cp) to normalize the Cp value. Then the normalized Cp values was used in the following equation for the strain quantification:

Log10NB1000S copies/gram = Log10 (((2^(Cp_norm_unknown - Cp_ZymoBIOMICS_control)) × 6 × $10^7$) / fecal_sample_weight_g)

For NB1000S quantification qPCR oligo sequences were GCCTTTACGCCGGATATTA for forward, GATGCCAGGCAGGTTAG for reverse and TTGCAGGAGCTGGGTTATAGCGAC with 5’ 6-FAM and 3’ BHQ-1. For spike-in quantification qPCR oligo sequences were AATGGTCGGAAGACTGA for forward, CATGGTACCGTCATCAAC for reverse and TGTATGGGAAGAATAAGGCCTACGTGT with 5’ 6-FAM and 3’ BHQ-1.

**Supplementary Text**

Engineering oxalate degradation

As no natural Bacteroides are known to consume oxalate, we took the approach of engineering a synthetic oxalate breakdown pathway, based largely on the oxalate breakown pathway of *Oxalobacter formigenes* (*Of*) but utilizing relevant genes from multiple organisms. 92 total genes including multiple homologs of OXS, ACOCT, FRC, OXC and formate:antiporters as well as other activities related to oxalate degradation were tested in various combinations for their ability to increase oxalate degradation. An initial strain containing a OXC (*Saccharomyces cerevisiae* YBR222C), OXS (*Sc* YEL020C) and oxlT (*Of* oxlT) yielded detectable oxalate removal.

Next, we iteratively add additional genes to test for improved activity. To simplify the screening, each gene was expressed as a single gene operon with the P_BfP1E6 promoter and a ribosome binding site (RBS) and leader peptide (LP18). RBSs are highly context dependent, and this approach maintains the local context to increase chances of high expression. Screening for increased rates yielded our first-generation strain, sWW152, that in addition to the genes above contained another OXS (*Arabidopsis thaliana* AAE3), another OCT (*Of* oxc), an ACOCT (*E. coli* yfdE) and an FRC (*Ec* frc). Adding a second copy of the first-generation oxalate degradation pathway, making sWW627, further increased the oxalate consumption rate. The resulting strain, however, had many repeats of expression elements and a duplicated pathway that could lead to genomic instability.

To improve genomic stability, we redesigned the pathway to a single operon design that does not repeat any regulatory elements. For each of the 7 genes in our first generation strain we created RBS library driving each gene with a C-terminal nanoluciferse fusion to quantify expression. High expression was achieved for most genes, including the transporter oxlT. Both OXC genes and one of the two OXS genes (*At* AAE3) had top hits about 1 log lower than the other genes (S1A). Selected RBSs from the library were used to create single-operon designs (S1B). The potential design space was sparsely sampled with 96 designs shuffling the RBSs and operon order, primarily focused on high gene expression, but included low expression as well, particularly for oxlT. Pathway designs were genomically integrated between two convergent genes, BVU_1237 and BVU_1238, with flanking homology arranged such that the vector, including antibiotic resistance cassette, can be removed, leaving only the oxalate degradation pathway. These oxalate pathways were integrated into a Bacteroides vulgatus (*Bv*) strain (ATCC 8482) containing a porphyran PUL, and oxalate degradation rates were assayed. Of the strains that demonstrated detectable oxalate degradation, eleven single-operon hits were advanced to antibiotic marker removal and further comparisons of fitness and activity.

Next, we assessed the activity and fitness of these 11 strains. To decouple oxalate degradation rates from fitness, oxalate consumption was measured for cultures that were concentrated to $10^{10}$ CFU/ml, approximately the level we typically reach in the gut, and oxalate reduction was measured over 10-80 minutes. For measuring fitness, cells were diluted 1:50 into rich media with or without oxalate 12mM. In the absence of oxalate, none of the cells show a significant growth defect, even the strains with the highest consumption rates of more than 15 mM oxalate per hour (Fig. S1C, open circles). Comparing oxalate consumption to doubling time in the presence of oxalate (Fig. S1B, closed circles), we see that our highest oxalate consumers are among the least fit in the presence of oxalate with a doubling time of about 7 hours, while lower producers have a faster doubling, with some close to the about 2.3 hour doubling time of the control strain lacking the oxalate pathway, NB144. Despite the general trend of highest oxalate consumption correlating with longer doubling times in 12 mM oxalate ($R^2$=0.61), some strains showed similar oxalate degradation rates but very different doubling times, such as sZR0310 and sZR308, both degrading around 9 mM per hour but having doubling times of 3.5 hours and 7.3 hours, respectively. No clear trend emerged between any gene's expected expression level (from Fig S1B) and the oxalate degradation or doubling times, though this may be due to the potentially large changes in RBS strength expected from changing the genetic context of the RBSs. None of the oxalate consuming strains matched the relatively fast 2.3 hour doubling time in oxalate of the control, but many strains clustered around a 3.2 to 3.6 hour doubling time, despite 10-fold differences between oxalate degradation rates. Among those strains with only modest growth defects, sZR0310 was the highest oxalate consumer, and thus that oxalate degradation pathway was selected for advancement into the clinical strain.

In addition to sZR0310 having a 40% increased oxalate degradation rate, compared to the best first-generation strain, sWW627, the refactored pathway is also significantly smaller, down from 2 copies of a 13 kilobase pathway to one copy of a <8 kilobase pathway (Fig. S1E). The two activities, OXS and OXC, that were originally performed by two genes are now performed by a single gene, despite the extra gene helping in the original context. Also, the sZR0310 pathway contains far fewer repeated expression parts, which we expect to improve genetic stability by reducing loss of the pathway components from homologous recombination.

Growth curves of sZR0310 in rich media without oxalate showed that its growth rate is not significantly different that the equivalent control strain without the oxalate pathway (Fig. S1D). In the presence of 12 mM oxalate, the growth rate of the control strain is slightly reduced while the growth rate of sZR0310 is substantially reduced and reaches a lower final OD. *Oxalobacter formigenes* makes used an oxalate to formate degradation pathway that is essential for it's growth by taking advantage of the cytoplasmic depletion of hydrogen ions to create an ATP generating gradient. In theory, it may have been possible to modify our Bacteroides strain to benefit from the breakdown of oxalate. However, that effort likely would require substantial effort, including tuning metabolism beyond our pathway, so we moved forward with the sZR0310 pathway, as it was among the lowest fitness defects in the refactored pathway set (Fig. S1C).

Dietary enteric hyperoxaluria model

Rats have been used as an animal model to study enteric hyperoxaluria (*6, 7*). To induce EH in Sprague-Dawley rats spinach (High in Oxalate) was added to a Low-Calcium chow (HOLC diet) to increase dietary oxalate absorption. By individually housing animals in metabolic cages, food intake, bacterial abundance, and 24-hour UOx excretion levels were quantified.

To determine the ability of our strain to reduce urine oxalate, we ran this EH model comparing three groups of rats (Fig. 1C): no induction or strain control (grey), induced and colonized with the porphyran-utilizing control strain NB124 (purple), or induced and colonized with the porphyran-utilizing and oxalate degrading therapeutic strain sWW627 (blue). After three days of baseline urinary oxalate (UOx) measurements, animals in induced groups were transferred to the HOLC diet to induce hyperoxaluria. After four days of disease induction, induced animals were split into the two treatment groups, each rat received a single dose of ~$10^9$ CFU by oral gavage administered on Day 7 and rats were placed on a diet containing 1.7% porphyran. Fecal bacterial abundance for colonized animals and 24-hour UOx for all animals were quantified throughout the experiment.

24-hour UOx plotted over time (Fig. 1B) shows an about five-fold increase in urine oxalate in response to the HOLC diet. Two days after strain administration, the 24-hour UOx levels begin to diverge for the induced groups treated with control or therapeutic strains. Averaging the 24-hour UOx over the final 3 days of the experiment for each rat gives a 47% drop in average UOx ($p < 0.001$) when comparing the induced groups given the control or the therapeutic strains.

To understand how porphyran dose and associated colonization level relate to urine oxalate reduction, we carried out a similar EH model varying porphyran dose (Fig. S2A). After disease induction, animals were split into three groups: no treatment control (no strain, no porphyran), colonized with the porphyran-utilizing control strain NB124 (purple), or colonized with the porphyran-utilizing and oxalate degrading therapeutic strain sWW627 (blue). Treatment groups each received a single dose of ~$10^9$ CFU by oral gavage administered on Day 7. For the remainder of the study, the two strain treatment groups were exposed to an increasing concentration of porphyran in the diet: starting with 0% dry weight (Days 8-10), 0.05% (Days 11-13), 0.17% (Days 14-16), 0.5% (Days 17-19) and culminating at the highest dose of 1.5% (Days 20-22).

As expected, the HOLC diet rapidly and uniformly induced elevated UOx levels (Fig. S2B, comparing Days 1-2 to Days 3-7). Following strain treatment on Day 7, the group mean UOx levels remained unchanged in the absence of porphyran to support bacterial colonization (Fig. S2B, comparing Days 3-7 before strain dosage, to Days 8-11 with strain present but 0% dietary porphyran). However, the oxalate-degrading research strain sWW627 resulted in a significant -24% reduction in UOx as compared to the untreated control at a low concentration of dietary porphyran tested (Fig. S2B, 0.17% w/w porphyran, Days 15-17, $p = 0.018$). As the dose of porphyran gradually increased over time, so did the efficacy of sWW627 for reducing UOx levels: scaling well over the tested dose range, and peaking at 41% reduction achieved during 1.5% porphyran treatment (Fig. S2B, Days 21-23, $p = 0.013$). The hyperoxaluria-induced but non-treated control group not receiving any strain or porphyran demonstrated fluctuating UOx levels over the study duration (Fig. S2B, grey bars), illustrative of variability in UOx over time observed in the treatment groups. Fecal strain enumeration by plating revealed the predictable strong correlation between porphyran dosage and bacterial abundance for both strains (Fig S2C). A reduced sWW627 colonization at low porphyran doses as compared to the control strain was noted, suggestive of an *in vivo* fitness defect conferred by oxalate-degradation that is no longer apparent at the highest dosage of porphyran (Fig. S2C, comparing Days 8-11, $p = 0.004$ to Days 21-24, $p = .846$).

<u>Gastric Bypass enteric hyperoxaluria model</u>

A second rat model intended to better mimic the underlying disease etiology of enteric SH was implemented. It has been observed that disruption of normal gastrointestinal (GI) physiology through IBD or gastric bypass surgery predisposes patients to SH (*8, 9*). In the case of gastric bypass surgery, the proposed link between surgical intervention and subsequent enteric SH is via fat malabsorption in the GI tract (*10*). When GI tissue necessary for fat absorption is surgically removed, the resulting increased levels of unabsorbed fat in the GI tract competes with free oxalate to bind with dietary calcium. It is hypothesized that when more fat is available to interact with calcium, levels of unbound, easily absorbable oxalate in the colon increases, thus leading to enhanced colonic uptake and urinary excretion of oxalate. As observed in humans, rats subjected to Roux-en-Y Gastric Bypass surgery (RYGB) develop secondary hyperoxaluria (*11, 12*).

To determine whether the oxalate-consuming research strain sWW627 can reduce UOx excretion in a model of gastric bypass surgery, Sprague-Dawley rats were subjected to RYGB surgery or a sham surgical procedure. Porphyran at 1.5% w/w was added to the diet for all animals, and RYGB animals were administered a single dose of $10^9$ CFU of either the *Bv* control strain containing only the porphyran modification (NB124, purple) or the oxalate-consuming sWW627 strain (blue). In groups that receive the control strain, elevated UOx was observed in the bypass surgery as compared to sham group (Fig 1C). Animals colonized with sWW627 exhibited a lower mean UOx than animals colonized with the control strain NB124. The difference in UOx was statistically significant in the final five days of the experiment corresponding to a roughly 40% decrease in oxalate for the sWW627 group, representing complete elimination of the increase in UOx due to RYGB surgery.

Engineering porphyran dependent biocontainment

To biocontain our therapeutic strain so that when porphyran is not present our cannot persist on alternative polysaccharides, we considered several approaches. We considered attempting to limit the ability of our strain to utilize alternative polysaccharides. However, removing the large repertoire of polysaccharide utilization genes estimated to make up almost 20% of the genome of typical strains (*13*) would be a substantial effort, and our efforts to target key pathways or regulators of carbohydrate metabolism suggested this approach may be difficult without imposing significant fitness defects or large engineering efforts. Approaches to express toxins or otherwise require active killing were avoided as common loss of function mutations to disable the pathway would be difficult to combat. Also, ensuring exposure of our strain to any active "kill" signal would be challenging as even a single cell located in a difficult to reach site like a biofilm or intestinal crypt that is not adequately exposed to the signal could repopulate the gut later. Indeed, we see that active killing of our strain using antibiotics that our strain is sensitive to frequently had little impact on the colonization level of our strain in human subjects (Fig. S10, discussed more below). Another common approach is knocking out thymidylate synthetase (thyA), an essential gene that can be complimented by providing thymidine when culturing the strain. The thyA knockout approach has been successfully used to attenuate therapeutic bacteria (*14, 15*). However, we are taking the unique approach of allowing our strain to engraft and expand in the gut, so it is not clear that a molecule widely utilized by organisms can be reliably delivered to the gut at non-limiting concentrations.

The approach we took used a porphyran responsive hybrid two-component system (HTCS), one of which exists naturally in the porphyran PUL, to drive expression of an essential gene, an aminoacyl-tRNA synthetase in our case, that is difficult to complement. One advantage of this approach is the simplicity of not introducing a second molecule needed for biocontainment. Identifying a second biocontainment molecule that is safe for humans, not absorbed or modified by the host or microbiota and can be sense by the target strain is challenging and would also introduce manufacturing, regulatory and clinical complexities. Another advantage of this approach is that common loss of function mutations would not break the biocontainment. Finally, with a modest engineering effort it is possible to implement this strategy without imposing any detectable fitness defect in the presence of porphyran.

To find a promoter capable of driving porphyran-dependent expression of an essential gene, we tested promoters in the porphyran PUL that we obtained from a sewage isolate that has a high homology to the originally characterized porphyran PUL (*3, 16*). Guided by previous expression data (*16*), we selected multiple intergenic regions from the porphyran PUL to drive luciferase expression. Constructs with each of these promoters driving luciferase expression were added to a bacteroides containing the porphyran PUL and luminescence was measured after growth in rich media, with or without porphyran (Fig S2A). The most highly induced promoters we cloned are shown, along with the constitutive promoter that drives the HTCS (BACPLE_01699) and constitutive phage promoters. The promoter in front of the BACPLE_1680 homologous gene, here termed P_por10, showed a strong induction and was selected for biocontainment of an essential gene.

Next, we sought to replace the native regulation of an essential gene with P_por10, replacing the native promoter and simultaneously tuning the RBS so that when induced the essential gene is near optimal levels, but when uninduced the essential gene levels are dropped low enough that the cells are unable to grow. We developed erythromycin selectable and 4CP counter-selectable vectors, containing P_por10 followed by an RBS library, and arranged the homology such that a loop-out on counter-selection could result in the native machinery being replaced with P_por10 and a member of the RBS library driving expression of an essential gene (Fig S2B). For each essential gene target, after counterselection many colonies were picked and screened for growth in rich media with porphyran and no growth without porphyran, which yielded biocontained strains (Fig. S2C) for the thyA (for proof-of-concept; BVU_1950), argS (BVU_1081), cysS (BVU_1009), lytB (BVU_1937) and RF-2 (BVU_1485). Of the strains that showed no detectable growth defect in the presence of porphyran, we tested the ability to outgrow in the presence of absence of porphyran. The strain with argS-dependent biocontainment showed the most growth restriction in the presence of porphyran and was selected for use in our clinical strain. Incorporating the argS biocontainment into the sZR0310, which already contained the porphyran PUL and oxalate degradation pathway, created the NB1000S strain that was advanced into the clinic (Fig. 2B).

Biocontainment in humanized mice

To understand how NB1000S may behave in human subjects, we investigated colonization dynamics of the germ-free mice that were colonized with stool from one of 4 health volunteers. Donor exclusion criteria included the use of antibiotics, antivirals, antifungals or immunosuppressants within the last 6 months; known GI disorders such as IBD, irritable bowel syndrome (IBS), Celiac disease, or Crohn's disease; atypical stool morphology as judged by the Bristol Stool Scale; human immunodeficiency virus (HIV) or Hepatitis B or C positive donors; and visibly bloody stool. Stool samples were flash frozen and resuspended in anaerobic phosphate-buffered saline (PBS) for oral gavage into germ-free animals at a partner research animal CRO, housed under gnotobiotic conditions.

To explore a diverse set of human gut microbiota communities and enhance the ability to extrapolate from nonclinical results, four donor stool samples were chosen for this study. Donor samples were selected to represent a breadth of demographic characteristics such as age, sex, body mass index (BMI), and race. The stool samples were used to independently generate distinct humanized mouse colonies— each representing a single donor community. Following a 3 week stabilization period after stool gavage, two humanized mice from each donor group were dosed with Novome strains representing two treatment conditions: a porphyran-consuming control strain (NB144) or NB1000S. porphyran was supplied in the diet before Novome strain treatment and maintained for 4 weeks. During this period, strain and transgene abundance in feces was quantified by qPCR. Shed strains were clonally isolated on Day 29, one day before removal of porphyran from the diet, and subjected to *in vitro* assays to assess maintenance of the key NB1000S phenotypes. After one month, porphyran was removed from the diet, and strain and transgene abundance in shed feces was monitored by qPCR for an additional 4 weeks.

As expected, supplementing porphyran in the diet facilitated rapid and robust engraftment of porphyran-consuming strains in both strain treatment conditions, across all of the humanized mouse donor groups (Fig. 2C). Interestingly, a fifth donor microbiota animal group tested in this study was not permissive for Novome strain colonization due to the pre-existing presence of a native porphyran-utilizing strain (confirmed by porphyran PUL locus detected by qPCR at Day 0, prior to Novome strain treatment: data not shown). An analysis of global metagenomics data suggests that only ~2% of adults in Western society harbor a porphyran-utilizing strain in their native microbiota, as compared to ~30% in Asian populations (*17*).

Removal of porphyran resulted in a dramatic and rapid reduction in NB1000S fecal abundance across all of the microbiota communities tested, dropping the NB1000S cells/g feces on average ~6 log to below the LOD within 2 weeks in 5 of 8 animals tested (Fig. 2C). In contrast, the porphyran-consuming, non-attenuated control strain NB144 exhibited a modest and variable drop in strain abundance across the humanized communities following porphyran removal.

In view of the high NB1000S starting abundance (~$10^{10}$ genome copies/g of feces) it is notable that the porphyran conditional attenuation strategy robustly reduced NB1000S shedding to below the limit of detection in 5 animals representing 3 microbiotas within 2 weeks. In donor D (red) both of the NB1000S colonized animals initially dropped in abundance after removal of porphyran; however, NB1000S abundance subsequently increased in abundance to ~ $10^8$. Similarly, in Donor A (blue) both NB1000S colonized animals exhibited a rapid drop in abundance to near the LOD; however, 1 week after porphyran removal subsequently only one of the Donor A animals re-emerged, and exhibited a ~ 2 log increase in strain density by the end of the study.

Comparison of the relative abundance of copies of the porphyran PUL vs. copies of OxlT amplicons within a given NB1000S colonized animal showed these introduced transgenes are highly correlated ($R^2$=0.99). This observation provides evidence against horizontal gene transfer of either set of transgenes to other microbial species within NB1000S treated animals. To further characterize shed NB1000S in the humanized mouse model, Novome also isolated shed clonal isolates of NB1000S in the feces at the end of Phase 1 of the study, one day before porphyran was removed. Because NB1000S is devoid of any antibiotic selection markers, culturing shed clonal isolates from complex feces was technically challenging and precluded a widespread analysis across the study duration; however, Novome was able to isolate 20 distinct clones from 7 of 8 animals representing all 4 donor microbiotas at Day 29. Phenotypic analysis of porphyran utilization, oxalate consumption, and porphyran conditional attenuation were confirmed using the *in vitro* phenotypic assay. All 20 of the shed NB1000S isolates maintained their key engineered phenotypic properties, consistent with the robust NB1000S phenotypic stability.

At the conclusion of the study, Novome utilized an extensive outgrowth procedure necessary to enrich for low-abundance NB1000S isolates from fecal pellets collected on the final day of Phase 2 and succeeded in obtaining isolates from 2 of the 3 animals in which strain persistence was observed (Fig. 2C). Phenotypic analysis of these isolates suggested they are no longer fully conditionally attenuated, as demonstrated by moderate growth in the absence of porphyran.

Overall, these data confirm that the porphyran conditional strain attenuation strategy succeeds at significantly reducing fecal shedding of NB1000S upon removal of porphyran, though the biocontainment did not achieve complete clearance of the strain below the limit of detection in every human donor microbiota tested in this humanized mouse experiment.

Additional preclinical characterization of NB1000S

To test genome stability of NB1000S in culture, NB1000S was diluted 1:1000 in rich media and outgrowing 10 successive times. The culture, after those ~100 doubling, was plated and 120 individual colonies were phenotypically assessed. None of the 120 phenotyped isolates had lost porphyran utilization, oxalate degradation or biocontainment. Whole genome sequencing was performed on 20 of the isolates, and the analyses did not reveal any sequence deviations from the NB1000S reference genome at the three examined sites of engineered modifications.

Novome defined the sensitivity of NB1000S to 13 compounds representing the major antimicrobial drug classes. The test panel included antimicrobials deemed most clinically relevant for *Bacteroides* species, as well as antibiotics utilized during strain generation prior to removal of resistance markers. As all antibiotic markers used in construction of NB1000S had been removed, NB1000S was expected to match the *Bacteroides vulgatus* ATCC 8482 type strain it is originally derived from. An antibiogram confirms that like the control type strain, NB1000S is sensitive to al tested antibiotics (Table S2).

Long-term NB1000S colonization in mice

We performed a long-term colonization experiment with the objective of assessing the long-term colonization dynamics and safety of NB1000S in mice, determine if NB1000S retains the ability to degrade oxalate, utilize porphyran and remains biocontained after over a year post colonization. Conventional mice were gavaged once with NB1000S and remained on a diet containing porphyran for the remainder of the study. Throughout the study, and even after over

600 days post-gavage, NB1000S colonization levels remained high, between $10^9$ and $10^{11}$ copies per gram of feces (Fig. S4B, blue). Consistent with all previous studies, animals continued to gain weight after NB1000S colonization and showed no abnormal signs of distress throughout the study.

After 21 or 421 days NB1000S isolates were phenotyped. While all isolates were biocontained, and nearly all could utilize porphyran as the sole carbon source, many isolates were unable to degrade oxalate (Fig. S4A). All isolates from samples collected on day 21 (78 isolates from 2 mice) were able to degrade oxalate, while only 48% of isolates from samples collected on day 421 (122/254 isolates from 6 mice) were able to degrade oxalate. Multiple different frameshift mutations and transposon integration events in the oxalate pathway were observed in a subset of NB1000S isolates. TaqMan qPCR primers and probes designed to target the two of the frequently observed transposon integrations, both in the oxalate pathway promoter region. The abundance of these transposons over the experiment is plotted along with the total NB1000S abundance as estimated by porphyran PUL abundance (Fig. S4B). This oxalate pathway breakage increases gradually over time, appearing to plateau only after several hundred days. Altogether, this indicates there is a clear but modest fitness defect related to oxalate breakdown, but no detectable fitness defect associated with biocontainment in the presence of porphyran.

Engraftment in healthy volunteers

The main pharmacodynamic (PD) endpoint was the level of engraftment of NB1000S in the subjects' GI tracts, as assessed by the number of genomes of the organism present per gram of stool using quantitative polymerase chain reaction (qPCR) assays. The oxalate degrading locus in NB1000S was the target for the primary qPCR assay for engraftment level, and unless otherwise specified this assay is referenced in the engraftment discussions below.

A total of 153 subjects were screened for the study (71 for Stage 1 and 82 for Stage 2), and 119 submitted stool samples and screened for native porphyran-utilizing microbiota. In study NOV-001-CL01 it was found that 11 of 119 screened subjects (9.2%) had porphyranase-positive flora based on the qPCR done at the Screening visit, higher than the expected approximately 2% of the Western population (*17*).

Subjects in the Stage 1 control cohorts Group A (placebo only; N = 3), Group B (NB1000S $10^9$ CFU only, no NB2000P; N = 4) and Group C (NB2000P 10g/day only, no NB1000S; N = 4) did not show the presence of NB1000S in the stool above the qPCR LOD during the treatment or follow-up periods (Fig. S9A), as expected.

In Stage 1 Group 1 (NB2000P 0.5g/day), two of the 8 HV subjects had peak stool NB1000S levels > $10^9$ genomes/g, two had peak levels just below $10^8$ genomes/g, three (including subject 102-116, who had left the CPU early, see above) had peak levels between $10^4$ and $10^7$ on one or two occasions, and one subject did not have NB1000S detectable in the stool above the LOD at any timepoint (Fig. S9A). In this and later groups, the peak level of engraftment was noted between Day 3 and Day 7 of treatment.

In Stage 1 Group 2 (NB2000P 10 g/day), two of the 8 HV subjects had peak stool NB1000S levels above $10^{10}$ genomes/g, three had peak levels between $10^9$ and $10^{10}$, one subject had levels just over the LOD on two occasions, and two did not have NB1000S detectable in the stool above the LOD at any timepoint (Fig. S9B). The time course of engraftment was similar to that observed in Group 1.

Some subjects in these first two Stage 1 HV cohorts did not show evidence of initial engraftment with NB1000S, with either no increase in fecal NB1000S above the qPCR LOD of $10^4$ organisms/g or a transient increase to < $10^7$ genomes per gram of (Fig. S9B), despite pre-treatment administration of Alka Selzer Gold (containing sodium bicarbonate 1050 mg, potassium bicarbonate 344 mg and citric acid 1000 mg), with the intent of raising gastric pH to aid the survival of NB1000S during passage through the upper gastrointestinal tract. For subsequent groups and for Stage 2 the administration of NB1000S was also preceded by the proton pump inhibitor omeprazole ER 40mg, given approximately 90 minutes before the NB1000S. For Group 2X and for Stage 2, a further change was the addition of a second dose of NB1000S $10^9$ CFU, with the first dose of NB1000S to be taken before food and the second dose after food.

With these changes the engraftment was more consistent, with all 8 subjects in Group 3 (NB2000P 2.5 g/day) and all 4 subjects in Group 2X (two doses of NB1000S $10^9$ CFU, NB2000P 20 g/day) showing initial engraftment to levels > $10^8$ genomes / g (Fig. S9C).

In Group 3 (NB2000P 2.5 g/day; NB1000S preceded by omeprazole), two of the 8 HV subjects had peak stool NB1000S levels above $10^{10}$ genomes/g, five had peak levels between $10^9$ and $10^{10}$ genomes/g, and one subject had a peak level between $10^8$ and $10^9$ genomes/g (Fig. S9C). In Group 2X (NB2000P 20 g/day; two doses of NB1000S preceded by omeprazole and *Alka-Seltzer Gold*), three of the four HV subjects had peak stool NB1000S levels above $10^{10}$ genomes/g, and the other had a peak level just below $10^9$ genomes/g (Fig. S9C).

One subject in Group 2X (102-163; represented by the red line in Figure S9C) had a peak level of engraftment of 1.51 x $10^{10}$ genomes/g during the initial part of the treatment period during which they were resident in the clinical pharmacology unit and dosing was monitored by unit staff, but appears to have been markedly non-compliant during the outpatient portion of the dosing period. The number of unopened sachets returned by the subject was indicative of 6 missed doses during this 7-day period, and no stool samples were collected during the outpatient treatment period or after the first post-treatment follow-up visit. This subject was lost to further follow-up.

One subject in Group 2 (102-139; represented by the pink line in Figure S9B) and one subject in Group 3 (102-147; orange line in Figure S9C) showed a decrease in fecal NB1000S level to below the qPCR LOD during the second week of the treatment period, followed by a transient increase to between $10^7$ and $10^8$ genomes/g following treatment cessation. These subjects reported full compliance with the NB2000P regimen throughout the treatment period, and non-compliance was not detected based on the counts of returned sachets of NB2000P. No discrepancies were noted between the fecal qPCR assay based on amplification of the oxalate degradation locus in NB1000S *versus* the qPCR based on amplification of the porphyranase locus in the organism. The reason for these transient reductions in colonization level is unclear.

Amongst subjects who showed engraftment with NB1000S the peak level of engraftment was related to the daily dose of NB2000P, with higher doses associated with higher mean levels of NB1000S as detected by fecal qPCR (Fig. 3C).

In summary, the engraftment data for Stage 1 of the study showed that NB1000S was able to colonize the GI tract to high levels ($10^{10}$ genomes/g of stool, representing 5-10% of the normal total bacterial flora). Colonization was more consistent with administration of two doses of NB1000S, preceded by both omeprazole and antacid. Higher daily doses of NB2000P were associated with higher levels of engraftment with NB1000S.

Together with the emerging safety and tolerability data (Tables S5-6), the engraftment data above was used to determine the regimen to be used for Stage 2 of the study: two doses of NB1000S $10^9$ CFU, one dose preferably before food and a second dose after food approximately 2 hours later; the first dose preceded by omeprazole 40mg ER 90 minutes prior and each dose preceded by two tablets of *Alka Seltzer Gold* 15 minutes prior; and NB2000P 10g/day throughout the treatment period.

Antibiotic treatment of healthy volunteers

The protocol specified that subjects who did not show clearance of NB1000S from the stool after cessation of NB2000P treatment (defined as having a fecal level of > $10^4$ copies/g after day 56 post treatment cessation) were to be treated with a 7-day course of oral antibiotics. Four Stage 1 HV subjects showed persistent fecal shedding of NB1000S, which was not associated with any particular pattern of clinical adverse events. In addition, some subjects with persistent fecal carriage of NB1000S were treated with antibiotics for incidental infections that arose during the follow-up period.

One Stage 1 subject with persistent shedding in Group 1 (Subject 102-115) was treated with amoxicillin/ clavulanate 500/125 mg BID for 7 days, which did not eliminate fecal carriage of NB1000S (Fig. S10).

One Stage 1 subject in Group 3 (Subject 102-151) was treated with amoxicillin/clavulanate 500mg/125 mg BID for 7 days, which did not eliminate fecal carriage of NB1000S (Fig. S10). The subject subsequently received 7 days of tetracycline 500 mg BID, which was followed by reduction of fecal NB1000S levels to below the LOD of the fecal qPCR assay (Fig. S10).

A further Stage 1 Group 3 subject, 102-154, was treated with amoxicillin/clavulanate 500/125 mg BID for 7 days, which similarly did not eliminate fecal carriage of NB1000S (Fig. S10). She was subsequently treated with 7 days of tetracycline 500 mg BID, which in this case was not followed by reduction of fecal NB1000S levels (Fig. S10).

One subject in Stage 1 Group 2X (Subject 102-158) was treated with oral cefdinir, a cephalosporin antibiotic, 300 mg BID for 7 days, for treatment of an incidental streptococcal pharyngitis. This did not eliminate fecal shedding of NB1000S (Fig. S10). This subject subsequently received a 7-day oral course of ciprofloxacin, a fluoroquinolone antibiotic, 500 mg BID for 7 days, for treatment of a urinary tract infection. After the ciprofloxacin course the fecal level of NB1000S declined to below the LOD of the fecal qPCR (Fig. S10).

Characterization of biocontainment escapes

To better understand the landscape of mutations that can enable biocontainment escape after porphyran removal, we characterized strains isolated from culture, from mice and from the Phase 1 healthy human volunteers. In culture, two strains containing the porphyran PUL either with or without biocontainment plus the porphyran promoter driving luciferase was each inoculated into duplicate 100 mL flasks that contained rich media and porphyran, in which fresh media lacking porphyran was flowed in at about a rate of 100 mL per 8 hours. Colony forming units (CFU) were quantified on plates with or without porphyran to determine abundance of total or biocontainment-escaped strains (Fig. 4A). While the non-biocontained strain remained highly abundance throughout the experiment, after the second day the biocontained strain had dropped ~1000-fold in abundance while the biocontainment escapes were rapidly increasing and eventually reached the same high level as the non-biocontained strains. From the terminal biocontainment culture, 385 strains were isolated, sequenced and luciferase was measured in the absence of porphyran. Many different biocontainment escapes were identified, with 95% of them resulting in high luciferase in the absence of porphyran (Fig. S11A). These high luciferase escapes contained mutations to the porphyran sensing HTCS that likely resulted in its constitutive activity. Most HTCS mutations identified occurred in or between the REC and HTH_ARAC domains, likely creating a conformational change similar to the one expected from phosphorylation of the REC domain. The other 5% of the mutants that did not produce high luciferase all contained transposon insertions or genome rearrangements that appeared to result in transcription of the essential gene independent from P_por10.

In the humanized mice shown in Figure 2C, in 3 animals representing 2 donors, persistent, low-level shedding of NB1000S was noted after porphyran was removed from the diet. Isolation of these persisting strains was technically challenging given the lack of antibiotic markers in the strain and the low overall colonization density. However, 6 clonal isolates originating from 2 cagemate animals were eventually cultured and subjected to whole genome sequencing. Across all 6 clones, sequencing highlighted the emergence in the porphyran hybrid-two component system (HTCS) of a single point mutation (S811L) that is directly upstream of the phosphorylatable histidine residue which likely results in activation of the HTCS and constitutive expression of the essential gene in the absence of porphyran.

In all 4 healthy volunteers with persistent fecal shedding, it was possible to establish the mechanism by which NB1000S had escaped this containment via genomic sequencing of subject-specific fecal isolates. In all cases, NB1000S harbored a distinct genomic mutation that caused the strain's conditional attenuation system to lose its responsiveness to porphyran. Metagenomic analysis suggests this was mediated by missense mutations in the porphyran-sensing HTCS at residues T675I and Q1111R in subjects 102-115 and 102-154, respectively. The Q1111R was also seen in a culture biocontainment escape and was associated with an activated P_por10 promoter. Subjects 102-151 and 102-158 appear to contain genomic rearrangements immediately upstream of the conditionally expressed essential gene, arginyl-tRNA synthetase (argS), similar to the 5% of mutations not in the HTCS from the culture experiment. Metagenomic analysis of the isolate from subject 102-151, suggests the strain contains a transposon integrated into the RBS of argS, likely providing transcription independent of P_por10. Metagenomic analysis was unable to determine the mechanism of persistence in subject 102-154, so it was isolated and whole genome sequenced, which uncovered an 81 kilobase inversion between porphyran-inducible argS promoter and an invertible sequence. This inversion eliminated a terminator upstream of argS, likely allowing readthrough transcription from BVU_1129.

Engineering multi-layered biocontainment

All observed biocontainment escapes appeared to be the result of rare gain-of-function HTCS mutations or genomic rearrangements targeting 462 bp region between the start of argS and an upstream terminator. We hypothesized that if multiple independent porphyran controlled biocontainment layers could be created (Fig. 4B), then the multiple mutations needed to break all layers of biocontainment would be exceedingly rare. Chimeric HTCSs have been generated to rewire signaling from a sensor domain to a different promoter (*18*). Using that approach we pursued creating two new chimeric HTCSs that have the porphyran-sensing domain fused to one of two unrelated regulatory domains that drive orthogonal promoters.

Initial testing of chimeric HTCS/promoter pairs generated from well-characterized fructan or arabinan components revealed repression in glucose-containing rich media (Fig. S11B), suggesting known Bacteroides cross-regulation to prioritize preferred carbon sources (*19*) may be problematic. We hypothesized that promoters with rare regulatory sequences would be less likely to have evolved extensive cross-regulation that would be preserved when moved to a different species. We focused on HTCS that had no known homolog outside of their species with more than 80% sequence identity in the promoter specificity determining HTH_ARAC domain. We also focused on HTCS with more than 25% identity to the porphyran sensing HTCS's Y_Y_Y domain that participates in transduction of signal from the sensor domain to the transmembrane and downstream signaling domains. Finally, we narrowed in on HTCSs from the PULDB that had associated promoters with apparent araC-type binding motifs. We selected three of these rare HTCSs and associated promoters and generated chimeras that showed a 2-to-15-fold induction of the associated promoter in response to porphyran only when the chimera was present (Fig. S11C).

Though all three chimeras displayed porphyran-responsiveness, none showed the 100-fold induction level we thought would be needed for robust biocontainment. To tune the chimeras to exhibit the large dynamic range needed for biocontainment, we generated a mutational library of Chimera 3, targeting the region near the fusion point and screened for a large fold-induction (Fig. S11D). The top hit exhibited more than a 100-fold induction in the presence of porphyran, with no significant cross-talk with the native porphyran dependent HTCS or promoter (Fig. 4C).

Expression of a second essential gene, cysteinyl-tRNA synthetases (cysS), was placed under control of our new chimera and screened using the method described in figure S3B to create a functional biocontainment layer. This layer of cysS biocontainment was combined with the original argS biocontainment to create a double biocontained (2xBC) strain. Unlike the previous biocontained strain (1xBC), the 2xBC strain successfully cleared from a chemostat (Fig. S11E). However, when tested for clearance *in vivo* in conventionally-raised mice, we observed escape via HTCS point mutation and apparent homologous recombination between the identical porphyran sensor domains in the two biocontained loci. To prevent this, we replaced the porphyran sensing domain with an alternate porphyran-sensing domain from a different porphyran PUL (*17*) that shared only 43% amino acid identity with the original porphyran sensing domain. In addition to avoiding homology, using a different domain may reduce the chances of shared non-specific interactions molecules other than porphyran. Swapping the sensor domain of the chimera altered the translation rate and required tuning of the chimera's RBS (Fig. S11F).

To create a third layer, we used the regulator domain of Chimera 1 (Fig. S11C). To minimize homology of the sensor domain with the other layers, we used a recoded sequence of the original porphyran sensing domain. We followed a similar strategy used to make the first chimera, mutating the chimera junction and tuning the HTCS RBS to make a functional chimera with more than a 100-fold porphyran induction. When targeting an essential gene, previously we had selected two tRNA synthetases, with the idea that since they were targeting the same process (translation), mutations to break one might have a reduced fitness advantage than if they were targeting unrelated processes. However, for the third layer, we thought that more rapid clearing may be beneficial, so murF was selected because it shows a more step like response to component concentration (*20*) and potential for killing cells rather than stalling translation. We used this chimera driving murF following the strategy described in figure S3B to first screen for a functional single layer of biocontainment, and then incorporating it into the previous 2XBC strain to generate to triple biocontained strain (3xBC).

<u>Performance of triple biocontained strain</u>

As an initial characterization, the 3xBC strain and a non-biocontained control were added to a 100 mL chemostat that was initially diluted at a rate of 100 mL per 12 hours with porphyran containing rich media, and after 3 days was switched to dilution with rich media lacking porphyran. Between 1 day and 4 days after porphyran had been removed from the feed media the 3xBC strain was rapidly cleared from almost $10^{12}$ CFU per flask to below the $10^{2}$ CFU per flask LOD (Fig. 4D). 10 days after porphyran had been removed from the media, porphyran was added to the flasks but no strains grew out over the following week, suggesting the 3x biocontained strain had been completely cleared.

Next, we tested the 3xBC strain and a non-biocontained control in individually housed mice originally fed a porphyran containing diet for the first week, in which both strains reach almost $10^{10}$ copies/g feces (Fig. 4E). Porphyran was then removed from the diet for 2 weeks and within the first 3 days the biocontained strain dropped to around the LOD of $10^{5}$ copies/g while the non-biocontained strain remained between $10^{8}$ and $10^{9}$ copies/g. Cages were changed on days 7, 12 and 21. After two weeks off porphyran, in which the 3xBC strain remained near or below the LOD, porphyran was reintroduced. All 6 mice with the control strain and 5 of the 6 mice with the biocontained strain returned to a $10^{9}$ to $10^{10}$ copies/g abundance, while one 3xBC mouse remained below the LOD, suggesting it had cleared. Strains were isolated from the 5 mice with detectable levels of 3xBC strains, and isolates were sequenced and phenotyped. Surprisingly, whole genome sequencing revealed no identifiable genomic mutations in any of the isolates relative to the gavage stocks. Phenotyping found that all 3x biocontained isolates retained the biocontainment phenotype, not growing on plates in the absence of porphyran (Fig. S12A). Though 3xBC attenuated the strain and did not mutate, failure to clear in most mice suggests a persistence mechanism not seen in the chemostat.

We also tested the 3xBC strains in the context of germ-free mice colonized with microbiota from three healthy volunteer subjects that failed to clear NB1000S, using samples from prior to the introduction of NB1000S. After the community was established, either 1x or 3x biocontained strains were gavaged while mice were on a porphyran containing diet. After porphyran was removed from the diet, all strains showed a 2 to 4 log reduction in the colonization, but none of the strains completely cleared from any of the 3 microbiotas. However, in the two most permissive microbiotas the 3xBC strains dropped to a significantly lower level than the 1xBC strain. In the least permissive of the three microbiotas, both strained dropped to between the LOD of $10^{5}$ and $10^{6}$ copies/g. This suggests the 3x biocontainment further attenuates the strain relative to 1x biocontainment but is not sufficient for clearance.

Because of the lack of clearance of the 3x biocontained strains, despite no evidence of mutational escape, we engineered biocontainment into other bacteroides species to investigate if the cryptic mechanism of persistence could be specific to our *B. vulgatus* strain. Two different species, *B. ovatus* and *B. thetaiotaomicron* were each engineered with two versions of 2xBC (argS + cysS or argS + murF). *B. ovatus* and *B. thetaiotaomicron* strains with 1xBC (argS, cysS or murF) or either 2xBC all appeared biocontained in culture. The new 2xBC strains were tested in mice, alongside the 3xBC *B. vulgatus* and non-biocontained versions of each species. All strains contained a porphyran PUL. Strains were gavaged into mice on a porphyran containing diet. When porphyran was removed from the diet, all non-biocontained species dropped about one log. 2xBC strains dropped about 2 logs, indicating some attenuation but not clearance, similar to *B. vulgatus*. Strains were isolated at the end of the experiment and two isolates were found to have a mutation in one of the two biocontainment layers, while most strains showed no mutations and retained their biocontained phenotype in culture.

Engraftment in enteric hyperoxaluria patients

In the Phase 2 part of the clinical trial NOV-001-CL01, engraftment with NB1000S was less consistent and was overall lower in EH patients as compared to that seen in HV receiving the same NB2000P dose as in Group 2 (10 g/day). Only 3 of the 9 EH patients who received active NOV-001 treatment achieved peak stool carriage at a level (approximately $10^{10}$ genomes/g) comparable to that seen in HV at the higher NB2000P dose levels tested in Stage 1 (subjects 121-212, 125-204 and 121-216; purple-, pink- and grey-colored lines in figure S13); and in one of these patients this level was only reached at the end of the 4-week period of treatment with NB2000P (Subject 121-216; grey line in figure S13). A direct comparison between Stage 1 and Stage 2 is not possible as no Stage 1 subjects had received exactly the same NOV-001 regimen as used in Stage 2 (in Stage 1 the change to 2 doses of NB1000S was made only in Group 2X, who received a higher porphyran dose than used in Stage 2).

In 4 patients the highest NB1000S levels seen were between $10^8$ to $10^9$ genomes/g. In 2 of these patients the level dropped to between $10^4$ and $10^5$ genomes/g during the treatment period. In 1 subject (125-206; gold-colored line in figure S13) the fecal NB1000S qPCR was marginally above the LOD at $1.29 \times 10^4$ genomes/g on a single day during the treatment period (study Day 8) and was < LOD throughout the remainder of the treatment and follow-up periods.

During screening 1 EH patient, Subject 103-203, was determined to be carrying porphyranase-positive fecal bacteria at baseline, with a level of $3.8 \times 10^7$ genomes/g at Screening and $1.05 \times 10^8$ genomes/g at Day 1, as determined by qPCR for the porphyranase locus. Fecal qPCR for the NB1000S oxalate degradation locus in Subject 103-203 remained below LOD throughout the Treatment and Follow-up periods of the study (red-colored line in figure S13).

Large horizontal gene transfer events detected in patients

In addition to the low engraftment levels noted on the NB1000S fecal qPCR for some EH patients, discrepancies were noted between the fecal qPCR targeting the oxalate degradation locus and the qPCR targeting the porphyranase locus (Fig. 5B). No such instances were noted in Stage 1. Although no Stage 2 patient had persistent fecal carriage of NB1000S following cessation of NB2000P based on the fecal qPCR targeting the oxalate degradation locus, 6 of the 9 NB1000S-dosed patients were found to have persistent fecal shedding of porphyran-utilizing organisms based on qPCR targeting the porphyranase locus (Fig. S14A). Some patients showed porphyranase-positive organisms at levels between $10^5$ and $10^{10}$ genomes/g despite the qPCR for the oxalate degradation locus being below the $10^4$ limit of detection (LOD), or had porphyranase qPCR results that were significantly higher than those obtained from the oxalate locus qPCR in the same fecal sample. Retesting a subset of the samples with qPCR primers that target different regions of the oxalate pathway or porphyran PUL confirmed this result results.

Three potential explanations for the discrepancy are (1) presence of a native porphyran utilizer, (2) deletion of the oxalate pathway from NB1000S or (3) transfer of the porphyran PUL from NB1000S to another member of the microbiota. A plot of the porphyran PUL and oxalate pathway over time for each patient (Fig. S14A), shows 1 patient (subject 103-203) is colonized with a native porphyran utilizer and 5 patients have a divergence between porphyran PUL and oxalate pathway which appears only after NB1000S introduction. An initial metagenomics analysis of fecal samples suggested that in subject 125-202 the oxalate pathway was missing from a genome that aligns to NB1000S, while in subject 117-201 the porphyran PUL is present in a genome that has low identity outside of the porphyran PUL suggesting horizontal gene transfer to another strain. The PUL strain abundance was not high enough in the remaining three subjects to suggest an explanation from metagenomic analysis.

To further investigate this discrepancy of low oxalate pathway and high porphyran PUL copies, we isolated the porphyran PUL containing strains from patient samples. Fecal samples from a point in which porphyran PUL copies were high but the oxalate copies were low were selected (Fig. S14A, red arrows). Selected samples were grown in defined media with gentamycin and with porphyran as the sole carbon source, plated, and then two colonies that gave positive porphyran PUL qPCR signal were subjected to whole genome sequencing. All isolates sequenced from the same patients appeared to be the same strain except in patient 125-204, where two distinct strains were isolated.

Mapping the WGS results to the NB1000S genome revealed some regions with near perfect matches (Fig. 5D, blue) and other regions with a high frequency of SNPs or deletions (Fig. 5C, grey) that indicate that portion of the genome was not derived from NB1000S. Comparing the location of the biocontainment, oxalate pathway and porphyran PUL to these regions (Fig. 5C, green box) indicates that in three patients (110-204, 117-201 and 121-212) an outgoing HGT event has transferred the porphyran PUL from NB1000S to another strain, in one patient (124-202) an incoming HGT event has replaced a >500 kb stretch of the NB1000S containing biocontainment and the oxalate pathway, and in the patient (125-204; two different strains were isolated) one strain showed an incoming HGT event and the other showed an outgoing HGT event.

To investigate potential mechanisms of outgoing HGT of the porphyran PUL, we attempted to isolate the pre-HGT base strains prior to incorporating the porphyran PUL using the baseline patient fecal sample before NB1000S was dosed. For each strain that had incorporated the porphyran PUL, we determined the antibiotic resistance profile and unique sequences that can be targeted via qPCR. Baseline fecal samples were each plated on plates containing antibiotics matching the HGT strain's resistance profile, and colonies were picked and screened with qPCR. Positive qPCR isolates were then sequenced revealing isolates with near perfect matching of the majority of the baseline strain to the post-HGT isolate, indicating that each strain had been successfully isolated. To visualize the origins of the post-HGT strains genomes, we compare each to the associated the pre-HGT genome or to NB1000S (Fig. S14B) and see the porphyran PUL (red) from NB1000S and some surrounding genome (blue) has been introduced to all three of the post-HGT isolates. Additionally, multiple ~1 kb regions in patients 110-204 and 117-201 isolates or >10 kb regions in the patient 121-212 isolate do not match the pre-HGT strain or NB1000S. It also appears that the isolate from patient 212-212 contains two >10 kb regions of DNA from NB1000S that are separated by almost a third of the genome.

Several elements potentially relevant to HGT were identified in the post-HGT strains that were not found in NB1000S or the pre-HGT strains. In the patient 125-202 isolate a 161 kb region with many phage associated genes was identified. The patient 125-204 incoming HGT isolate and the 110-204 isolate contain different regions with type IV secretory system conjugative DNA transfer proteins. The patient 125-204 isolate also contained an ICE-like element. We also noticed in all three isolated pre-HGT strains that transposases were adjacent to the site of integration. Many hypothetical genes were introduced into all post-HGT strains, potentially associated with uncharacterized phage or other HGT-relevant functions. Because of the wide variety of introduced HGT-associated genes and the multiple rearrangements between the pre-HGT and post-HGT genomes, we were unable to determine with confidence the specific HGT mechanisms responsible.

Splitting porphyran PUL components reduces HGT *in vitro*

Considering these Phase 2 HGT findings, efforts to reduce HGT could substantially improve this platform. Multiple methods have been proposed for reducing HGT, which offer promising solutions to prevent HGT when used alone or in combination (*14, 21*). We tested one simple solution of splitting the porphyran PUL elements into different locations in the chromosome to see if HGT events could be reduced.

We were unable to reproduce *in vitro* the large >500kb HGT events seen in the clinic but did observe incoming transfer of antibiotic resistance into NB1000S at rates of $\sim 10^{-8}$ when strains were co-cultured on solid media. To test whether splitting up the porphyran PUL reduced rates of outgoing HGT, measurably high rates of outgoing HGT were essential. Chromosomal conjugal elements in Bacteroides containing tetracycline resistance have been shown to increase the rate of transfer of other mobilizable elements to recipient strains. Also, pretreatment with tetracycline of HGT donor strains can further increase rates of transfer (*22*). Here, we test the ability of the donor strain NB1000S with an integrative conjugative element containing tetracycline resistance (ICE-TetR) to transfer an NBU-integrated antibiotic resistance gene to two different recipient strains, a *Bv* type strain, and the pre-HGT *Bv* isolate from a Ph2a participant. Both ICE-TetR integration and tetracycline induction increased conjugation efficiency increase, up to $2 \times 10^{-6}$ in the pre-HGT strain (Fig. S15A).

To test whether splitting up the porphyran PUL reduced transfer of the complete PUL to recipient strains, a complementation model was used. The recipient strain contained almost the entire porphyran PUL except for two genes, a glycoside hydrolase (GH) and a sulfatase, both essential for porphyran utilization. Donor strains contained the GH and sulfatase, integrated via NBU at the same locus (together) or at two different loci (separate). Donor strains also contained one of two ICE-TetR elements to increase rates of HGT. Transconjugants were identified by growth on selective minimal media with porphyran as the sole carbon source, since only transconjugants with both the GH and sulfatase would be able to utilize porphyran. Two different donor strains were tested, and transconjugants were only observed when donor strains contained the GH and sulfatase at the same locus (Fig. S15B).

Patient urine oxalate analysis

In the Phase 2 part of clinical study NOV-001-CL01 urine oxalate was measured as a preliminary assessment of efficacy. One of the inclusion criteria was 24-hour urinary oxalate (UOx) ≥ 60 mg based on the mean of two adequate collections during Screening at least 3 days apart, and neither < 40 mg oxalate. High levels of variability in UOx (mg/24hr) were observed, including more than 2-fold changes between readings for the same subject on different collection days, and a relatively low correlation ($R^2$=0.20) between those subsequent collections (Fig. S16A). Normalizing the level of oxalate against the level of creatinine (mg/mL) in the urine collection, done commonly to compensate for incomplete 24-hour collections and for variation in hydration state / fluid intake, yielded an oxalate to creatinine ratio (OCR) which gave a modest improvement in the correlation ($R^2$=0.34) between subsequent Screening measurements (Fig. S16B).

In assessing preliminary efficacy, five criteria were predefined comparing active treatment to control for the absolute or percent change in UOx, the percent change in OCR, and the proportion of patients achieving ≥ 20% reduction in 24-hour UOx or OCR. All five predefined criteria showed slight improvements in active compared to placebo groups, but none were statistically significant. For example, for the criteria "proportion of patients achieving ≥ 20% reduction in 24-hour OCR excretion from baseline to end of treatment, NOV-001 compared to placebo" we found an insignificant improvement from 33% of placebo to 44% of treatment (Fig. S16C).

In an *post hoc* analysis, to reduce the impact of variability for an individual measured at different times, we averaged the three pre-NB1000S measurements (two screening days and day 1) and compared those to the average of the last three measurements of the treatment (days 14, 21 and 28), which reduced variability within groups, and showed an increased the impact of treatment to a 27% reduction in OCR (Fig. S16D). Another factor possibly obscuring any signal are the observation that NB1000S did not successfully engraft in 3 of the 9 patients. Some correlation between higher colonization and lower UOx was observed (Fig. S16E). Since we expect successful engraftment to be required for efficacy, we considered analysis dividing the subjects into placebo, treated non-engrafters and treated engrafters, delineated in Fig. S14A, which shows placebo and non-engrafters group together while engrafted subjects show a noticeable but not statistically significant reduction in UOx (Fig. S16F) or OCR (Fig. 5E). Placebo and non-engrafted subjects together compared to engrafted subjects shows statistically significant reductions in UOx (Fig. 15G) and OCR (Fig. 15H). The ~40 mg/24h UOx reduction seen in 3 of the engrafted subjects is a promising magnitude as UOx level above 40 mg/24h are considered the threshold for hyperoxaluria diagnosis and the changes in UOx was not correlated with starting UOx concentrations ($R^2$=0.01).

Overall, the data suggests the potential for efficacy of NOV-001 in treating hyperoxaluria but remains inconclusive. The sample sizes in this initial clinical trial were not necessarily intended to provide sufficient power to show statistically significant differences between treatment groups. However, the positive directional change in predefined endpoints and the significance observed in *post hoc* analysis suggests further studies into efficacy might be worthwhile. If strain improvements to reduce the observed oxalate dependent toxicity were made and increased the colonization levels to be more consistent with Phase 1, further reduction in UOx than observed here could be a possibility.

Safety Evaluation

The primary endpoint for both Stages 1 and 2 of the study was the assessment of safety and tolerability of NOV-001, including both serious and non-serious AEs, and assessments of the incidence, nature, severity, and relatedness of treatment-emergent adverse events (TEAEs) to study product. Based on the mechanism of action and the nonclinical experience NOV-001 was not anticipated to pose significant safety risks for humans. Details of the safety evaluation can be found in the NOV-0001-CL01 Clinical Study Report.

Treatment-emergent AEs (TEAE) reported in Stage 1 of Study NOV-001-CL01 are summarized by study group according to maximum severity and to the PI's assessment of relatedness to study product in Table S5. TEAEs reported in Stage 1 of Study NOV-001-CL01 are also summarized by study group according to MedDRA System Organ Classification (SOC) and Preferred Term (PT) according to the number of individuals with a given PT event in Table S6.

Treatment-emergent AEs (TEAE) reported in Stage 2 of Study NOV-001-CL01 are summarized by study group according to maximum severity and to the PI's assessment of relatedness to study product in Table S7. TEAEs reported in Stage 2 of Study NOV-001-CL01 are also summarized below by study group and SOC and PT according to the number of individuals with a given PT event in Table S8.

In Study NOV-001-CL01 there were no deaths, no withdrawals from the study or serious adverse events (SAE) attributed to study product, and no subjects reported to have adjusted dose level due to adverse events (AEs). The majority of AEs were mild and transient, and events of a gastrointestinal nature predominated (e.g. flatulence, diarrhea). Review of the AE profiles from the subjects who had persistent fecal shedding of NB1000S after treatment cessation does not suggest a significant clinical safety concern. Assessment of safety and tolerability is limited by the small number of subjects and short duration of treatment in this study; however, there were no clinical safety issues identified that would preclude further clinical development or which would require additional safety provisions in subsequent studies.

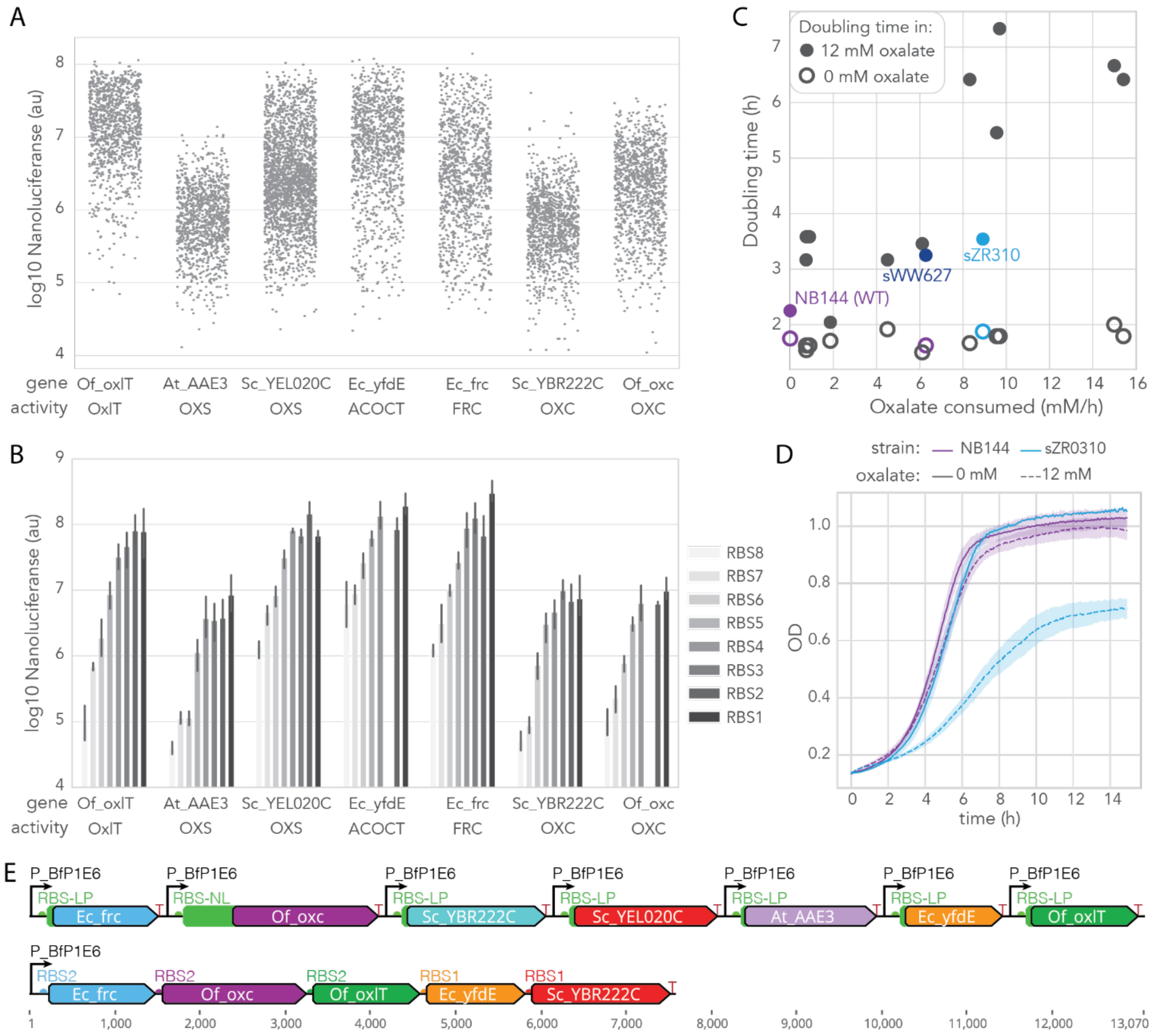


**Fig. S1. Oxalate pathway was optimized to balance between high oxalate degradation and oxalate dependent toxicity.** (**A**) RBS libraries from seven genes found to increase oxalate degradation in preliminary oxalate degrading strains were screened for each gene via luminescence from translational fusion nanoluciferase. Points represent a single measurement of each library member. (**B**) Different library members with varying RBS strengths were selected and remeasured in triplicate. (**C**) Eleven new single-operon strains, along with previous versions and a non-degrading control, were screened for oxalate degradations rates cells concentrated to $10^{10}$ CFU/ml and for growth rate in rich media with 0 mM or 12 mM oxalate(x-axis) and doubling times (y-axiS). Open circle or closed circle indicate growth rates in measured 0 mM or 12 mM oxalate, respectively. Highlighted are the non-oxalate degrading control (NB144; purple), the fastest oxalate degrading strain prior to optimization (sWW627; dark blue) and the optimized strain selected for advancement (sZR310; light blue). (**D**) $OD_{600nm}$ growth curves show growth of control (NB144; purple) or oxalate degrading strain (sZR310; light blue) grown in rich media with 0 mM (solid lines) or 12 mM (dashed lines) oxalate. (**E**) Oxalate pathway diagram for pre-optimization sWW627 (above) and optimized sZR310 (below).

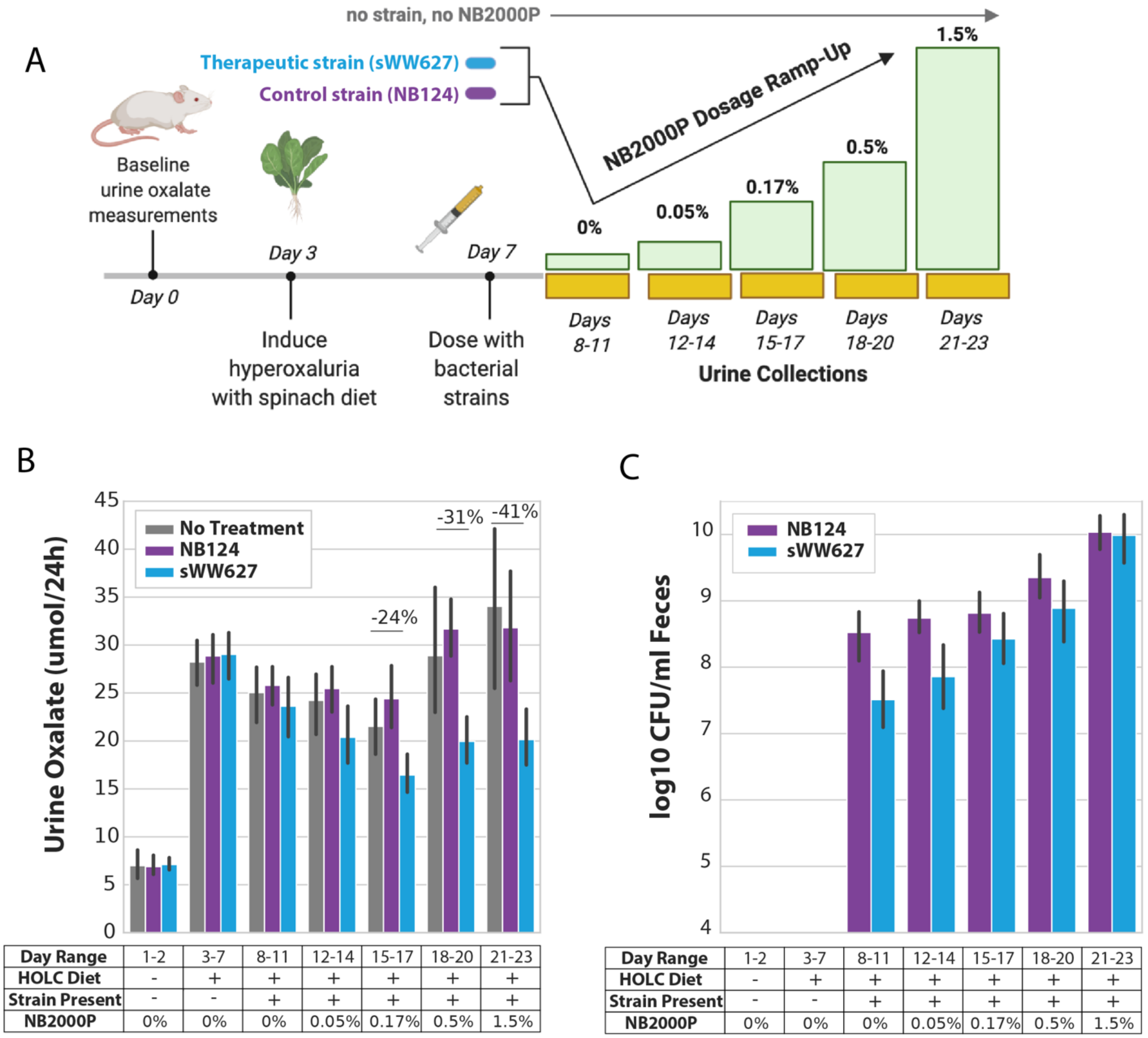


**Fig. S2. Oxalate-degrading strain significantly reduces urine oxalate in a rodent model of SH in porphyran dose dependent manner.** (**A**) Graphical summary of study design. Hyperoxaluria was induced from baseline in Sprague-Dawley rats ($n = 8$ animals per group) by the addition of spinach to the low-calcium diet starting on Day 2 (HOLC diet). On Day 7, animals were split into three groups: no treatment control, or orally administered one dose of $10^9$ CFU of either the control strain NB124 containing only the porphyran-utilization modification (purple), or sWW627 containing both porphyran-utilization and oxalate degradation modifications (blue). After day 8, the two strain treatment groups were exposed to increasing concentrations of porphyran. Yellow bars indicate continuous urine collection in relation to dietary changes (represented as green boxes), which are made at the beginning of the 24-hour interval whereas urine is collected at the end of each 24 hour interval. Individual animal urine oxalate levels and fecal strain abundance were quantified throughout the study duration. (**B**) Urine oxalate levels (μmol/24 h) shown for each treatment group over study duration as denoted by color, binned into day ranges corresponding to experimental conditions as summarized in table below the x-axis. Reduction in daily urine oxalate excretion between control and engineered strain groups was statistically significantly different where noted (data are mean ± 95% confidence interval, T-test). (**C**) Fecal strain abundance (data are mean ± 95% CI) for the two colonized treatment groups as denoted by color are shown over study duration, binned into day ranges corresponding to experimental conditions summarized in table below the x-axis. CFU were statistically significantly different at Days 8-11 ($p$ = 0.004), Days 12-14 ($p$ = 0.010), T-test.

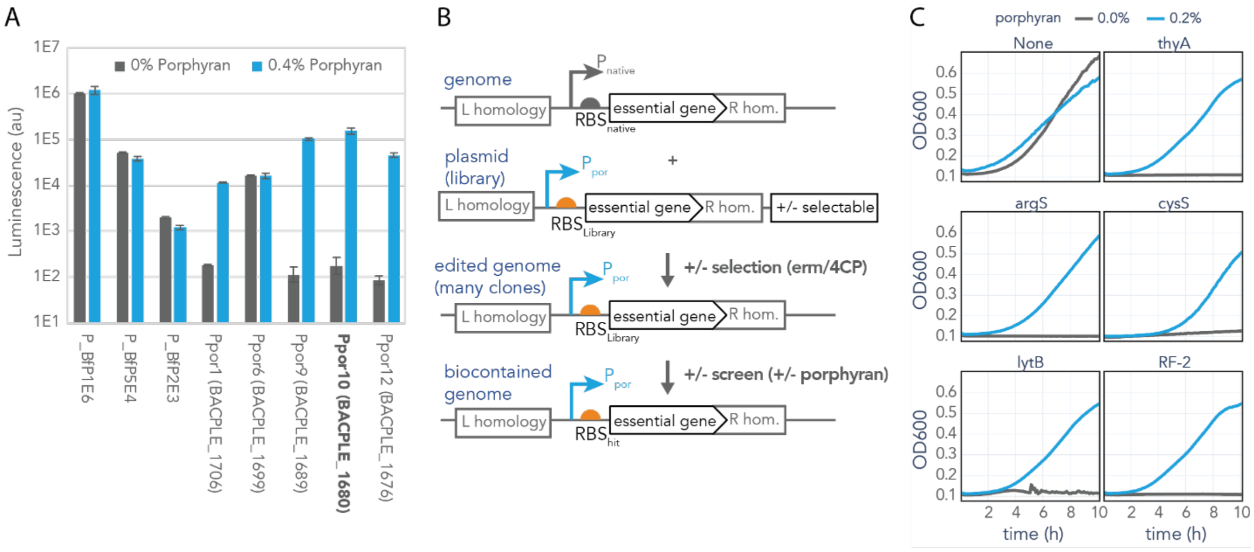


**Fig. S3. Strains are biocontained by replacing native essential gene promoter and RBS with a porphyran inducible promoter and tuned RBS.** (**A**) Luminescence is measured from constitutive promoters (left three) and selected promoters from the porphyran PUL (right five) driving luciferase expression in rich media with 0% (grey) or 0.4% (blue) porphyran. (**B**) Genome modification schematic showing starting genome and plasmid (top), result after both positive and then negative selection of plasmid components (middle), and biocontained strain after screen (bottom). (**C**) $OD_{600nm}$ growth curves in rich media with 0% (grey) or 0.4% (blue) of control (upper left) or biocontained strains labeled by the porphyran-regulated essential gene.

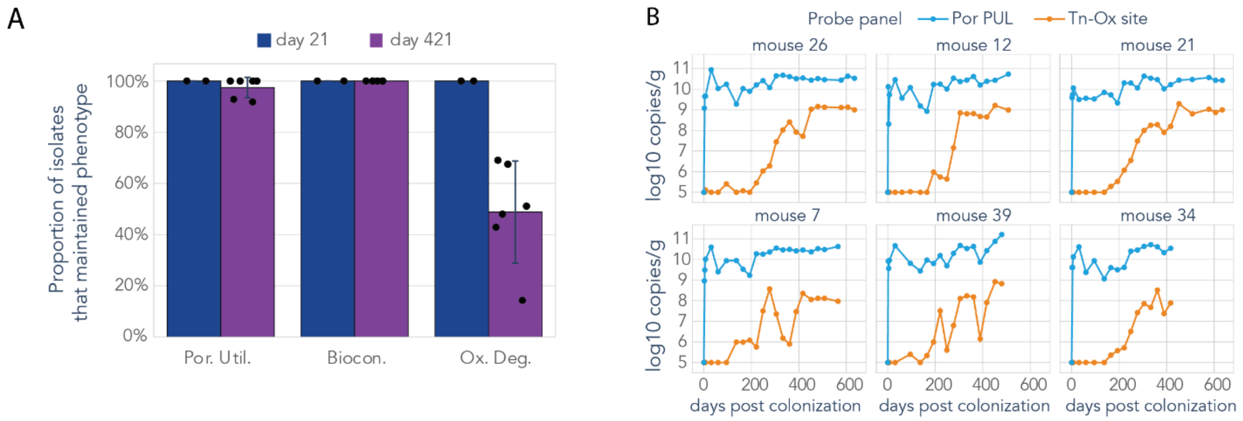


**Fig. S4. Long-term colonization shows NB1000S porphyran and biocontainment maintained, while mutations breaking oxalate pathway gradually accumulate.** (**A**) Percentage of isolates from each mouse on day 21 (blue) or day 421 (purple) that passed phenotypic tests for growth on porphyran as a sole carbon source (left), no growth in rich media lacking porphyran (center), or complete removal of oxalate from media (right) are plotted. Black points represent proportion of strains isolated from individual mice. (**B**) Log10 copies per gram feces assessed via qPCR of porphyran PUL (blue) or common transposon insertion that eliminates oxalate consumption (orange) are plotted over the more than 400 days the mice are colonized.

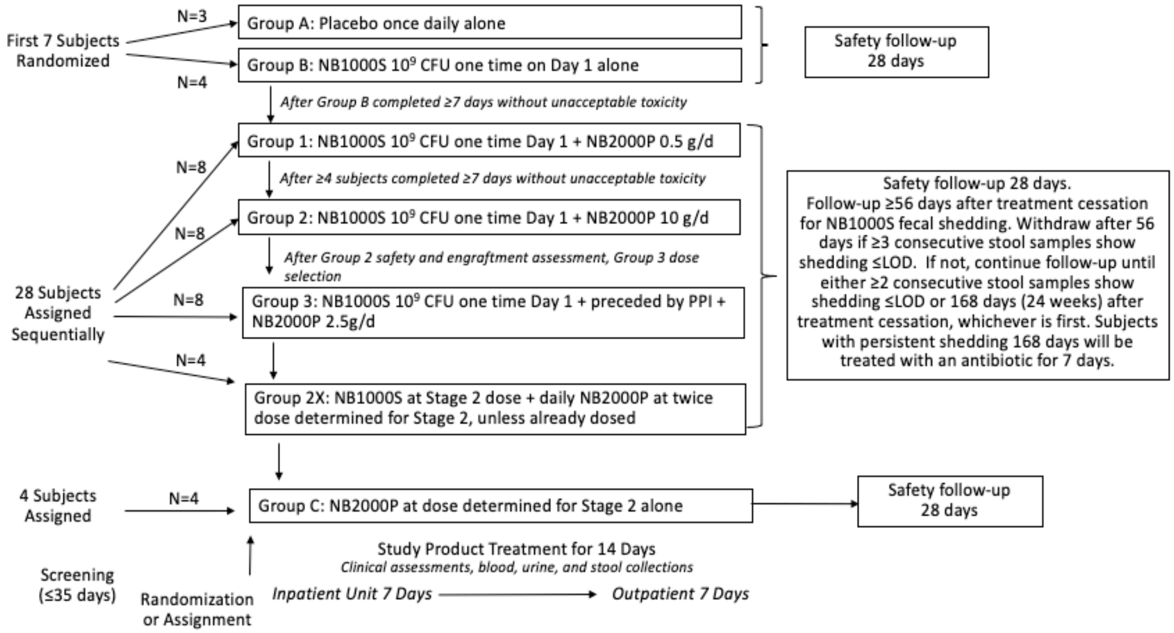


**Fig. S5. Clinical design: Stage 1 Schematic and Actual Enrollment**.

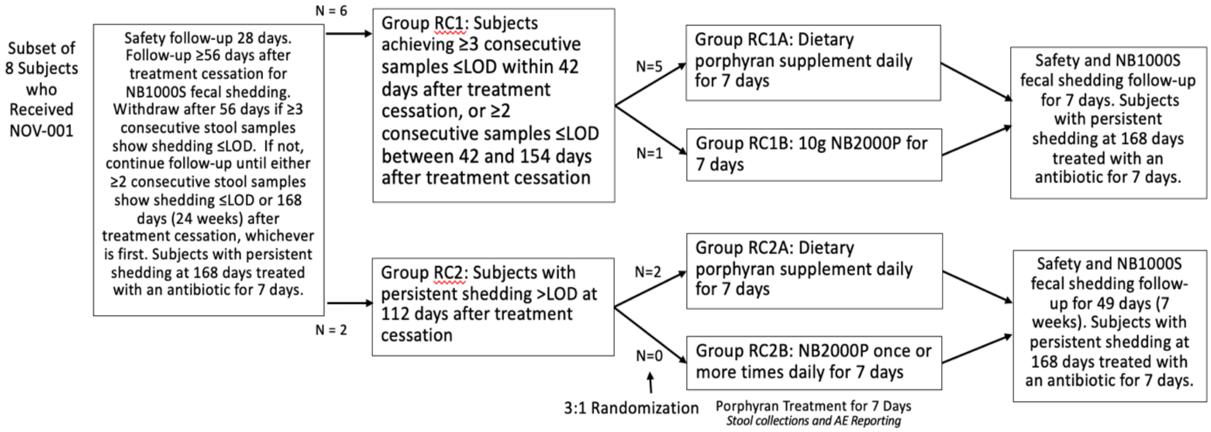


**Fig. S6. Clinical design: Stage 1 Porphyran Rechallenge Schematic and Actual Enrollment**

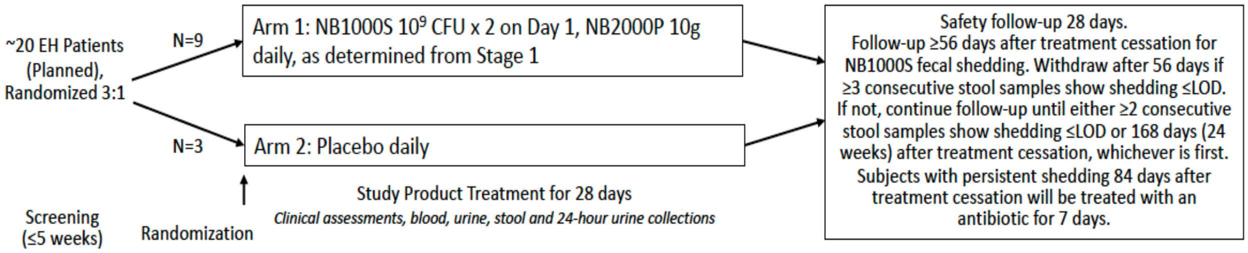


**Fig. S7. Clinical design: Stage 2 Schematic and Actual Enrollment**

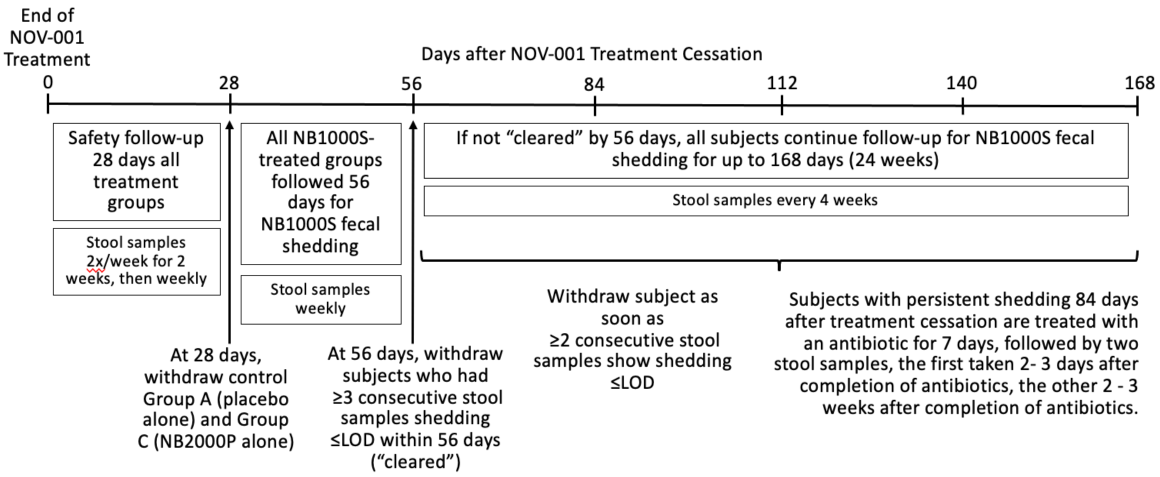


**Fig. S8. Clinical design: Follow-Up Periods**

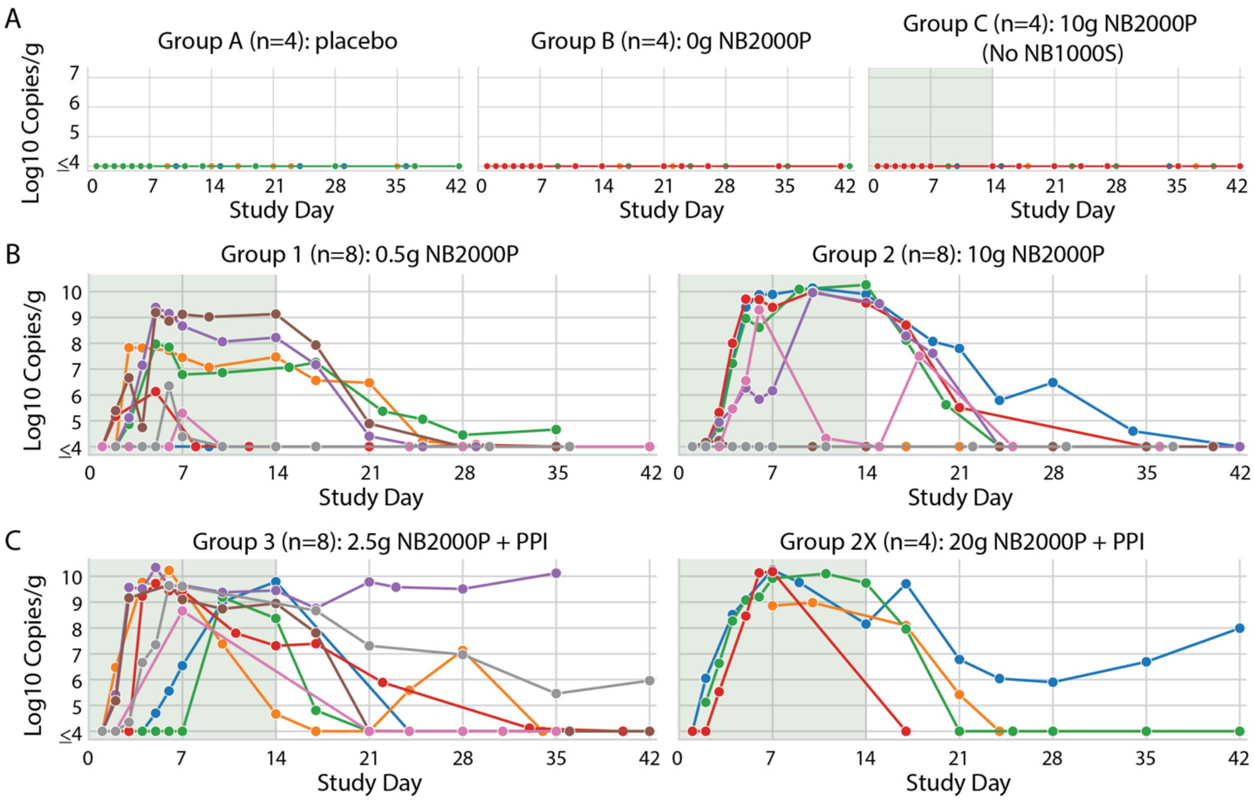


**Fig. S9. NB1000S robustly engrafts in healthy volunteers when dose is preceded by omeprazole.** (**A**) Copies of NB1000S are not detected in the feces of healthy volunteers above the $10^4$ copies/g limit of detection (LOD) for groups dosed placebo (Group A), NB1000S strain without NB2000P (Group B) or 10g NB2000P without strain (Group C). (**B**) Copies of NB1000S are detected in the feces during porphyran dosing (green) of most healthy volunteers above the LOD for patients dosed once with antacid then NB1000S and then daily administered porphyran at 0.5 g/day (Group 1) or 10 g/day (Group 2). (**C**) Copies of NB1000S are detected in the feces during porphyran dosing (green) of all healthy volunteers above the LOD for patients dosed once with antacid and omeprazole then NB1000S and then daily administered porphyran at 2.5 g/day (Group 3) or 10 g/day twice a day (Group 2X).

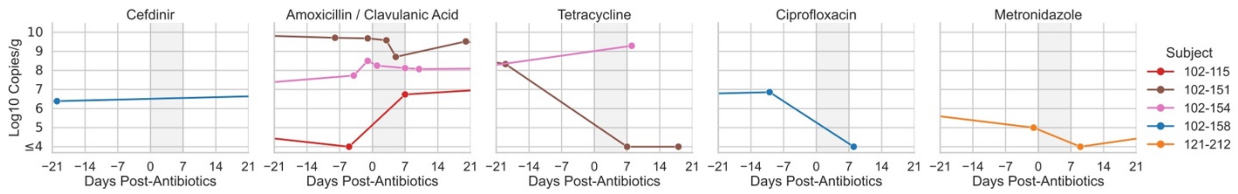


**Fig. S10. Effect of antibiotic treatment on carriage of NB1000S.** Subjects with persistent fecal shedding of NB1000S were treated with antibiotics either per Protocol or for incidental infections. Shading represents the period of antibiotic treatment. Fecal qPCR's targeted the porphyranase locus.

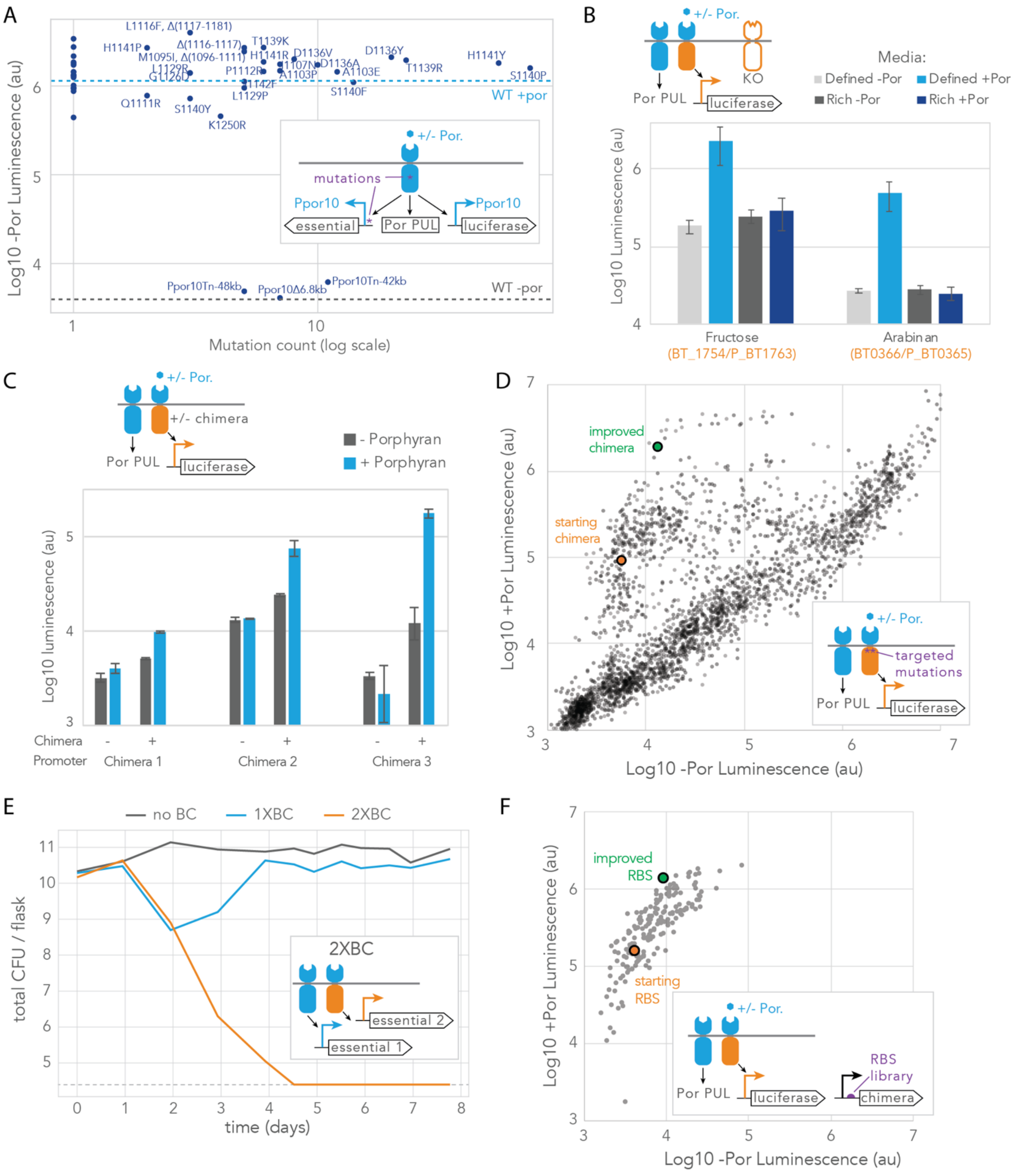


**Fig. S11. HTCS chimera creation and tuning creates additional layer of biocontainment to make 2xBC strain.** (**A**) Biocontainment escapes from culture are characterized by frequency of mutations observed (of the 385 screened), genotype (labeled by HTCS residue or P_por10 disruption for all mutations occurring more than once), and activity of P_por10 driven luciferase reporter of HTCS activity in the absence of porphyran. Wildtype activity is shown with the dashed lines in the presence (light blue) or absence (dark blue) of porphyran. (**B**) HTCS chimeras designed using the porphyran sensing domain fused to the fructose (left) or arabanan (right) sensing domains are measured driving luciferase in strains with the native fructose (BT_1754) or arabanan (BT_0366) HTCS knocked out. Luminescence is measured in defined media with glucose (light grey) or porphyran (light blue) as the sole carbon source, or in rich media (BHIS) with 0% porphyran (dark grey) or 0.5% porphyran (dark blue). Lack of induction in glucose-containing rich media (dark blue) suggests cross-repression in common regulatory elements may be problematic. (**C**) HTCS chimeras were designed using the porphyran sensing domain fused to rare HTCS regulatory domains. Luminescence is shown from associated promoter driving luciferase in rich media (BHIS) with 0% porphyran (dark grey) or 0.5% porphyran (dark blue) alone (left) or with the Chimera present (right). (**D**) Luminescence from mutations around the transmembrane region of Chimera 3 with the associated promoter driving luciferase are shown without (x-axis) or with (y-axis) porphyran. Each point represents a member of the mutation library and the orange point represents the starting Chimera 3. (**E**) Strains with no biocontainment (grey), the original 1x biocontainment (blue) or the new mutated Chimera 3 dependent double biocontainment (orange) were inoculated into a chemostat. The chemostat initially contained porphyran, which was steadily diluted out as culture was plated on porphyran containing plates to estimate CFU per flask (y-axis). Limit of detection is indicated with dashed line. (**F**) Members of an RBS library of chimera 3 mutant with altered porphyran sensor coding are plotted with luminescence in the absence (x-axis) or presence (y-axis) of porphyran.

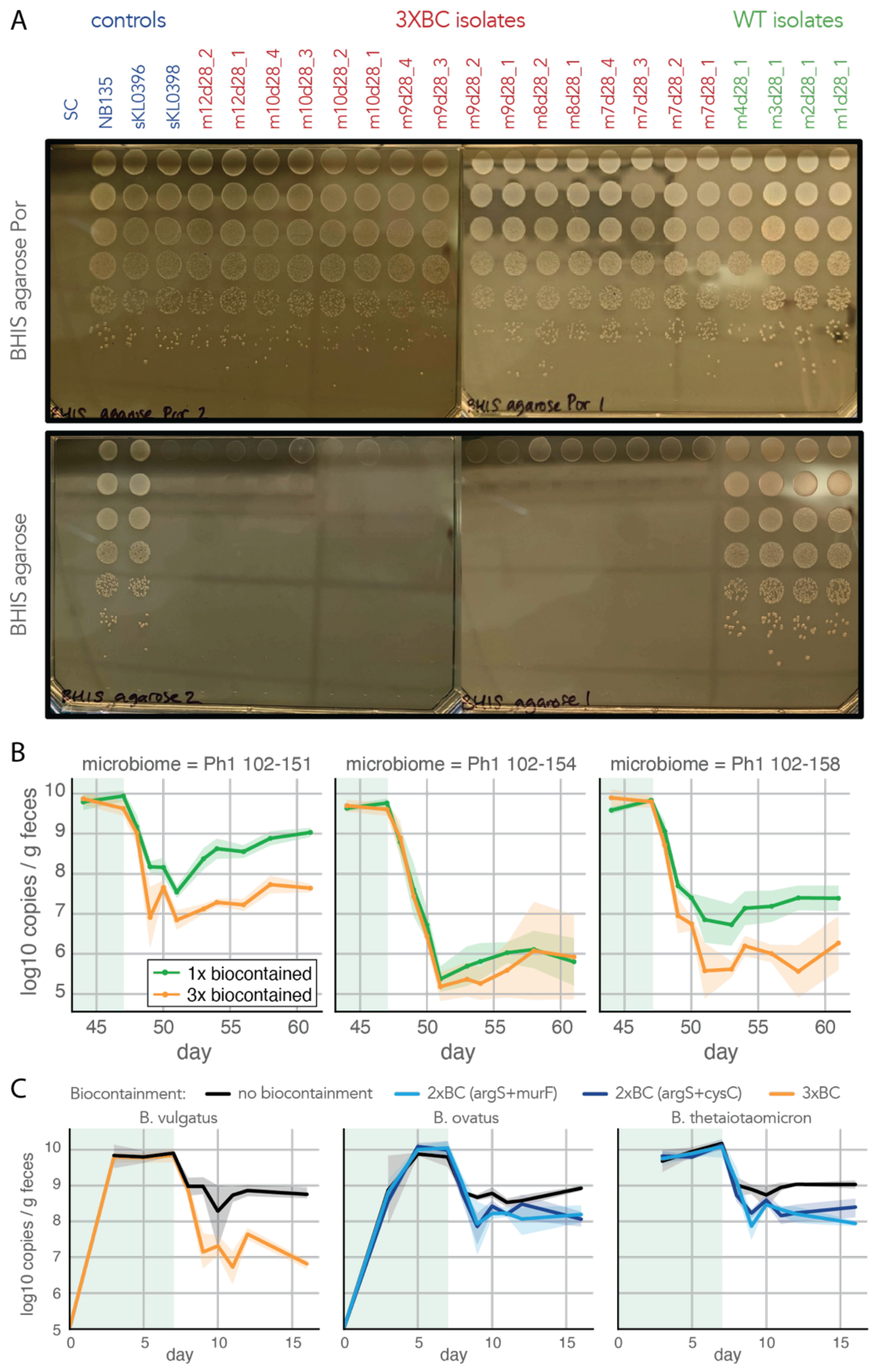


**Fig. S12. Biocontained strains can persist in vivo in the absence of porphyran without mutating.** (**A**) 3xBC isolates that failed to clear from mice (red) and non-biocontained control isolates (green) and original stock of control strains (blue; SC=sterile control, NB135 is Bv type strain, sKL0396 is porphyran consuming non-biocontained strain, and sKL0396 is 3xBC strain) are dilution (10x per row) plated on agarose plates with or without porphyran. (**B**) Gnotobiotic mice were colonized with communities from three human subjects in which NB1000S failed to clear, using samples taken before NB1000S dosing. Average log10 copies per gram feces of the strain (via porphyran PUL qPCR) are plotted for the NB1000S, single biocontained (green), and the triple biocontained (orange) strains. Shaded area is 95% confidence interval. Green highlighting is period of porphyran treatment. LOD is $10^5$ copies per gram. (**C**) Mice were gavaged with *Bv* with 3xBC, or *Bo* and *Bt* with 2xBC (using argS+murF in light blue or argS+cysS in dark blue), or non-biocontained *Bv, Bo,* or *Bt* with porphyran PUL. Porphyran is initially in the diet and is removed on day 7. Log10 copies of porphyran PUL are plotted. Shaded area is 95% confidence interval. Green highlighting is period of porphyran treatment. LOD is $10^5$ copies per gram.

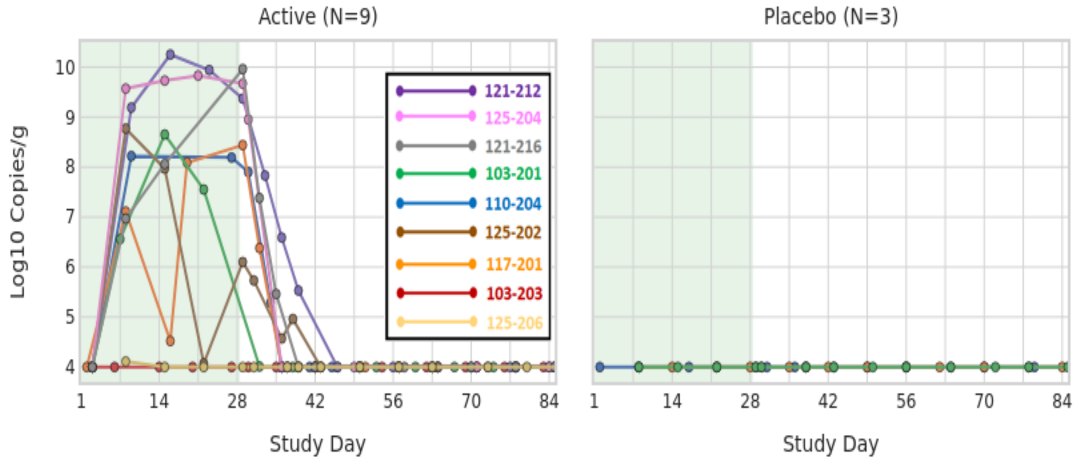


**Fig. S13. Fecal qPCR for NB1000S, Stage 2**
Enteric hyperoxaluria patients in Stage 2 received omeprazole 40mg ER followed by two doses of NB1000S 109 CFU on Day 1, and NB2000P 10 g/day during the 4-week treatment period (green shaded area). Legend with subject identifiers included for NOV-001 actively-treated subjects only.

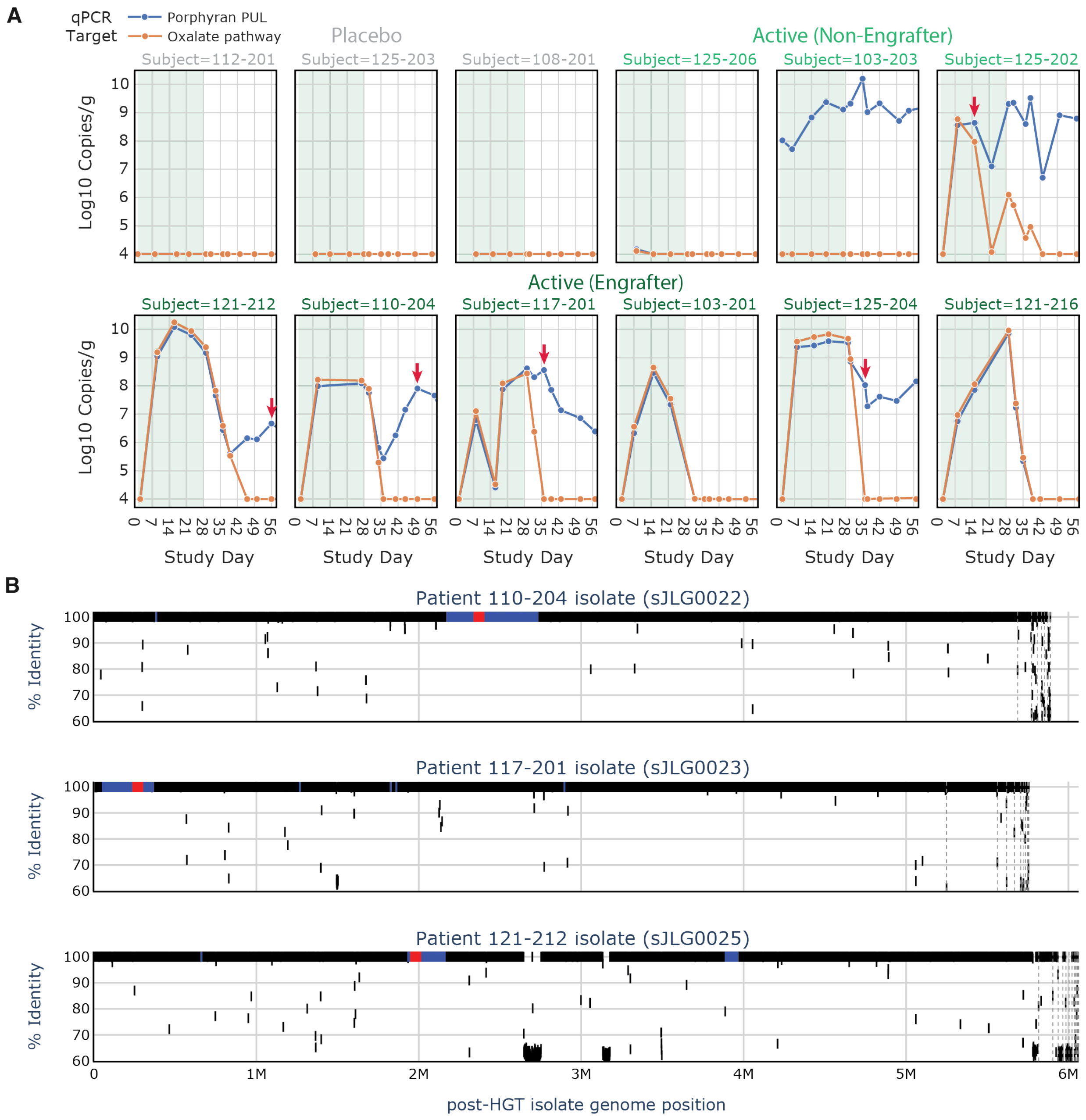


**Fig. S14. Porphyran consuming strains lacking oxalate pathway were mapped to pre-HGT isolate genome to identify introduced DNA.** (**A**) Abundance of the porphyran PUL (blue) and oxalate pathway (orange) assessed via qPCR is shown for patients given placebo (grey; 112-201, 124-103 and 108-201), patients treated with NB1000S that failed to engraft (light green; 125-206, 103-202 and 125-202) and patients treated with NB1000S that engrafted (dark green; 121-212, 110-204, 117-201, 103-201, 125-204 and 121-216). Porphyran treatment duration shown in green. Point of post-HGT strain isolation shown with red arrows. LOD is $10^4$ copies/g. (**B**) Post-HGT strain percent identity to the pre-HGT strain (isolated before NB1000S was dosed) at each position in the post-HGT strain genome (1 kb bins) is plotted. Regions do not match the pre-HGT strain but have 100% identity to NB1000S are highlighted in blue or in red if the NB1000S match is to the porphyran PUL. Dashed vertical lines separate contigs of post-HGT genome.

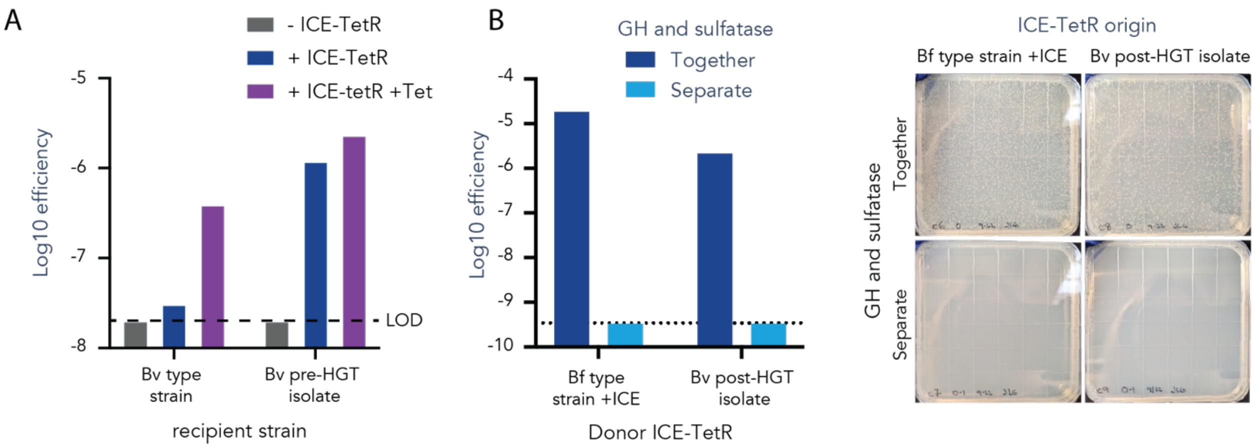


**Fig. S15. Separating porphyran PUL components on the chromosome reduces rates of outgoing HGT.** (**A**) To increase transfer rates for HGT assay, a donor strain was created by integrating an ICE-TetR NB1000S carrying the NBU-integrated antibiotic resistance and transfer to one of two recipient strains (*Bv* type strain on left or *Bv* pre-HGT clinical isolate on right) was quantified. HGT efficiency (fraction of recipient colonies to receive antibiotic resistance) is plotted for donor lacking the ICE-TetR, with the ICE-TetR, or with the ICE-TetR and exposure to tetracycline is plotted. The limit of detection (LOD) represents a single colony on the transconjugant selection plate. (**B**) HGT assay was adjusted to quantify transfer of porphyran PUL components. The recipient strain contained almost the entire porphyran PUL except for two genes, a glycoside hydrolase (GH) and a sulfatase, both essential for porphyran utilization. Donor strains contained the GH and sulfatase, integrated via NBU at the same locus (together) or at two different loci (separate). Donor strains also contained one of two ICE-TetR elements to increase rates of HGT. Transconjugants were identified by growth on selective minimal media with porphyran as the sole carbon source, since only transconjugants with both the GH and sulfatase would be able to utilize porphyran. Two different donor strains were tested, and transconjugants were only observed when donor strains contained the GH and sulfatase at the same locus.

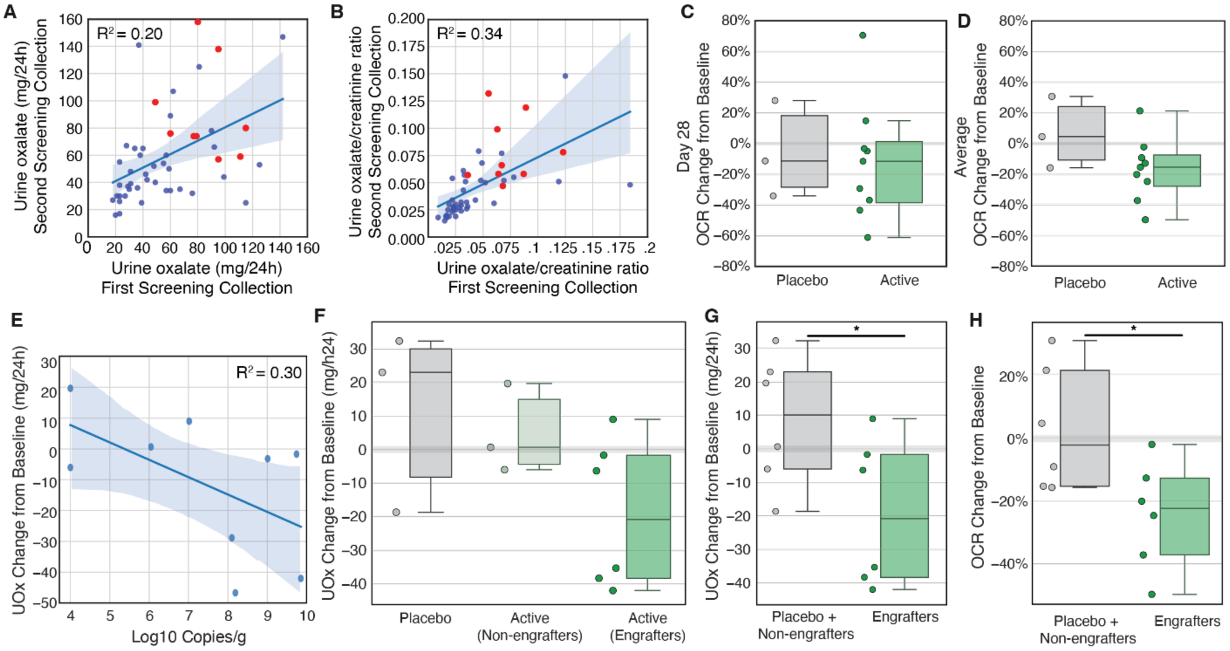


**Fig. S16. Urine oxalate is lower in enteric hyperoxaluria patients treated with NOV-001 compared to placebo.** (**A**) Comparisons between the two 24-hour urinary oxalate excretion (UOx; mg/24h) values obtained from two urine collections done as part of the Screening process for NOV-001-CL01 Stage 2. Randomized subjects are highlighted in red. Fit for all samples ($R^2$=0.20) shown by blue line with blue shading representing 95% confidence interval. (**B**) Same comparison as (A) but showing urine oxalate concentrations (mg/mL) divided by urine creatinine concentrations (mg/mL) to get an oxalate to creatinine ratio (OCR), giving a fit with $R^2$=0.34. (**C**) Change in OCR measured between Screening baseline and last day of treatment (day 28) for randomized patients given Placebo or Active (treated with NOV-001). (**D**) Average change in OCR measured between pre-NB1000S samples (Screening baseline and day 1) and last three samples in treatment (days 14, 21 and 28) for randomized patients given Placebo or Active (**E**) Comparison of Log10 Copies/g of NB1000S oxalate and average change in UOx from pre-NB1000S baseline for Active patients. Fit shown by blue line ($R^2$=0.30) with blue shading representing 95% confidence interval. (**F**) Average change in UOx compared to pre-NB1000S baseline for patients given Placebo, patient given NOV-001 where NB1000S did not successfully engraft (defined as colonization on the last day lower than $10^7$ copies/g), and patient given NOV-001 where NB1000S successfully engrafted. (**G**) Comparison of average UOx between combined patients given Placebo and active patients where NB1000S failed to engraft (grey) and active patients where NB1000S successfully engrafted (green; P=0.04). (**H**) Comparison of average OCR change from pre-NB1000S baseline between combined patients given Placebo and Active patients where NB1000S failed to engraft (grey) and Active patients where NB1000S successfully engrafted (green; P=0.03).

| Strain | Figures | Brief description | Species | Phenotype | Detailed description |
|---|---|---|---|---|---|
| **NB075** | S3C,S11E | Bv + porphyran PUL | B. vulgatus | por+, ermR | porphyran PUL (variant) B. vulgatus ATCC8482 |
| **NB1000S** | 1A, 2B, 2C, 3A, 3B, 3C, 4E, 5B, 5C, 5D, 5E, S4A, S4B, S5, S6, S7, S8, S9A, S9B, S9C, S10, S12B, S13, S14A, S14B, S16A-H | Therapeutic strain (Bv + porphyran PUL + 1xBC + oxalate pathway) | B. vulgatus | por+, porAUX | arginyl-tRNA_synthetase-Term23-Ppor10s6v7-RJ-RBS(sWW180)-argS-OriT-TetQ-KanR1-KanR2-BxEPheSx2-R6K + BvInt-15A-P_BfP1E6-sWW874L-RBS2-Ec_frc!-sWW871L-RBS2-Of_oxc!-sWW875L-RBS5-Of_oxlT!-sWW873L-RBS2-Ec_yfdE!-sWW870L-RBS2-Sc_YBR222C!-term-BvInt-15B-ErmG-BxEPheSx2 + porphyran PUL (markerless) B. vulgatus DSM 1447 / ATCC 8482 |
| **NB124** | 1B, 1C, S2A, S2B, S2C | Bv + porphyran PUL | B. vulgatus | por+, ermR, camR | camR + porphyran PUL (markerless) B. vulgatus DSM 1447 / ATCC 8482 |
| **NB144** | 2C, 4D, S1C, S1D | Bv + porphyran PUL (base strain used to make NB1000S) | B. vulgatus | por+ | porphyran PUL (markerless) B. vulgatus DSM 1447 / ATCC 8482 |
| **sBB1498** | 4D | Bv + porphyran PUL + 3xBC | B. vulgatus | por+, porAUX | revcomp(P_BT1311-sDZ0212-RBS-Por recode1-17106RR Chimera HTCS)-Term23-P_17106-3-sDZ0255-RBS-UDP-N-acetylmuramoylalanyl-D-glutamyl-2, 6- diaminopimelate-D-alanyl-D-alanyl ligase + revcomp(P_17150-2-V2-sDZ0218RBS-Por(todS_19)-17150RR Chimera HTCS-term)-P_17150-1-V8-CysS-RBS_C8-cysteinyl-tRNA_synthetase + Term23-Ppor10s6v7-RJ-RBS(sWW180)-argS + porphyran PUL (markerless) B. vulgatus DSM 1447 / ATCC 8482 |
| **sCG838** | S11B | Bo (porphyran utilizer) + porphyran/arabinan chimeric HTCS + arabinan reporter | B. ovatus | por+, tetR, ermR, | Ppor6_RBS(wt)_HTCS_Porphyran(1-798)-Arabinan(899-1420)!-term P_arabinan(P_BT0365)-RJ-RBS(E139J15)-NanoLuc!-term Porphyran utilizing Bacteroides ovatus isolate |
| **sCG850** | S11B | Bo (porphyran utilizer) + porphyran/fructose chimeric HTCS + fructose reporter | B. ovatus | por+, tetR, ermR, | Ppor6_RBS(wt)_HTCS_Porphyran(1-797)-Fructose(406-921)!-term P_fruc(P_BT1763)-RJ-RBS(E139J15)-NanoLuc!-term Porphyran utilizing Bacteroides ovatus isolate |
| **sDZ0028L** | S11F | Bv + por. PUL + RBS library for porphyran chimera 3 + chimera 3 reporter | B. vulgatus | por+, ermR | CamR2-ColE1-rcCon5-P_BfP5E4-[sfGFP-ePheS AarI Dropout RBS]-Por(todS_19)-17150RR Chimera HTCS-term-rcConS-CamR1-CamR2-ColE1-Con5-Term23-P_17150-1-V8-RJ-RBS(E139J15-Luc)-NanoLuc!-term-ConE-CamR1-oriT-ErmG-KanR1-KanR2-NBU2-attN2-R6K-RBSlib2 + porphyran PUL (markerless) B. vulgatus DSM 1447 / ATCC 8482 |
| **sDZ0276** | 4E,S12B | Bv + por. PUL + 3xBC + oxalate pathway | B. vulgatus | por+, porAUX | revcomp(P_BT1311-sDZ0212-RBS-Por recode1-17106RR Chimera HTCS)-Term23-P_17106-3-sDZ0255-RBS-UDP-N-acetylmuramoylalanyl-D-glutamyl-2, 6- diaminopimelate-D-alanyl-D-alanyl ligase + revcomp(P_17150-2-V2-sDZ0218RBS-Por(todS_19)-17150RR Chimera HTCS-term)-P_17150-1-V8-CysS-RBS_C8-cysteinyl-tRNA_synthetase + Term23-Ppor10s6v7-RJ-RBS(sWW180)-argS + BvInt-15A-P_BfP1E6-sWW874L-RBS2-Ec_frc!-sWW871L-RBS2-Of_oxc!-sWW875L-RBS5-Of_oxlT!-sWW873L-RBS2-Ec_yfdE!-sWW870L-RBS2-Sc_YBR222C!-term-BvInt-15B + porphyran PUL (markerless) B. vulgatus DSM 1447 / ATCC 8482 |
| **sHR0005** | 4C | porphyran HTCS + porphyran reporter | B. vulgatus | por+, tetR, ermR | Ppor10-RJ-RBS(E139J15-Luc)-NanoLuc!-term + Ppor6-RBS(HTCSwt)-Porphyran HTCS-term-ErmG + porphyran PUL (markerless) B. vulgatus DSM 1447 / ATCC 8482 |
| **sHR0006** | 4C | chimera 3 + porphyran reporter | B. vulgatus | tetR, ermR, | Ppor10-RJ-RBS(E139J15-Luc)-NanoLuc!-term + Ppor6-RBS(HTCSwt)-Por-17150 Chimera (e3-118-s34)-term-ErmG + porphyran PUL (minus porphyran HTCS) B. vulgatus DSM 1447 / ATCC 8482 |
| **sHR0009** | 4C | porphyran HTCS + chimera 3 reporter | B. vulgatus | por+, tetR, ermR | P_17150-1-V8-RJ-RBS(E139J15-Luc)-NanoLuc!-term-TetQ + Ppor6-RBS(HTCSwt)-Porphyran HTCS-term-ErmG + porphyran PUL (markerless) B. vulgatus DSM 1447 / ATCC 8482 |
| **sHR0010** | 4C | chimera 3 + chimera 3 reporter | B. vulgatus | tetR, ermR | P_17150-1-V8-RJ-RBS(E139J15-Luc)-NanoLuc!-term-TetQ + Ppor6-RBS(HTCSwt)-Por-17150 Chimera (e3-118-s34)-term-ErmG + porphyran PUL (minus porphyran HTCS) B. vulgatus DSM 1447 / ATCC 8482 |
| **sJLG0015** | S14B | 110-204 B. vulgatus HGT partner strain isolate | B. vulgatus | tetR | 110-204 B. vulgatus HGT partner strain isolate |
| **sJLG0016** | S14B, S15A | 117-201 B. vulgatus HGT partner strain isolate | B. vulgatus | tetR | 117-201 B. vulgatus HGT partner strain isolate |
| **sJLG0017** | S14B, S15A | 121-212 B. vulgatus HGT partner strain isolate | B. vulgatus | tetR, ermR, | 121-212 B. vulgatus HGT partner strain isolate |

**Table S1. Strains in this study**

| | | | | | |
|---|---|---|---|---|---|
| **sJLG0021** | S15A | NB1000S + camR | B. vulgatus | por+, porAUX, camR | camR + NB1000S: arginyl-tRNA_synthetase-Term23-Ppor10s6v7-RJ-RBS(sWW180)-argS-OriT-TetQ-KanR1-KanR2-BxEPheSx2-R6K + BvInt-15A-P_BfP1E6-sWW874L-RBS2-Ec_frc!-sWW871L-RBS2-Of_oxc!-sWW875L-RBS5-Of_oxlT!-sWW873L-RBS2-Ec_yfdE!-sWW870L-RBS2-Sc_YBR222C!-term-BvInt-15B + porphyran PUL (markerless) B. vulgatus DSM 1447 / ATCC 8482 |
| **sJLG0022** | S14B | 110-204 HGT strain isolate | B. vulgatus | por+, tetR | 110-204 HGT strain isolate |
| **sJLG0023** | S14B | 117-201 HGT strain isolate | B. vulgatus | por+, tetR, ermR, | 117-201 HGT strain isolate |
| **sJLG0025** | S14B | 121-212 HGT strain isolate | B. vulgatus | por+, tetR, ermR, | 121-212 HGT strain isolate |
| **sJLG0029** | S15A | NB1000S + camR + sJLG0017_TetQ_ICE | B. vulgatus | por+, tetR, porAUX, camR | sJLG0017_Tet(Q)_ICE + camR + NB1000S: arginyl-tRNA_synthetase-Term23-Ppor10s6v7-RJ-RBS(sWW180)-argS-OriT-TetQ-KanR1-KanR2-BxEPheSx2-R6K + BvInt-15A-P_BfP1E6-sWW874L-RBS2-Ec_frc!-sWW871L-RBS2-Of_oxc!-sWW875L-RBS5-Of_oxlT!-sWW873L-RBS2-Ec_yfdE!-sWW870L-RBS2-Sc_YBR222C!-term-BvInt-15B-ErmG-BxEPheSx2 + porphyran PUL (markerless) B. vulgatus DSM 1447 / ATCC 8482 |
| **sJLG0050** | S15B | Bv + porphyran GH29/sulfatase (together) + Bf-ICE-TetR | B. vulgatus | tetR, ermR | Prom-RBS-GH29-RBS-sulfatase-Term-ConEnoBsaI-OriT-ErmG-KanR1-KanR2-NBU2-attN2-R6K-ConSnoBsaI + NB191 ICE-TetR + B. vulgatus DSM 1447 / ATCC 8482 |
| **sJLG0052** | S15B | Bv + porphyran GH29/sulfatase (together) + isolate-ICE-TetR | B. vulgatus | tetR, ermR | Prom-RBS-GH29-RBS-sulfatase-Term-ConEnoBsaI-OriT-ErmG-KanR1-KanR2-NBU2-attN2-R6K-ConSnoBsaI + sJLG0017 ICE-TetR + B. vulgatus DSM 1447 / ATCC 8482 |
| **sJLG0054** | S15B | Bv + porphyran GH29 + sulfatase (separate) + Bf-ICE-TetR | B. vulgatus | tetR, ermR, camR | Prom-RBS-sulfatase-Term-ConEnoBsaI-OriT-CamR-KanR1-KanR2-NBU2-attN2-R6K-ConSnoBsaI + Prom-RBS-GH29-Term-ConEnoBsaI-OriT-ErmG-KanR1-KanR2-NBU2-attN2-R6K-ConSnoBsaI + NB191 ICE-TetR + B. vulgatus DSM 1447 / ATCC 8482 |
| **sJLG0056** | S15B | Bv + porphyran GH29 + sulfatase (separate) + isolate-ICE-TetR | B. vulgatus | tetR, ermR, camR | Prom-RBS-sulfatase-Term-ConEnoBsaI-OriT-CamR-KanR1-KanR2-NBU2-attN2-R6K-ConSnoBsaI + Prom-RBS-GH29-Term-ConEnoBsaI-OriT-ErmG-KanR1-KanR2-NBU2-attN2-R6K-ConSnoBsaI + sJLG0017 ICE-TetR + B. vulgatus DSM 1447 / ATCC 8482 |
| **sKL0396** | S12A | Bv + porphyran PUL + porphyran reporter | B. vulgatus | por+, tetR, ermR, camR | camR + tetR + Ppor10-RJ-RBSe100s23-NanoLuc-terminator-ermG + porphyran PUL (markerless) B. vulgatus DSM 1447 / ATCC 8482 |
| **sKL0398** | S12A | Bv + porphyran PUL + porphyran reporter + 3xBC | B. vulgatus | tetR, ermR, porAUX, camR | camR + tetR + Ppor10-RJ-RBSe100s23-NanoLuc-terminator-ermG + revcomp(P_BT1311-sDZ0212-RBS-Por recode1-17106RR Chimera HTCS)-Term23-P_17106-3-sDZ0255-RBS-UDP-N-acetylmuramoylalanyl-D-glutamyl-2, 6- diaminopimelate-D-alanyl-D-alanyl ligase + revcomp(P_17150-2-V2-sDZ0218RBS-Por(todS_19)-17150RR Chimera HTCS-term)-P_17150-1-V8-CysS-RBS_C8-cysteinyl-tRNA_synthetase + Term23-Ppor10s6v7-RJ-RBS(sWW180)-argS + porphyran PUL (markerless) B. vulgatus DSM 1447 / ATCC 8482 |
| **sWD1371** | S12C | Bo + 2xBC (P_por-argS, P_chimera3-cysS) | B. ovatus | por+, ermR, porAUX, | NB206 Ppor-cysS_v1 LILO + NB206 Ppor-argS_v1 LILO + Porphyran PUL + Bacteroides ovatus DSM 1896 / ATCC 8483 |
| **sWD1372** | S12C | Bo + 2xBC (P_por-argS, P_chimera3-murF) | B. ovatus | por+, ermR, porAUX, | NB206 Ppor-murF_v1 LILO + NB206 Ppor-argS_v1 LILO + Porphyran PUL + Bacteroides ovatus DSM 1896 / ATCC 8483 |
| **sWD1373** | S12C | Bt + 2xBC (P_por-argS, P_chimera3-cysS) | B. theta. | por+, ermR, porAUX | NB207 Ppor-cysS_v1 LILO + NB207 Ppor-argS_v1 LILO + Porphyran PUL + Bacteroides thetaiotaomicron DSM 2079 / VPI-5482 / ATCC 29148 |
| **sWD1374** | S12C | Bt + 2xBC (P_por-argS, P_chimera3-murF) | B. theta. | por+, ermR, porAUX | NB207 Ppor-murF_v1 LILO + NB207 Ppor-argS_v1 LILO + Porphyran PUL + Bacteroides thetaiotaomicron DSM 2079 / VPI-5482 / ATCC 29148 |
| **sWW039** | S3A | Bo (porphyran utilizer) + porphyran reporter (P_por10) | B. ovatus | ermR | Ppor10-RJ-RBSe100s23-NanoLuc-terminator Porphyran utilizing Bacteroides ovatus isolate |
| **sWW090** | S3C | Bo (porphyran utilizer) + 1xBC (P_por thyA) | B. ovatus | ermR | ThyA biocontained strain: term-Ppor10-RJ-RBSlib-ThyA-NanoLuc-term + DE(thyA) Porphyran utilizing Bacteroides ovatus isolate |
| **sWW180** | S3C, S11E | Bo (porphyran utilizer) + 1xBC (P_por argS) | B. ovatus | por+, ermR, porAUX | reverseGFP-Term-Ppor10-RBShit-arginyl-tRNA_synthetase porphyran PUL (variant) B. vulgatus ATCC8482 |
| **sWW202** | S3C | Bo (porphyran utilizer) + 1xBC (P_por cysS) | B. ovatus | ermR | reverseGFP-Term-Ppor10-RBS_hit-cysteinyl-tRNA_synthetase porphyran PUL (variant) B. vulgatus ATCC8482 |
| **sWW205** | S3C | Bo (porphyran utilizer) + 1xBC (P_por lytB) | B. ovatus | ermR | reverseGFP-Term-Ppor10-RBS_hit-cytidylate kinase + penicillin tolerance protein LytB porphyran PUL (variant) B. vulgatus ATCC8482 |
| **sWW206** | S3C | Bo (porphyran utilizer) + 1xBC (P_por RF2) | B. ovatus | ermR | reverseGFP-Term-Ppor10-RBS_hit-peptide chain release factor RF porphyran PUL (variant) B. vulgatus ATCC8482 |
| **sWW554** | 1B | Bv + porphyran PUL + 2x early oxalate pathway | B. vulgatus | por+, tetR, ermR | 2x(P_BfP1E6-LP18FUS-Ec_frc!-term-P_BfP1E6-RJ-RBS-NLuc-Of_oxc!-term-P_BfP1E6-LP18FUS-Sc_YBR222C!-term-P_BfP1E6-LP18FUS-Sc_YEL020C!-term-P_BfP1E6-LP18FUS-At_AAE3!-term-P_BfP1E6-LP18FUS-Ec_yfdE!-term-P_BfP1E6-LP18BCD-Of_oxlT!-term) porphyran PUL (variant) B. vulgatus ATCC8482 |

**Table S1. Strains in this study (continued)**

| | | | | | |
|---|---|---|---|---|---|
| **sWW627** | 1C, S1C, S2A, S2B, S2C | Bv + porphyran PUL + 2x early oxalate pathway | B. vulgatus | por+, tetR, ermR | 2x(P_BfP1E6-LP18FUS-Ec_frc!-term-P_BfP1E6-RJ-RBS-NLuc-Of_oxc!-term-P_BfP1E6-LP18FUS-Sc_YBR222C!-term-P_BfP1E6-LP18FUS-Sc_YEL020C!-term-P_BfP1E6-LP18FUS-At_AAE3!-term-P_BfP1E6-LP18FUS-Ec_yfdE!-term-P_BfP1E6-LP18BCD-Of_oxlT!-term) porphyran PUL (markerless, variant) B. vulgatus ATCC8482 |
| **sWW650** | S11C | Bv + porphyran PUL + chimera 3 reporter | B. vulgatus | por+, tetR | P_por-17150-V6-RBS(E139J15-Luc)-NanoLuc!-term-TetQ + porphyran PUL (markerless, variant) B. vulgatus ATCC8482 |
| **sWW658** | S11C | Bv + porphyran PUL + chimera 1 reporter | B. vulgatus | por+, tetR | P_por-17106-V4-RBS(E139J15-Luc)-NanoLuc!-term-TetQ + porphyran PUL (markerless, variant) B. vulgatus ATCC8482 |
| **sWW755** | S11C | Bv + porphyran PUL + chimera 2 reporter | B. vulgatus | por+, tetR | P_10809-4-RBS(E139J15-Luc)-NanoLuc!-term-TetQ + porphyran PUL (markerless, variant) B. vulgatus ATCC8482 |
| **sWW757** | S11C | Bv + porphyran PUL + chimera 2 reporter + chimera 2 | B. vulgatus | por+, tetR, ermR | Ppor6-RBS(HTCSwt)-Por-10809-chim1-term-ErmG + P_10809-4-RBS(E139J15-Luc)-NanoLuc!-term-TetQ + porphyran PUL (markerless, variant) B. vulgatus ATCC8482 |
| **sWW769** | S11C | Bv + porphyran PUL + chimera 1 reporter + chimera 1 | B. vulgatus | por+, tetR, ermR | Ppor6-RBS(HTCSwt)-Por-17106-chim1-term-ErmG + P_por-17106-V4-RBS(E139J15-Luc)-NanoLuc!-term-TetQ + porphyran PUL (markerless, variant) B. vulgatus ATCC8482 |
| **sWW777** | S11C | Bv + porphyran PUL + chimera 3 reporter + chimera 3 | B. vulgatus | por+, tetR, ermR | Ppor6-RBS(HTCSwt)-Por-17150-chim1-term-ErmG + P_por-17150-V6-RBS(E139J15-Luc)-NanoLuc!-term-TetQ + porphyran PUL (markerless, variant) B. vulgatus ATCC8482 |
| **sWW942** | S11E | Bv + porphyran PUL + 2xBC (P_por-argS, P_chimera3-lytB) | B. vulgatus | por+, ermR, porAUX, camR | Term23-Ppor10s6v7-RJ-RBS(sWW180)-argS + Term23-P_17150-1-V8-RBS1-cytidylate kinase + penicillin tolerance protein LytB + Ppor6-RBS(HTCSwt)-Por-17150 Chimera (e3-118-s34)-term-ErmG+ porphyran PUL (markerless, variant) B. vulgatus ATCC8482 |
| **sWW1059L** | S11D | Bv + porphyran PUL + chimera 3 reporter + chimera 3 junction library | B. vulgatus | por+, tetR, ermR | Ppor6-RBS(HTCSwt)-PorHTCS-library-17150HTCS-chim1-term-ErmG + P_17150-1-V8-RJ-RBS(E139J15-Luc)-NanoLuc!-term-TetQ + porphyran PUL (variant) B. vulgatus ATCC8482 |
| **sZR0103** | 4A | Bv + porphyran PUL + porphyran reporter | B. vulgatus | por+, tetR, ermR | Ppor10-RJ-RBS(E139J15-Luc)-NanoLuc!-term + porphyran PUL (variant) B. vulgatus ATCC8482 |
| **sZR0250** | 4A, S11A | Bv + porphyran PUL + porphyran reporter + 1xBC (P_por argS) | B. vulgatus | por+, tetR, ermR | Ppor10-RJ-RBS(E139J15-Luc)-NanoLuc!-term + reverseGFP-Term-Ppor10-RBShit-arginyl-tRNA_synthetase + porphyran PUL (variant) B. vulgatus ATCC8482 |
| **sZR0310** | S1C, S1D | Bv + porphyran PUL + oxalate pathway | B. vulgatus | por+, tetR, ermR | BvInt-15A-P_BfP1E6-sWW874L-RBS2-Ec_frc!-sWW871L-RBS2-Of_oxc!-sWW875L-RBS5-Of_oxlT!-sWW873L-RBS2-Ec_yfdE!-sWW870L-RBS2-Sc_YBR222C!-term-BvInt-15B-ErmG-BxEPheSx2 + porphyran PUL (markerless) B. vulgatus DSM 1447 / ATCC 8482 |

**Table S1. Strains in this study (continued)**

| Antimicrobial Compound | NB1000S is Sensitive | NB1000S MIC (ug/ml) | control MIC (ug/ml) |
|---|---|---|---|
| Meropenem | yes | 0.25 | 0.25 |
| Tigecycline | yes | 0.25 | 0.25 |
| Amoxicillin-clavulanate | yes | 0.5 | 0.25 |
| Tetracycline | yes | 0.5 | 0.5 |
| Clindamycin | yes | 0.5 | 0.12 |
| Imipenem | yes | 1 | 0.12 |
| Moxifloxacin | yes | 1 | 0.5 |
| Metronidazole | yes | 1 | 1 |
| Erythromycin | yes | 2 | 2 |
| Piperacillin-tazobactam | yes | 4 | 1 |
| Chloramphenicol | yes | 4 | 2 |
| Piperacillin | yes | 8 | 2 |
| Cefoxitin | yes | 8 | 2 |

**Table S2. Antibiogram**
NB1000S compared against progenitor B. vulgatus type strain DSM/1447 / ATCC 8482, obtained from DSMZ (DSM 1447 / ATCC 8482)

| Activity | Screen-ing[1] | Study Product Treatment | | | | | | | | | Follow-Up after Treatment[2] | | | | | | | | | | Extended Follow-Up[2] | |
|---|---|---|---|---|---|---|---|---|---|---|---|---|---|---|---|---|---|---|---|---|---|---|
| | | Inpatient Unit[3] | | | | | | | Outpatient | | Outpatient | | | | | | | | | | Outpatient | |
| **Unit/Clinic Visit #:** | **1** | **2** | **3** | **4** | **5** | **6** | **7** | **8** | **9** | **10** | | **11** | | | | | | | | | | **12[4]** |
| **Study Day #:** | | **1** | **2** | **3** | **4** | **5** | **6** | **7** | **10** | **14** | **17** | **21** | **24** | **28** | **35** | **42[4,5]** | **49** | **56** | **63** | **70[4]** | **77 to 182 Every 4 Wks[2,4]** | |
| **Window (±Days):** | | | **0** | **0** | **0** | **0** | **0** | **0** | **1** | **2** | **2** | **2** | **2** | **2** | **3** | **3** | **3** | **3** | **3** | **3** | **5** | **5** |
| Informed Consent | X | | | | | | | | | | | | | | | | | | | | | |
| Inclusion/Exclusion Criteria | X | Confirm | | | | | | | | | | | | | | | | | | | | |
| Medical History | X | Update | | | | | | | | | | | | | | | | | | | | |
| Height | X | | | | | | | | | | | | | | | | | | | | | |
| Body Weight and BMI Calculation | X | | | | | | | X | | X | | X | | | | | | | | | | X |
| Complete Physical Exam | X | | | | | | | | | | | | | | | | | | | | | |
| Targeted Physical Exam[6] | | X | | | | | | X | | X | | X | | | | | | | | | | X |
| Vital Signs[7] | X | X | X | X | X | X | X | X | | X | | X | | | | | | | | | | X |
| Urine Drug Screen | X | X | | | | | | | | | | | | | | | | | | | | |
| Blood Chemistry/Hematology/ESR/CRP | X | | | | | | | | | X | | X | | | | | | | | | | X |
| eGFR | X | | | | | | | | | X | | X | | | | | | | | | | X |
| Blood Coagulation | X | | | | | | | | | | | | | | | | | | | | | |
| Urinalysis | X | X | | | | | | | | X | | X | | | | | | | | | | X |
| Blood for Biomarkers | X | X | | | | | | | | X | | | | | | | | | | | | X |
| Pregnancy Testing[8] | X | X | | | | | | | | | | | | | | | | | | | | X |
| Stool qPCR Porphyran PUL | X | X | X | X | X | X | X | X | X | X | X | X | X | X | X | X | X | X | X | X | X | X |
| Stool qPCR NB1000S | X | X | X | X | X | X | X | X | X | X | X | X | X | X | X | X | X | X | X | X | X | X |
| Stool for Oxalate (Stored, possible assay) | | X | X | X | X | X | X | X | X | X | X | X | X | X | X | X | X | X | X | X | | X |
| Randomization/Assignment | | X | | | | | | | | | | | | | | | | | | | | |
| Dispense/Review Study Product Administration Instructions | | X | X | X | X | X | X | X | X | X | | | | | | | | | | | | |
| Study Product Administration[9] | | Treatment for 14 Days | | | | | | | | | | | | | | | | | | | | |
| Adverse Event (AE) Recording[10] | X | X | X | X | X | X | X | X | X | X | | X | | | | | | | | | | X |
| GI Symptom Grading[11] | X | X | | | | | | X | | X | | X | | | | | | | | | | X |
| Prior and Concomitant Meds[12] | X | X | X | X | X | X | X | X | X | X | | X | | | | | | | | | | X |
| Prior and Concomitant Procedures/Treatments | X | X | X | X | X | X | X | X | X | X | | X | | | | | | | | | | X |

### Table S3. Schedule of Study Activities for Stage 1 (Healthy Volunteer Subjects)

AE = adverse event, BP = blood pressure, BMI = body mass index, CRP = C-reactive protein, eGFR = estimated glomerular filtration rate, ESR = erythrocyte sedimentation rate, ET = early termination, GI= gastrointestinal, hCG = beta-human chorionic gonadotropin, PUL = polysaccharide utilization locus, qPCR = quantitative polymerase chain reaction, Wks = weeks.

1. Screening period is up to 35 days for Stage 1.
2. The duration of follow-up is determined on an individual subject basis. All groups including Control Groups A and C who receive no NB1000S will have ≥ 28 days of follow-up for safety reporting after the study product treatment period ends (to Day 42). Subjects who received NB1000S will be followed for ≥ 56 days after treatment cessation for monitoring of fecal shedding of NB1000S (to Day 70). Subjects who have ≥ 3 consecutive stool samples with fecal shedding of NB1000S ≤ LOD within 56 days after treatment cessation will be withdrawn from study at Day 70. All other subjects (identified as persistent shedding) will continue to be followed until either ≥ 2 consecutive stool samples show NB1000S shedding ≤ LOD or 168 days (24 weeks) after treatment cessation (to Day 182), whichever occurs first. Subjects with persistent shedding at 168 days after treatment cessation (or at ET) must be treated with an antibiotic for 7 days.
3. The subjects' admission to the inpatient unit is anticipated to occur on Day -1, the day before Visit 2 Day 1.
4. Visit 12 (end of study) activities to be completed for all subjects, regardless of day of withdrawal. Visit 12 activities to be completed for control Groups A and C on Day 42. Visit 12 to be completed on Day 70 for subjects withdrawing 56 days after treatment cessation. For subjects continuing on extended follow-up, Visit 12 will not be completed until day of withdrawal. All Visit 12 activities required as an early termination (ET) visit if subject withdraws prematurely.
5. Day 42 is last day on study for control Groups A and C.
6. The "targeted" physical examination is based on positives on a review of symptoms, previous physical examination findings, and adverse events (AEs).
7. Vital signs consist of blood pressure (BP), heart rate, respiratory rate, and body temperature. Subject should be seated for at least 3 minutes before measuring BP and heart rate.
8. Pregnancy testing only in women of childbearing potential. The Screening and Visit 2 Day 1 tests must be serum (not urine) beta-human chorionic gonadotropin (hCG), and the end of study test may be serum or urine. In addition, a urine pregnancy test maybe performed on Day 1 if needed because the turnaround time for results of the serum test is not sufficiently rapid.
9. Subjects assigned to NB2000P or placebo are dosed one or more times daily for 14 days. If assigned to NB1000S, NB1000S administered one-time on Day 1 or more often, according to treatment group.

10. The period of "adverse event" recording begins immediately after informed consent is given and continues until the subject is withdrawn from the study and, for those subjects who require antibiotic treatment at the end of the study because of persistent fecal shedding of NB1000S, until that antibiotic treatment is completed.
11. GI symptoms of abdominal pain, bloating, constipation, diarrhea, flatulence, nausea, and vomiting severity grading.
12. All concomitant medication taken within 30 days prior to the Screening visit and through the patient's entire time on study are to be record

| Activity | Screen-ing[1] | Study Product Treatment | | | | | Follow-Up after Treatment[2] | | | | | | | | | | Extended Follow-Up[2] | |
|---|---|---|---|---|---|---|---|---|---|---|---|---|---|---|---|---|---|---|
| **Clinic Visit #:** | 1 | 2 | 3[13] | 4[13] | 5[13] | 6[13] | | 7[13] | | | | | | | | | | 8[3] |
| **Study Day #:** | | 1 | 7 | 14 | 21 | 28 | 31 | 35 | 38 | 42 | 49 | 56 | 63 | 70 | 77 | 84[2,3] | 112 to 196 Every 4 Wks[2] | |
| **Window (±Days):** | | | 2 | 2 | 2 | 2 | 3 | 3 | 3 | 3 | 3 | 3 | 3 | 3 | 3 | 3 | 5 | 3 |
| Informed Consent | X | | | | | | | | | | | | | | | | | |
| Inclusion/Exclusion Criteria | X | Confirm | | | | | | | | | | | | | | | | |
| Medical History | X | Update | | | | | | | | | | | | | | | | |
| Height | X | | | | | | | | | | | | | | | | | |
| Body Weight and BMI Calculation | X | X | X | X | X | X | | X | | | | | | | | | | X |
| Complete Physical Exam | X | | | | | | | | | | | | | | | | | |
| Targeted Physical Exam[4] | | X | X | X | X | X | | X | | | | | | | | | | X |
| Vital Signs[5] | X | X | X | X | X | X | | X | | | | | | | | | | X |
| Blood Chemistry/Hematology/ESR/CRP | X | | | X | | X | | X | | | | | | | | | | X |
| eGFR | X | | | X | | X | | X | | | | | | | | | | X |
| Blood Coagulation | X | | | | | | | | | | | | | | | | | |
| Urinalysis | X | X | | X | | X | | X | | | | | | | | | | X |
| Blood for Biomarkers | X | X | | | | X | | | | | | | | | | | | X |
| Pregnancy Testing[6] | X | X | | | | | | | | | | | | | | | | X |
| 24-Hr Urine Oxalate and Other Analytes[7] | 2X7 | X | X | X | X | X[8] | | | | | | | | | | | | |
| Stool qPCR Porphyran PUL | X | X | X | X | X | X | X | X | X | X | X | X | X | X | X | X | X | X |
| Stool qPCR NB1000S | X | X | X | X | X | X | X | X | X | X | X | X | X | X | X | X | X | X |
| Randomization/Assignment | | X | | | | | | | | | | | | | | | | |
| Dispense/Review Study Product Administration Instructions | | X | X | X | X | X | | | | | | | | | | | | |
| Study Product Administration[9] | | Treatment for 28 Days | | | | | | | | | | | | | | | | |
| Adverse Event (AE) Recording[10] | X | X | X | X | X | X | | X | | | | | | | | | | X |
| GI Symptom Grading[11] | X | X | X | X | X | X | | X | | | | | | | | | | X |
| Prior and Concomitant Meds[12] | X | X | X | X | X | X | | X | | | | | | | | | | X |
| Prior and Concomitant Procedures/Treatments | X | X | X | X | X | X | | X | | | | | | | | | | X |

### Table S4. Schedule of Study Activities for Stage 2 (EH Patients)

AE = adverse event, BP = blood pressure, BMI = body mass index, CRP = C-reactive protein, eGFR = estimated glomerular filtration rate, ESR = erythrocyte sedimentation rate, hCG = beta-human chorionic gonadotropin, PUL = polysaccharide utilization locus, qPCR = quantitative polymerase chain reaction, Wks = weeks.

1. Screening period is up to 35 days for Stage 2.
2. The duration of follow-up is determined on an individual patient basis. All patients will be followed for ≥ 56 days after treatment cessation for monitoring of fecal shedding of NB1000S (to Day 84). Patients who have ≥ 3 consecutive stool samples with fecal shedding of NB1000S ≤ LOD within the 56 days after treatment cessation will be withdrawn from study at Day 84. All other patients (identified as persistent shedding) will continue to be followed until either ≥ 2 consecutive stool samples show NB1000S shedding ≤ LOD or 168 days (24 weeks) after treatment cessation (to Day 196), whichever occurs first. Patients with persistent shedding at 84 days after treatment cessation (or at ET) must be treated with an antibiotic for 7 days at around 112 days after treatment cessation (or at ET) and have two follow-up stool samples for NB1000S qPCR.
3. Visit 8 (end of study) activities to be completed for all patients, regardless of day of withdrawal. Visit 8 activities to be completed on Day 84 for subjects withdrawing 56 days after treatment cessation. For subjects continuing on extended follow-up, Visit 8 activities will not be completed until day of withdrawal. All Visit 8 activities required as an early termination (ET) visit if patient withdraws prematurely.
4. The "targeted" physical examination is based on positives on a review of symptoms, previous physical examination findings, and adverse events (AEs).
5. Vital signs consist of blood pressure (BP), heart rate, respiratory rate, and body temperature. Patient should be seated for at least 3 minutes before measuring BP and heart rate.
6. Pregnancy testing only in women of childbearing potential. The Screening and Visit 2 Day 1 tests must be serum (not urine) beta-human chorionic gonadotropin (hCG), and the end of study test may be serum or urine. In addition, a urine pregnancy test maybe performed on Day 1 if needed because the turnaround time for results of the serum test is not sufficiently rapid.
7. The two 24-hour urine collections for urinary oxalate during the Screening period must be separated by at least 3 days. Analytes other than oxalate are pH, sodium, potassium, chloride, creatinine, calcium, citrate, magnesium, phosphorus, uric acid, and sulfate.

8. The last 24-hour urine collection must be completed no later than 24 hours after the last study product administration.
9. NOV-001 dose as determined from Stage 1. NB2000P dosed once or more times daily for 28 days. NB1000S is administered two times on Day 1.
10. The period of "adverse event" recording begins immediately after informed consent is given and continues until the subject is withdrawn from the study and, for those subjects who require antibiotic treatment at the end of the study because of persistent fecal shedding of NB1000S, until that antibiotic treatment is completed.
11. GI symptoms of abdominal pain, bloating, constipation, diarrhea, flatulence, nausea, and vomiting severity grading.
12. All concomitant medication taken within 30 days prior to the Screening visit and through the patient's entire time on study are to be recorded.
13. Visit may be conducted in-person or remotely, as appropriate and when feasible.

| Study Group: | A | B | C | 1 | 3* | 2 | 2X | All Stage 1 |
|---|---|---|---|---|---|---|---|---|
| NB1000S doses: | 0 | 1 | 0 | 1 | 1 | 1 | 2 | |
| NB2000P dose/day: | 0 | 0 | 10 | 0.5 | 2.5 | 10 | 20 | |
| Subjects / group: | 3 | 4 | 4 | 8 | 8 | 8 | 4 | |
| **Overall** | | | | | | | | |
| Mild | 2 | 3 | 3 | 14 | 7 | 20 | 6 | **55** |
| Moderate | | 2 | | 3 | 1 | 1 | | **7** |
| Severe | | | | 1[#] | | | | **1**[#] |
| Serious | | | | 1 | | | | **1** |
| **All TEAEs** | 2 | 5 | 3 | 18 | 8 | 21 | 6 | **63** |
| **Related**** | | | | | | | | |
| Mild | 2 | 3 | 1 | 8 | 1 | 9 | 4 | **28** |
| Moderate | | 2 | | 1 | | 1 | | **4** |
| Severe | | | | | | | | **0** |
| Serious | | | | | | | | **0** |
| **All Related TEAEs** | 2 | 5 | 1 | 9 | 1 | 10 | 4 | **32** |

***Table 12-1:***

**Table S5. Summary of TEAEs by Severity/Seriousness and Relatedness to Study Product, Stage 1**

Analysis of Stage 1 TEAEs by maximum severity and relatedness. Numbers in the Table refer to the total number of TEAEs in each group.

* NOV-001-treated study groups are shown in order of ascending dose of NB2000P

** For this binary analysis of TEAE relatedness, events were considered "Related" if the PI assessed the event as being Possibly Related, Probably Related, or Related to either NB1000S or to NB2000P; events assessed as Not Related and Unlikely Related were considered "Unrelated."

[#] The severe AE in Group 1 was also an SAE.

No TEAEs were reported for Group RC; this column is omitted from the in-text Tables.

| | | A | B | C | 1 | 3 | 2 | 2X |
|---|---|---|---|---|---|---|---|---|
| | **Study Group:** | **A** | **B** | **C** | **1** | **3** | **2** | **2X** |
| | NB1000S doses: | 0 | 1 | 0 | 1 | 1 | 1 | 2 |
| | NB2000P dose/day: | 0 | 0 | 10 | 0.5 | 2.5 | 10 | 20 |
| | Subjects / group: | 3 | 4 | 4 | 8 | 8 | 8 | 4 |
| **SOC** | **Preferred Term** | | | | | | | |
| Gastrointestinal | Abdominal distension | | | | 1 | | 2 | |
| | Abdominal pain | | | | 1 | | 2 | |
| | Abdominal pain upper | | 1 | | | | | |
| | Constipation | | 1 | | 3 | | | |
| | Diarrhea | | 1 | | 3 | 1 | 3 | 1 |
| | Dry mouth | | | 1 | | | | |
| | Eructation | | | | | | 1 | |
| | Flatulence | | | 1 | 3 | 2 | 4 | 1 |
| | GI sounds abnormal | | | | | | 1 | 1 |
| | Mouth swelling | | 1 | | | | | |
| | Nausea | | | | 2 | | 2 | |
| | Vomiting | 1 | | | | | 1 | |
| Infections* | Cellulitis | | | | | 1 | | |
| | Cystitis | | | | | | | 1 |
| | Pharyngitis streptococcal | | | | | | | 1 |
| Injury | Animal scratch | | | | | 1 | | |
| Musculoskeletal | Arthralgia | | | | 1 | | | |
| | Musculoskeletal chest pain | | | | 1 | | | |
| | Myalgia | | | | | 1 | | |
| | Pain in jaw | | 1 | | | | | |
| Nervous | Alcoholic seizure | | | | 1 | | | |
| | Anosmia | | | | | | 1 | |
| | Headache | 1 | | | 1 | | 1 | |
| | Migraine | | | | | 1 | | |
| Renal | Glycosuria | | | 1 | | | | |
| Reproductive | Pelvic pain | | | | 1 | | | |
| Respiratory | Sinus congestion | | | | | | 1 | |
| | Upper-airway cough syndrome | | | | | | 1 | |
| Vascular | Hypertension | | | | | 1 | | |
| | **Any TEAE (N):** | 1 | 3 | 3 | 7 | 6 | 7 | 1 |
| | **Any TEAE (%):** | 33.3 | 75.0 | 75.0 | 87.5 | 75.0 | 87.5 | 25.0 |

**Table S6. Number of Subjects with ≥ 1 TEAE, Stage 1**

Stage 1 TEAEs by type of event (MedDRA System Organ Classification and Preferred Term), number of individuals in each Group with ≥1 of each PT event.. AEs by PT were counted only once for a given subject within each PT and SOC.

* Abbreviations: Infections – Infections and Infestations; Injury – Injury, Poisoning and procedural complications; Musculoskeletal - Musculoskeletal and connective tissue disorders; Renal – Renal and urinary disorders; Reproductive – Reproductive and breast disorders; Respiratory – Respiratory, thoracic and mediastinal disorders; GI – Gastrointestinal.
NOV-001 treated groups are shown in order of ascending dose of NB2000P

| Study Group:<br>Patients (N): | Active<br>9 | Placebo<br>3 | All Patients<br>12 |
|---|---|---|---|
| **Overall TEAEs** | | | |
| Mild | 40 | 7 | 47 |
| Moderate | 6 | 3 | 9 |
| Severe | 0 | 0 | 0 |
| Serious | 0 | 0 | 0 |
| **All TEAEs** | 46 | 10 | 56 |
| **Related* TEAEs** | | | |
| Mild | 19 | 4 | 23 |
| Moderate | 4 | 0 | 4 |
| Severe | 0 | 0 | 0 |
| Serious | 0 | 0 | 0 |
| **All Related TEAEs** | 23 | 4 | 27 |

**Table S7. Summary of TEAEs by Severity/Seriousness and Relatedness to Study Product, Stage 2**

Analysis of Stage 2 TEAEs by maximum severity and relatedness. Numbers in the Table refer to the total number of TEAEs in each group.

| | Study Group: | Active | Placebo | All Stage 2 |
|---|---|---|---|---|
| | Patients (N): | 9 | 3 | 12 |
| **SOC** | **Preferred Term** | | | |
| Gastrointestinal | Abdominal distension | 4 | 1 | 5 |
| | Abdominal pain | 3 | | 3 |
| | Constipation | 1 | 2 | 3 |
| | Diarrhea | 4 | 1 | 5 |
| | Eructation | 1 | | 1 |
| | Flatulence | 7 | 3 | 10 |
| | Nausea | 3 | 1 | 4 |
| | Vomiting | 1 | | 1 |
| General | Influenza like illness | 1 | | 1 |
| | Oedema peripheral | 1 | | 1 |
| Infections | Influenza | | 1 | 1 |
| | Urinary tract infection | 1 | | 1 |
| Investigations | C-reactive protein increased | 1 | | 1 |
| | Sex hormone level abnormal | 1 | | 1 |
| | Hepatic enzyme increased | 1 | | 1 |
| Musculoskeletal | Arthralgia | 1 | | 1 |
| Nervous | Hypoaesthesia | 1 | | 1 |
| | Migraine | 1 | | 1 |
| | Paraesthesia | 1 | | 1 |
| Skin | Rash | 1 | | 1 |
| | **Any TEAE (N):** | 8 | 3 | 11 |
| | **Any TEAE (%):** | 88.9 | 100 | 91.7 |

**Table S8. Number of Subjects with ≥ 1 TEAE, Stage 2**
Analysis of Stage 2 TEAEs by type of event (MedDRA System Organ Classification and Preferred Term), number of individuals in each Group with ≥1 of each PT event. AEs by PT were counted only once for a given subject within each PT and SOC.

| Strain name | Human subject | Description | Partner organism | HGT transfer |
|---|---|---|---|---|
| sJLG0015 | 110-204 | pre-HGT strain | B. vulgatus | - |
| sJLG0016 | 117-201 | pre-HGT strain | B. vulgatus | - |
| sJLG0017 | 121-212 | pre-HGT strain | B. vulgatus | - |
| sJLG0022 | 110-204 | post-HGT strain | B. vulgatus | outgoing 561 kb |
| sJLG0023 | 117-201 | post-HGT strain | B. vulgatus | outgoing 259 kb |
| sJLG0024 | 125-202 | post-HGT strain | B. dorei | incoming 724 kb |
| sJLG0025 | 121-212 | post-HGT strain | B. vulgatus | outgoing 216 kb |
| sJLG0026 | 125-204 | post-HGT strain | B. dorei | incoming 667 kb |
| sJLG0027 | 125-204 | post-HGT strain | B. vulgatus | outgoing 222 kb |

**Table S9. Strains isolated from Phase 2 patients.**
